# AI Implementation in U.S. Hospitals: Regional Disparities and System-Level Implications

**DOI:** 10.1101/2025.06.27.25330441

**Authors:** Yeon-Mi Hwang, Madelena Y. Ng, Malvika Pillai, Michelle P. Sahai, Tina Hernandez-Boussard

## Abstract

AI has the potential to improve healthcare delivery, but uneven geographic adoption and implementation can reinforce existing care gaps and inefficiencies. We analyzed data from 3560 U.S. hospitals using the 2023 American Hospital Association (AHA) Annual Survey, 2023-2024 AHA IT Supplement, community-level socioeconomic indicators, and 2023-2025 Center for Medicare & Medicaid Services (CMS) hospital quality metrics to assess: where AI is implemented, what factors are associated with implementation, and patterns of early AI adoption across geographic regions. We found that hospital AI implementation is significantly clustered, with geographic hotspots and coldspots of adoption. Regions with higher healthcare access need indicators were less likely to have hospitals with AI-based predictive models. Geographically weighted regression reveals that factors associated with AI implementation vary by region, suggesting that adoption patterns reflect diverse local contexts and institutional characteristics. These findings provide a baseline understanding of early AI deployment patterns in U.S. hospitals. Future efforts should develop standardized, detailed, and model-specific AI implementation metrics and consider local context rather than pursuing uniform deployment strategies.

## 1. Introduction

Hospitals across the United States (U.S.) are rapidly adopting artificial intelligence (AI) and machine learning (ML) technologies to enhance clinical decision-making and streamline operations.^1–3^ However, this technological revolution is occurring unevenly across regions and institutions.^2,4^

Disparities in digital health engagement are well documented at both the patient and institutional level. Patients from lower socioeconomic groups tend to have lower digital health literacy^5^ and older adults are less likely to use digital health tools compared with younger populations.^6^ At the institutional level, high-resource hospitals are more likely to successfully implement and evaluate AI systems than those with low resources.^2^ This institutional disparity is particularly important because prior research has shown that hospitals with advanced digital infrastructure achieve better safety and quality outcomes,^7^ suggesting that uneven AI adoption could affect patient care. Recent surveys provide further context about the institution-level AI implementation and related challenges. Poon et al. ^8^ examined generative AI use across nonprofit health systems and found that AI applications had mixed results due to underdeveloped tools, funding issues, and unclear regulations. This suggests that AI implementation faces challenges beyond just resource disparities.

Building on these institutional-level findings, recent work by Chen and Yan ^4^ using the 2023 American Hospital Association (AHA) Information Technology (IT) Supplement data show that hospitals in areas with greater socioeconomic deprivation were significantly less likely to implement predictive AI models and reveal uneven distribution of AI implementation patterns across the U.S. Such geographic disparities are particularly concerning because they compound existing health disparities. For example, rural areas have higher percentages of elderly residents and lower-income populations, and already experience worse health outcomes.^9,10^ When these communities have lower rates of AI adoption, existing disparities may be further amplified. Our prior research has further demonstrated that AI implementation varies at the county level and correlates with regional health outcomes, suggesting this technological disparity may translate into differences in care quality and patient outcomes.^11^ Ensuring consistent healthcare quality and accessibility remains a critical public health objective, making it essential to understand geographic patterns in AI implementation.

To address this gap, we conducted a nationwide geospatial analysis of AI implementation patterns across U.S. hospitals, integrating multiple data sources to examine both the geography of AI adoption and its relationship to healthcare delivery patterns. We hypothesized that AI implementation follows a disproportionate geographic distribution with identifiable clusters of high and low AI implementation. Using geospatial analytical techniques with data from 2023-2024 AHA, area-level socioeconomic indicators, community characteristics, and Centers for Medicare & Medicaid Services (CMS) quality metrics, we aimed to: (1) characterize the spatial distribution of hospital AI implementation and identify specific geographic clusters, hotspots, and coldspots that can inform targeted research and policy discussion; (2) examine geographic variation in factors associated with AI implementation to understand regional variation patterns; (3) assess relationships between indicators of community need and AI implementation patterns; and (4) explore potential associations between AI adoption and hospital quality performance patterns. This study provides a snapshot of early AI adoption. Rather than simply documenting the disparities, this framework offers geographic targets, baseline data for evaluating AI deployment, and insights into regional implementation contexts that can inform future research and policy discussion.

## 2. Method

### 2.1 Data Source

This study utilized data from the 2023 American Hospital Association (AHA) Annual Survey and 2023-2024 Information Technology (IT) Supplement.^12^ The AHA Annual Survey collects comprehensive information on institutional characteristics and capabilities from hospitals nationwide. IT Supplement specifically focuses on IT infrastructure, applications, and operational capacities. The AHA Data previously included basic questions on predictive modeling, the 2023 version expanded to include detailed questions on predictive modeling for the first time, covering use cases, development approaches, and evaluation methodologies.^2^ The 2024 version further expanded coverage by adding a few additional predictive model evaluation questions and large language model (LLM) integration into electronic health records. Our analysis included all hospital facilities that responded to either the 2023 or 2024 AHA IT Supplement. To assess potential selection bias, we compared characteristics of survey respondents versus non-respondents (Table S1).

### 2.2 Variables

#### 2.2.1. Hospital Predictive Model and AI Implementation Measures

Hospitals were classified into three categories based on their predictive modeling capabilities using data from the 2023-2024 AHA IT Supplement: no predictive models (0), non-AI predictive models (1), or AI predictive models (2). For hospitals that responded to both the 2023 and 2024 surveys, we used the 2024 response as it reflects the most recent implementation status. For descriptive statistics, hospitals that were survey respondents but had missing responses to this item were retained as ‘no response’ to provide transparency and to understand data patterns across groups, including non-response rates. For subsequent analyses, a small number of hospitals with missing responses were recoded as having no predictive models (assigned a value of 0), based on the assumption that non-response from the survey respondent likely indicated lack of implementation. As this imputation can cause potential bias, we conducted sensitivity analyses to test the impact. This AI implementation level served as our primary variable of interest: it was used as a dependent variable when examining associations with geographic and institutional factors, and as an independent variable when assessing relationships with quality metrics.

We developed three exploratory secondary measures to capture broader aspects of implementation, as validated composite measures for these specific aspects of AI implementation were not available in the existing literature. These secondary measures assessed predictive model implementation breadth, development approach, and evaluation practices using structured scoring systems as proxy indicators for these implementation dimensions. All secondary measures incorporated the primary AI implementation score (0-2) as a foundational component, then added domain-specific points to create composite scores that were subsequently standardized to a 0-1 scale for comparability and to prevent range-driven inflation across different metrics. The breadth score added 0.25 points for each reported model use case. The development score assigned additional points based on model source: 1.5 points for EHR vendor-developed or self-developed models, 1.0 point for third-party solutions, and 0.5 points for public domain models, reflecting integration complexity and governance considerations. The evaluation score incorporated reported assessments of model accuracy and bias, with points assigned incrementally based on evaluation comprehensiveness across model portfolios. For hospitals responding to the 2024 survey, an enhanced evaluation score included post-implementation monitoring and governance accountability structures. Additionally, we calculated a separate LLM readiness score for 2024 respondents, measuring large language model integration status and implementation timeline. Throughout the manuscript, references to ‘the evaluation score’ correspond to the version based on survey items consistent across both years (referred to as the 2023 version in Table S2) to ensure comparability.

These scoring systems are exploratory and judgment-based rather than empirically validated, developed through author consensus and informed by published literature,^2,13–15^ and should be interpreted as relative indicators for comparative analysis rather than absolute measures of implementation quality or capability. The flexible framework allows other researchers to adjust weights and scoring criteria based on their specific research priorities and available data. Since survey questions underlying these secondary measures encompass both AI and non-AI predictive models, we use the term ‘model’ rather than ‘AI model’ when referring to these broader implementation measures (detailed scoring criteria and reasoning behind the scoring system in Table S2). In this manuscript, ‘model’ and ‘AI’ refer to predictive models only.

When referring to generative AI, we specify LLM.

#### 2.2.2. Hospital Characteristics

Hospital characteristics were obtained from the 2023 AHA Annual Survey dataset to serve as proxy indicators for institutional resource levels. While the 2024 AHA Annual Survey was not yet available at the time of analysis, hospital characteristics remain relatively stable over time and should have minimal impact on the analysis. We included organizational attributes such as children’s hospital, teaching hospital status, critical access hospital designation, rural referral center status, ownership type, bed capacity, health system affiliation, frontline hospital designation, Joint Commission accreditation status, presence of subsidiary hospitals, community hospital designation, and delivery system model. Detailed definitions and categorizations of these variables are provided in Table S3. We additionally calculated Core Index and Friction Index as developed by Strawley et al.,^16^ The Core Index measures adoption of foundational interoperability capabilities, with higher scores indicating greater interoperability. The Friction Index quantifies barriers to interoperability, with higher scores indicating greater barriers. We included these interoperability indices, calculated from AHA IT Supplement responses, as they efficiently summarize multiple dimensions of IT infrastructure and exchange capabilities that may influence a hospital’s readiness to implement AI systems.

#### 2.2.3. Geographic and Community Characteristics

Rural-urban classification and hospital geographic coordinates were obtained from the AHA Annual Survey. Hospitals were linked to their service area, county, state, census division, and census region using information from the same survey.

Area deprivation index (ADI)^17,18^ and social vulnerability indices (SVI)^19^ were first aggregated to five-digit zip codes and then to hospital service areas (HSAs) using a ZIP code-HSA crosswalk from Dartmouth Atlas.^20^ We used median values for this aggregation because deprivation indices typically include numerous small areas with similar socioeconomic profiles within each service region, making the median more robust to outliers while representing the typical deprivation level.

Healthcare Professional Shortage Area (HPSA) and Medically Underserved Area (MUA) scores were obtained from the Health Resources and Services Administration.^21^ These designations represent geospatial boundaries with scores reflecting the degree of healthcare shortage and underservice, respectively. We spatially joined these areas to HSAs wherever they overlapped. For hospital service areas intersecting with multiple HPSAs or MUAs, often with substantially different scores, we calculated mean values to capture the combined influence of all shortage designations. Hospitals without overlapping HPSA or MUA designations received a score of 0. We additionally collected county-level digital infrastructure metrics from 2023 U.S. Census Bureau data^22^ including: internet access percentage (households with any type of internet subscription), broadband percentage (households with high-speed internet subscription), and computing device percentage (households with at least one computing device such as desktop, laptop, tablet, or smartphone). These digital infrastructure variables were linked to hospitals based on their county location (Table S3).

#### 2.2.4. Hospital Quality Metrics

We obtained 70 continuous hospital quality of care metrics from the Centers for Medicare & Medicaid Services (CMS). These standardized metrics assess clinical performance, care quality, and patient safety across U.S. hospitals. For our longitudinal analyses, we collected quarterly data from 2022, 2023, 2024, and the first three quarters of 2025, yielding 15 measurement timepoints.

### 2.3 Analysis

#### 2.3.1 Descriptive statistics

We generated descriptive statistics comparing hospital and community characteristics across regions and AI implementation categories. Continuous variables are presented as means with standard deviations, while categorical variables are shown as frequencies with percentages. Statistical comparisons across groups were conducted using ANOVA for continuous variables and chi-square tests for categorical variables. ANOVA assumes normality within groups, homogeneity of variance, and independence of observations; chi-square tests assume independence and adequate expected cell frequencies. We assumed these assumptions were adequately met given the large sample size and the descriptive nature of the analysis. All statistical tests were two-sided with significance set at P<0.05. Bonferroni correction was applied for multiple testing correction. For the state-level analysis, states with less than 20 hospitals were dropped to avoid unreliable statistical estimates due to small sample sizes. This included Alaska, DC, Delaware, Hawaii, Maine, Rhode Island, Vermont, and Wyoming.

#### 2.3.2 Community Need and AI Implementation Pattern Analysis

We examined patterns between AI implementation and community need indicators through cross-tabulation analysis. This analysis was exploratory, examining the relationship between need and implementation based on our initial interest in geographic disparity patterns rather than testing a specific a priori hypothesis about where AI should be implemented. Given that AI implementation scores represented distinct and meaningful categories (no predictive models, non-AI predictive models, AI predictive models), we retained these original classifications to preserve interpretability.

For need indicators, we evaluated whether each measure’s distribution allowed for interpretable tertile categorization. Need measures that created distinct tertiles with adequate variation across categories were divided into tertiles to enable systematic comparison across need levels. However, for need indicators where many areas had the same values (particularly zero values), preventing adequate differentiation, we instead used binary categorization.

We examined Primary Healthcare Professional Shortage Area (HPSA), Dental HPSA, Mental HPSA, and Medically Underserved Area overall (MUA), MUA infant population component, MUA elderly population component, Area Deprivation Index (ADI), and Social Vulnerability Index (SVI) - Overall, SVI Theme 1, SVI Theme 2, SVI Theme 3, and SVI Theme 4 as need indicators.

We analyzed these indicators in two conceptually distinct groups: healthcare access need indicators (HPSA and MUA measures) represent areas with insufficient healthcare infrastructure and workforce, while socioeconomic need indicators (ADI and SVI) reflect community-level social and economic challenges. We do not assume that AI adoption will necessarily mitigate structural disparities or that higher AI adoption in high-need areas is inherently beneficial. These different types of need may require different AI solutions and implementation approaches, and inadequate AI design or implementation could potentially worsen existing disparities rather than reduce them.

It should be noted that AI implementation requires substantial technical, financial, and human resources, as well as institutional expertise in data governance, integration, and workflow adaptation. These observed patterns may reflect early-stage adoption constraints or institutional capacity differences rather than representing areas where AI should necessarily be prioritized.

#### 2.3.3 Spatial Clustering and Hotspot Analysis

##### 2.3.3.1 Geographic Visualization and Distribution

We visualized the geographic distribution of AI implementation through heatmaps across the U.S. and within individual census divisions. It should be noted that areas with higher hospital density may naturally show greater concentrations of AI implementation. Census divisions are categorized into nine regional groups (New England, Middle Atlantic, East North Central, West North Central, South Atlantic, East South Central, West South Central, Mountain, and Pacific).

##### 2.3.3.2 Spatial Clustering Analysis

We first assessed spatial clustering of hospitals using a nearest neighbor analysis using hospital coordinates, calculating the average nearest neighbor distance with a KDTree algorithm. To examine clustering of AI implementation, we calculated Global Moran’s I using projected hospital coordinates and AI implementation scores. We defined spatial relationships using the six nearest neighbors (k=6), which allowed detection of localized spatial clustering while maintaining statistically significant Moran’s I and z-score values (Figure S1). Spatial clustering effect size was measured using Moran’s I, ranging from −1 (perfect spatial dispersion/checkerboard pattern) to +1 (perfect spatial clustering). This test assumes that observations are conditionally independent given the spatial structure, which is a reasonable assumption at the national level. Census division and state level analyses were also conducted to account for potential variation in local dependencies and policy environments. Given prior evidence of clustering in hospital technology adoption, we conducted these tests to validate and localize spatial patterns at multiple geographic level. We report both unadjusted and adjusted p-values corrected for multiple comparisons using the Benjamini-Hochberg false discovery rate (BH FDR) procedure and Storey’s q-value method.

We applied DBSCAN clustering to identify geographically proximate hospitals with similar AI implementation patterns. These clusters serve as practical units for stakeholders to target interventions and develop region-specific approaches to AI implementation. DBSCAN assumes that meaningful clusters can be detected based on local point density and that standardized features (coordinates and AI scores) reflect relevant similarity. To address the uneven geographic distribution of hospitals, we computed local density estimates for each hospital using its k=6 nearest neighbors, then upweighted AI implementation scores in higher-density regions to enhance cluster detection while preserving DBSCAN’s density-based logic. All features were standardized before clustering. We selected the eps parameter (0.35) based on the inflection point where the number of clusters stabilized and proportion of noise remained acceptably low (Figure S2). As DBSCAN is a non-parametric clustering algorithm that does not produce p-values, no multiple testing correction was applied. For the state-level analysis, states with less than 20 hospitals were dropped to avoid unreliable statistical estimates due to small sample sizes. We note that DBSCAN may under-detect patterns in low-density rural regions.

##### 2.3.3.3 Hotspot Analysis

We complemented the clustering analysis with a Getis-Ord Gi* hotspot analysis to identify statistically significant spatial concentrations of high and low AI implementation. Hotspots can guide policymakers toward successful regional examples, while coldspots highlight areas needing targeted support and resources. Using projected hospital coordinates and AI implementation scores, we computed Gi* z-scores based on each hospital’s six nearest neighbors, consistent with our DBSCAN parameter. This analysis assumes that nearby hospitals may influence each other, and that any remaining differences are independent once geography is accounted for. It also assumes the AI scores are reasonably distributed for identifying unusual clusters. Hotspot (high implementation) and coldspot (low implementation) classifications were assigned based on z-score significance thresholds (90%, 95%, 99%). We report both unadjusted and adjusted p-values corrected for multiple comparisons using the BH FDR and Storey’s q-value method. For visualization purposes, we aggregated these classifications to the HSA level. For descriptive statistics, we further aggregated counts to state, census division, and national levels. For the state-level analysis, states with less than 20 hospitals were dropped to avoid unreliable statistical estimates due to small sample sizes.

#### 2.3.4 Predictive Modeling and Feature Importance Analysis

We employed a random forest regression model to identify the most influential factors predicting hospital AI implementation levels. Missing values were imputed using Multiple Imputation by Chained Equations (MICE) to account for systematic missingness patterns and preserve correlations between variables. The model used 100 trees with 5-fold cross-validation and incorporated both hospital-level characteristics and geospatial features as predictors. Model performance was assessed using R² calculated as the proportion of variance explained through 5-fold cross-validation. We calculated SHapley Additive exPlanations (SHAP) values to determine feature importance and understand the directional relationships between predictors and AI implementation. SHAP values quantify each feature’s contribution to individual predictions while accounting for feature interactions.^23^

#### 2.3.5 Geographically Weighted Regression Analysis

We implemented a geographically weighted regression to visualize how the association between hospital AI implementation scores and structural, socioeconomic, and technological factors vary across geographic space. Hospital coordinates were used to compute spatial weights based on Gaussian kernels. All continuous variables were standardized prior to analysis to allow for coefficient comparability. We used a bandwidth of 2 decimal degrees, calculated as approximately 222 km radius at mid-US latitudes.^24^ This bandwidth encompasses multiple counties or most smaller states, providing a good balance to capture meaningful regional patterns while ensuring sufficient local observations for stable coefficient estimation. The resulting spatially varying coefficients were mapped to assess regional heterogeneity. These results were interpreted descriptively to illustrate potential geographic variation, rather than as formal statistical inference.

#### 2.3.6 Longitudinal Analysis of Hospital Quality Trends by AI Implementation Status

We examined associations between AI implementation levels and hospital quality metrics using a longitudinal approach. Building on our prior county-level cross-sectional study that established preliminary patterns of associations between AI implementation and health outcomes,^11^ this analysis addresses previous methodological constraints by employing hospital-level data across 15 quarterly time points from 2022 to 2025 with adjustment for regional factors.

Important limitations must be acknowledged regarding this exploratory analysis. AI adoption in 2023 or 2024 was in early stages with limited deployment and often minimal integration into clinical workflows. Our data captured AI implementation status as of 2023 or 2024 but not precise implementation timing, and our AI implementation measure reflects broad categories without detailed information about specific applications, clinical targets, or utilization patterns. Additionally, observed associations may reflect underlying institutional readiness and capacity rather than direct AI effects, as hospitals with AI implementation may differ systematically in ways that independently influence quality performance.

Despite these constraints, this exploratory analysis serves several important purposes: advancing beyond our previous analysis to examine quality trajectory patterns across hospitals with different AI implementation levels, identifying potential areas where AI may be associated with quality differences over time, and providing additional insights about technology adoption and performance relationships. However, causal attribution cannot be made from these associations.

We analyzed quality metrics with less than 50% missingness in the third quarter 2025 report to balance data completeness with sample size. We strategically selected confounding variables to avoid high-dimensional modeling issues: the top three predictors of AI implementation from section 2.3.4 to control for systematic differences between hospitals, and the top five LASSO-identified outcome-specific predictors for each quality metric. We excluded mediating features such as interoperability capabilities from covariate selection.

We examined quality metric trajectories across 15 quarterly time points from 2022 to 2025 using linear mixed-effects models adjusted for the selected covariates, with Huber-White robust variance estimation to account for potential heteroscedasticity and departures from normality. Models included random intercepts for hospitals with fixed effect time (continuous), AI implementation level (categorical), and time-by-AI level interactions to estimate differential slopes, adjusting for selected covariates. Models were fitted using maximum likelihood estimation with AI level 0 (no predictive models) as the reference group. Effect sizes are reported as β coefficients representing the change in outcome per unit change in predictor. Model assumptions were assessed through Q–Q plots and the Shapiro–Wilk test of residuals. BH-FDR correction was applied for multiple testing.

## 3. Results

### 3.1 Hospital AI Implementation Patterns

Out of 6093 hospitals registered in AHA, 3560 hospitals (58.4%) responded to either 2023 or 2024 AHA IT Supplement (Table S1). Of them, 2155 hospitals (60.5%) responded to both years, 937 hospitals (26.3%) responded only in 2023 and 468 hospitals (13.1%) responded only in 2024. Since this could cause selection bias, we present descriptive statistics stratified by response pattern (Table S4). Among AHA IT Supplement respondent, 1043 (29.3%) reported using no predictive models, 568 (16.0%) employed non-AI predictive models, and 1738 (48.8%) utilized AI-based predictive models (Table 1). The remaining 5.9% (211 hospitals) did not report their AI implementation status and were classified as non-users in subsequent analyses. Sensitivity analysis showed this imputation did not change the significance or location of spatial clusters.

**Table 1.**
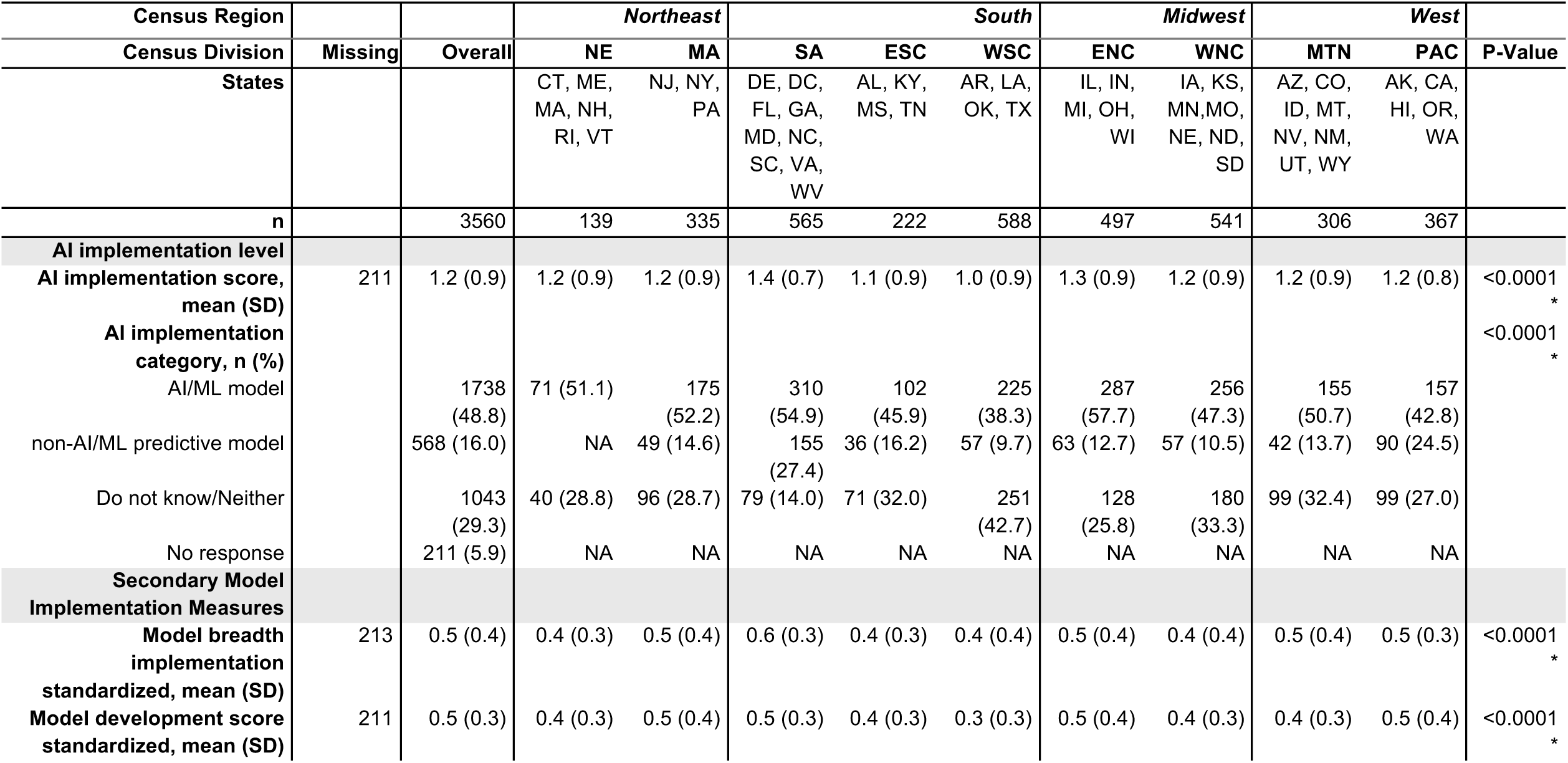
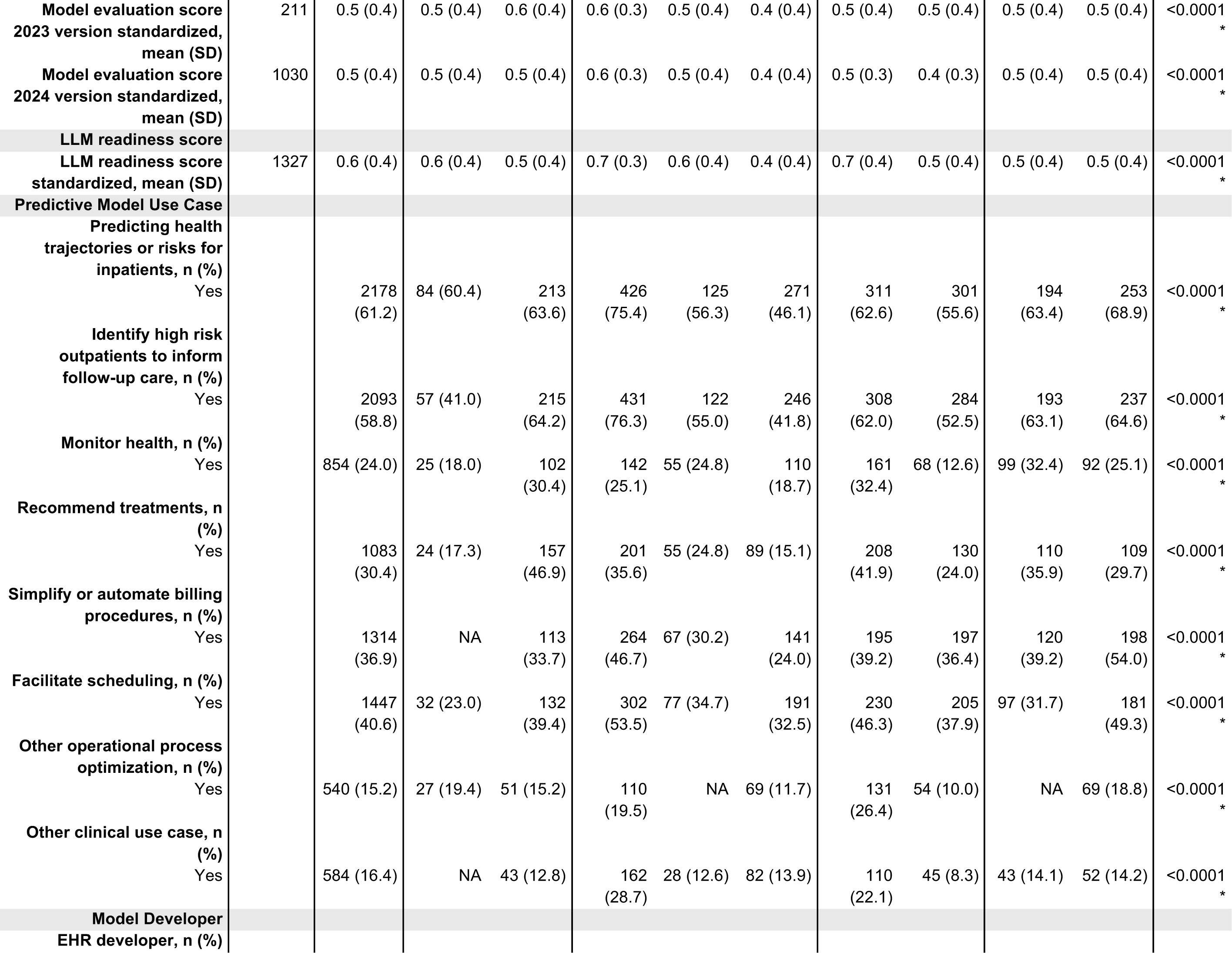
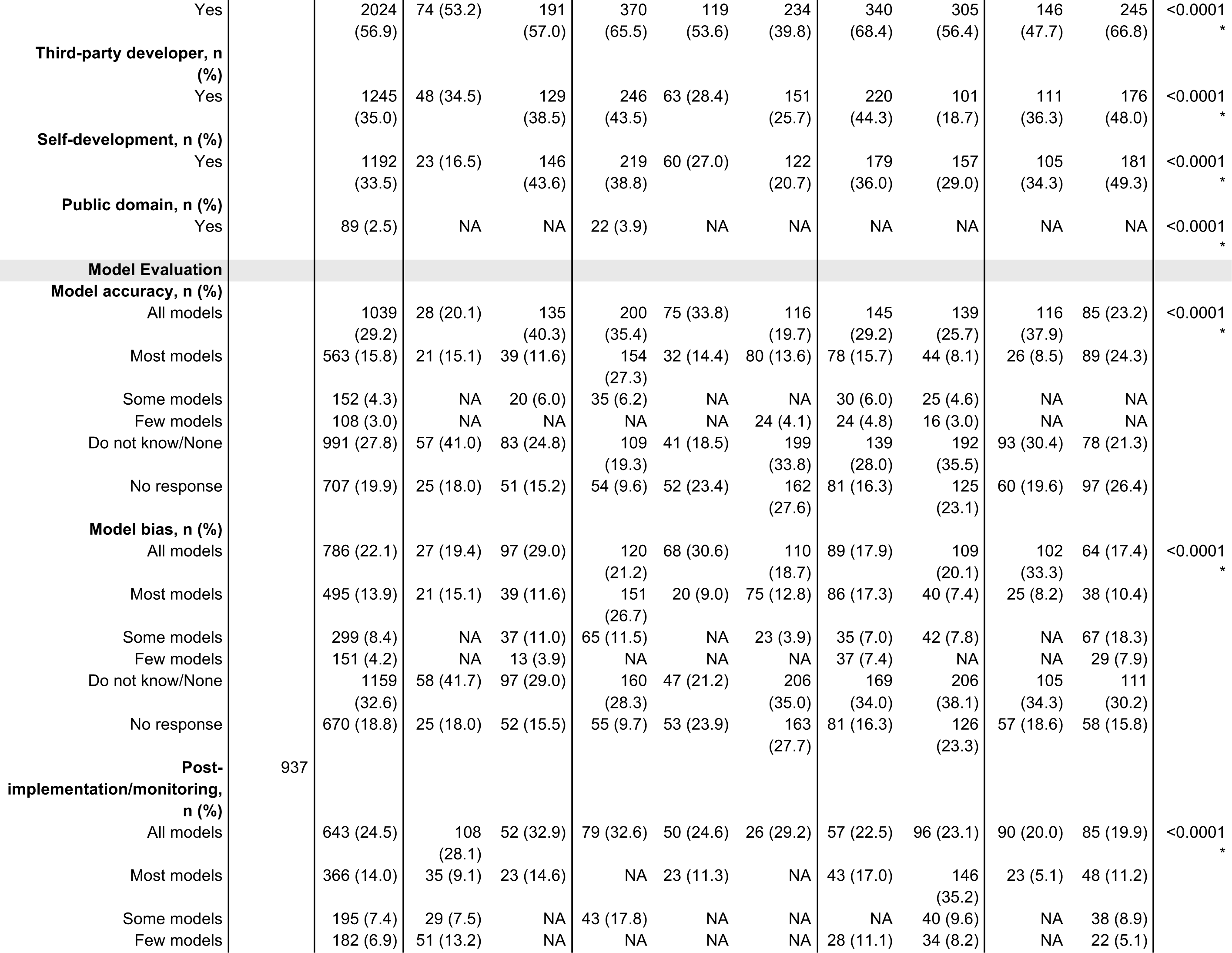
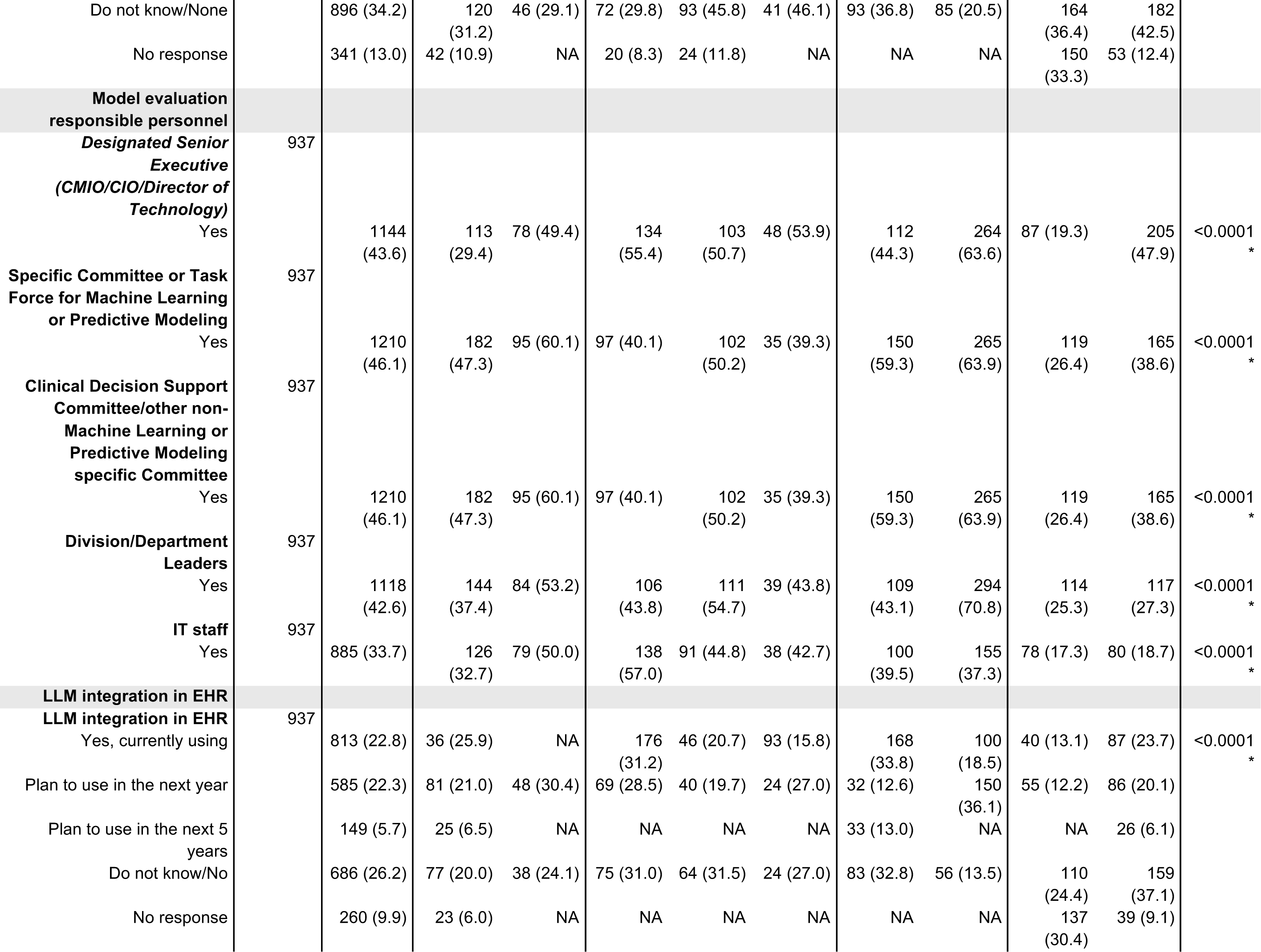
Descriptive statistics of AI and model implementation survey responses across census division. Census division abbreviations: NE=New England, MA=Mid Atlantic, SA=South Atlantic, ENC=East North Central, WNC=West North Central, ESC=East South Central, WSC=West South Central, MTN=Mountain, PAC=Pacific. State abbreviations: AL=Alabama, AK=Alaska, AZ=Arizona, AR=Arkansas, CA=California, CO=Colorado, CT=Connecticut, DE=Delaware, FL=Florida, GA=Georgia, HI=Hawaii, ID=Idaho, IL=Illinois, IN=Indiana, IA=Iowa, KS=Kansas, KY=Kentucky, LA=Louisiana, ME=Maine, MD=Maryland, MA=Massachusetts, MI=Michigan, MN=Minnesota, MS=Mississippi, MO=Missouri, MT=Montana, NE=Nebraska, NV=Nevada, NH=New Hampshire, NJ=New Jersey, NM=New Mexico, NY=New York, NC=North Carolina, ND=North Dakota, OH=Ohio, OK=Oklahoma, OR=Oregon, PA=Pennsylvania, RI=Rhode Island, SC=South Carolina, SD=South Dakota, TN=Tennessee, TX=Texas, UT=Utah, VT=Vermont, VA=Virginia, WA=Washington, WV=West Virginia, WI=Wisconsin, WY=Wyoming. Model scores and variables are defined in Tables S1 and S2. Cells with counts below 20 were suppressed to prevent re-identification through cross-tabulation; neighboring cells were also suppressed as needed. P-values were calculated using chi-square tests for categorical variables and ANOVA for continuous variable

Table 1 and Figure 1 presents AI implementation levels stratified by U.S. census division. Predictive models were most commonly utilized to predict patient health trajectories (61.2%), with the majority developed by hospitals’ EHR vendors (56.9%). The majority of hospitals reported no evaluation or no reponse for model accuracy (47.4%) and bias (51.4%).

**Figure 1.**
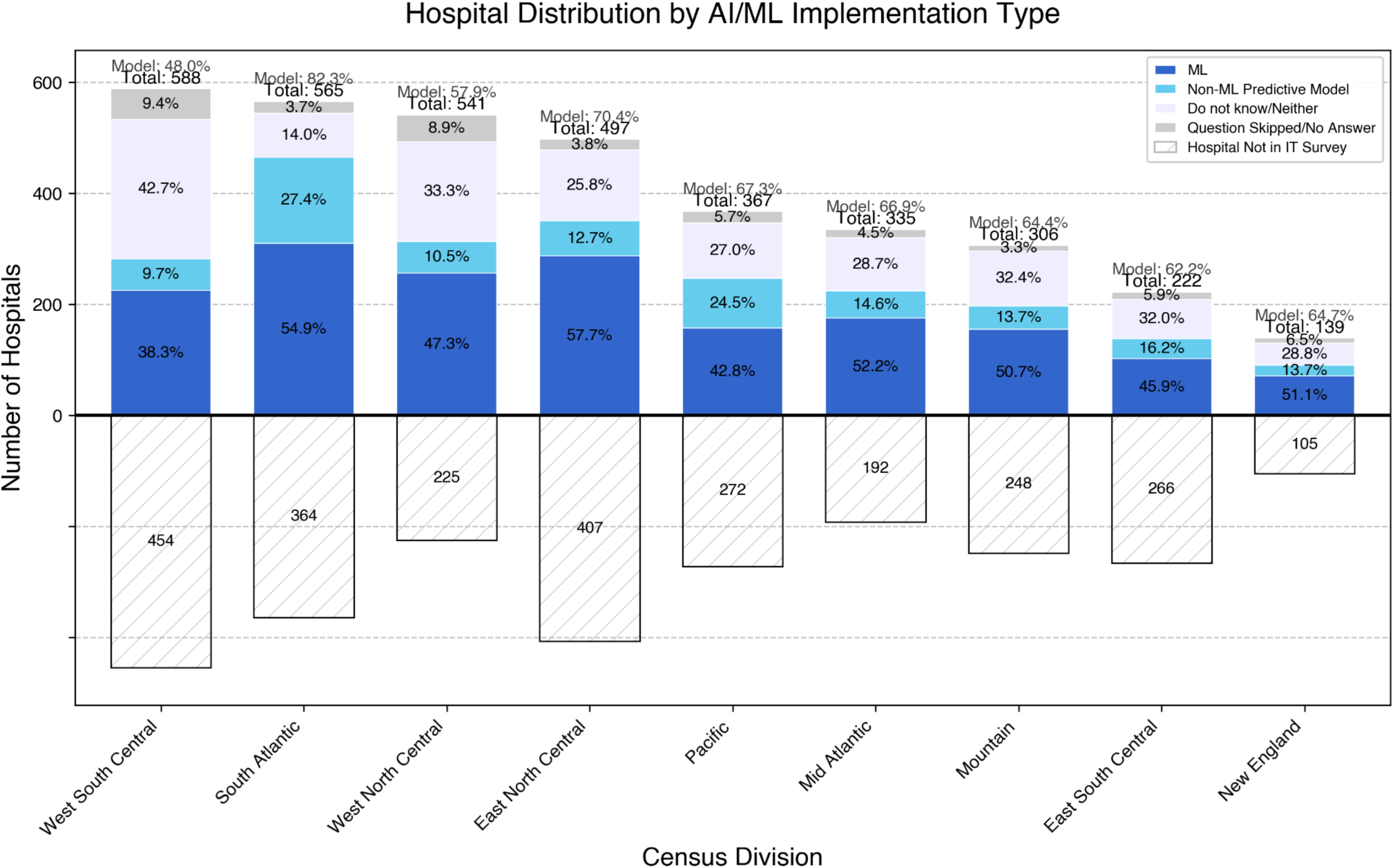
Hospital distribution by AI/ML implementation type across U.S. census divisions. Stacked bars show the proportion of hospitals with AI-based predictive models (lavender), non-AI predictive models (light blue), or no predictive models (dark blue). Gray segments indicate hospitals reporting “do not know/neither” or skipping the survey item. The total number of hospitals in the IT survey and the overall adoption rate (“Model: xx%”) are shown above each bar. Numbers of hospitals registered in the AHA but not included in the IT survey are displayed as striped bars below the x-axis. AI/ML adoption varied substantially across regions, with the South Atlantic division showing the highest adoption and the West South Central division the lowest.

Geographic variation was notable across census divisions and states (Figure 1). At the census division level, the South Atlantic division showed the highest base AI implementation scores (1.4). At the state level, South Carolina and South Dakota had the highest scores (1.8). In contrast, Idaho and Montana were lowest (0.6). Detailed scores and statistical comparisons by census division and state are provided in Table 1, S5, and S6.

Descriptive statistics for hospital and community characteristics are stratified by census division (Table S5), by state (Table S6), and by hospital AI implementation level (Table S7). Most characteristics differed significantly across geographic regions. However, when stratified by AI implementation level, hospital Medicaid inpatient proportion, overall SVI, and SVI theme 1,2, showed no statistically significant differences in distribution.

Heatmap analysis revealed consistent concentration of AI implementation in metropolitan regions across all census divisions (Figure S3). However, these concentration patterns partly reflect the underlying clustering of hospitals themselves, as confirmed by our nearest neighbor analysis showing significant hospital clustering (Nearest Neighbor Ratio: 0.419, z-score: −66.34, P<0.0001).

### 3.2 Community Need and AI Implementation Pattern Analysis

Cross-tabulation analysis revealed that implementation ratios varied widely across healthcare need indicators, ranging from 0.40 to 1.13 across all measures (Figure 2), with healthcare access need indicators consistently showing lower ratios compared to socioeconomic disadvantage measures. Tertile distribution is provided in Table S8. Socioeconomic disadvantage measures and Mental HPSA were analyzed using tertile distributions.

**Figure 2.**
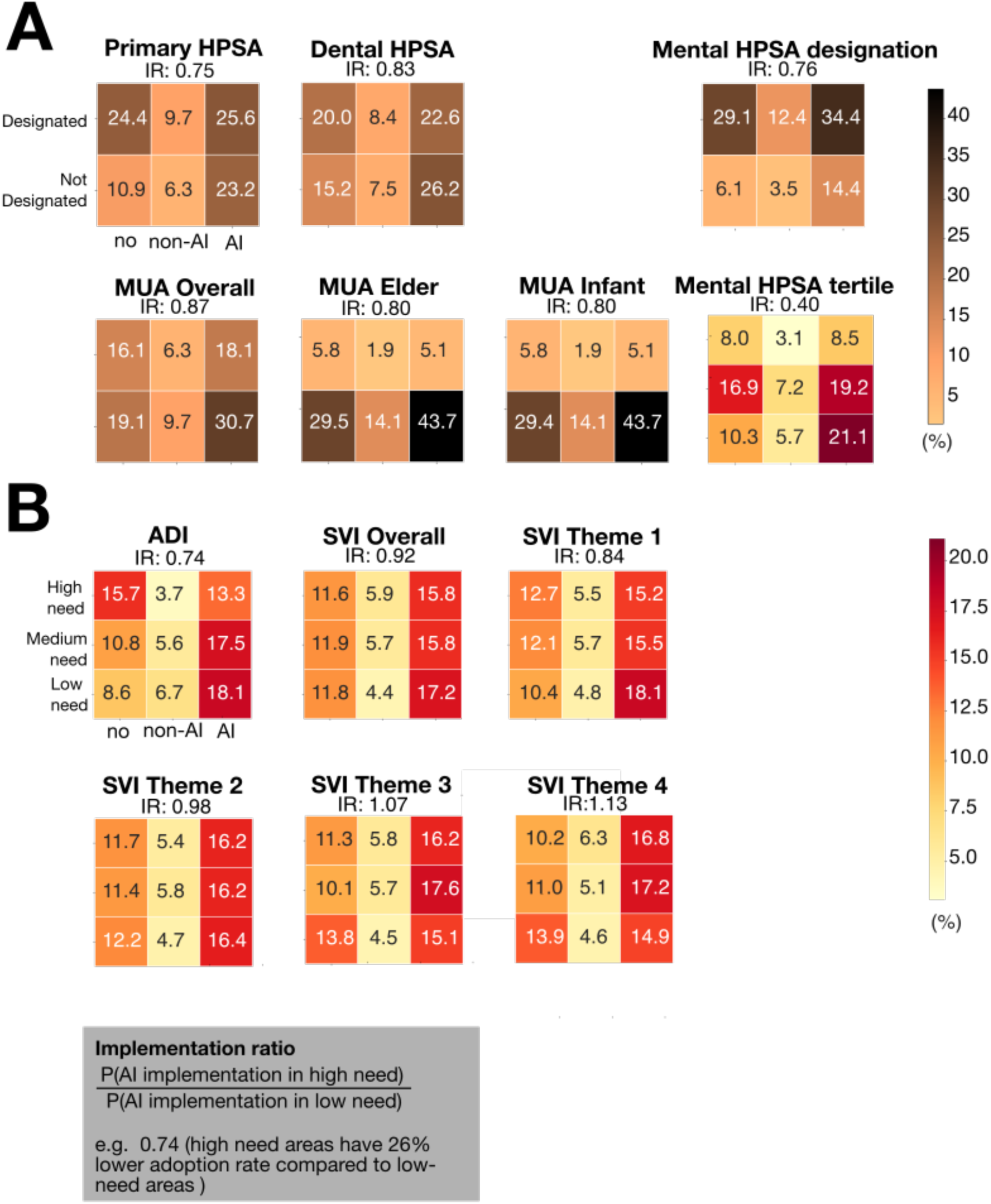
Alignment between community need and AI implementation. AI Artificial intelligence; HPSA Healthcare professional shortage area; MUA Medical underservice area, SVI Social Vulnerability Index Cross-tabulation analysis showing the distribution of hospitals across AI implementation categories (no AI, non-AI predictive models, AI predictive models) stratified by healthcare need levels. Panel A shows healthcare access need indicators including Health Professional Shortage Areas (HPSA) and Medically Underserved Areas (MUA). Panel B shows socioeconomic disadvantage indicators including Area Deprivation Index (ADI) and Social Vulnerability Index (SVI) themes. Implementation ratios (IR) represent the probability of AI implementation in high-need areas relative to low-need areas, with values <1.0 indicating lower implementation in high-need areas. Colors represent the percentage of hospitals in each category, with darker colors indicating higher percentages. Mental HPSA is shown in both binary designation and tertile formats. Table S8 present tertile threshold.

Mental Health Professional Shortage Areas demonstrated the most pronounced disparities, with the tertile version showing an implementation ratio of 0.40 and the binary designation version showing a ratio of 0.76. Other healthcare access indicators showed similar patterns: Primary Care HPSA (IR: 0.75), Dental HPSA (IR: 0.83), MUA Overall (IR: 0.87), MUA Elder (IR: 0.80), and MUA Infant (IR: 0.80).

Socioeconomic disadvantage need measures showed more variable and generally less pronounced disparities. The ADI had an implementation ratio of 0.74. SVI measures demonstrated different patterns across themes: Theme 1 (IR: 0.84), Theme 2 (IR: 0.98), Theme 3 (IR: 1.07), and Theme 4 (IR: 1.13), with the overall SVI showing near-equitable implementation (IR: 0.92). Notably, SVI Themes 3 and 4 showed slightly higher AI adoption rates in high-need areas, contrasting with the consistent under-implementation seen across all healthcare access indicators.

### 3.3 Spatial Clustering and Hotspot Analysis

#### 3.3.1. Detection of Spatial Clustering Patterns

Moran’s I analysis revealed significant spatial autocorrelation for all model implementation metrics (P<0.0001; Table S9). Regional analysis confirmed clustering in most census divisions, with variation in degree of clustering across regions and states (Table S9).

#### 3.3.2. Identification of Density-Based Clusters

The adaptive DBSCAN clustering algorithm identified 434 distinct clusters of hospital AI implementation at the base level across 3328 hospitals nationwide (91.93%). Clusters averaged 12.2 hospitals per cluster, with regional variation in cluster size. The Pacific region exhibited the largest average cluster size (27.4 hospitals), while the East South Central region had the smallest clusters (4.6 hospitals). Complete DBSCAN clustering results for all model implementation metrics are presented in Table S10.

#### 3.3.3. Hotspot and Coldspot Analysis

Hospital-level analysis identified statistically significant 658 hotspots and 570 coldspots of AI implementation when unadjusted for multiple testing. Multiple testing correction substantially reduced significant findings, with Storey’s q-value method retaining approximately half of the hotspots/coldspots while BH FDR correction eliminated nearly all significant results. At the hospital service area level (n=2191), we detected 378 hotspots and 408 coldspots for AI base score (Figure 3). The South Atlantic census division exhibited the highest mean z-score across all AI model implementation measures. At the state level, South Carolina led for base score and evaluation score. North Carolina led for breadth score and development score. Complete hotspot analysis results are presented in Figure S4-S7 and Table S11-S18.

**Figure 3.**
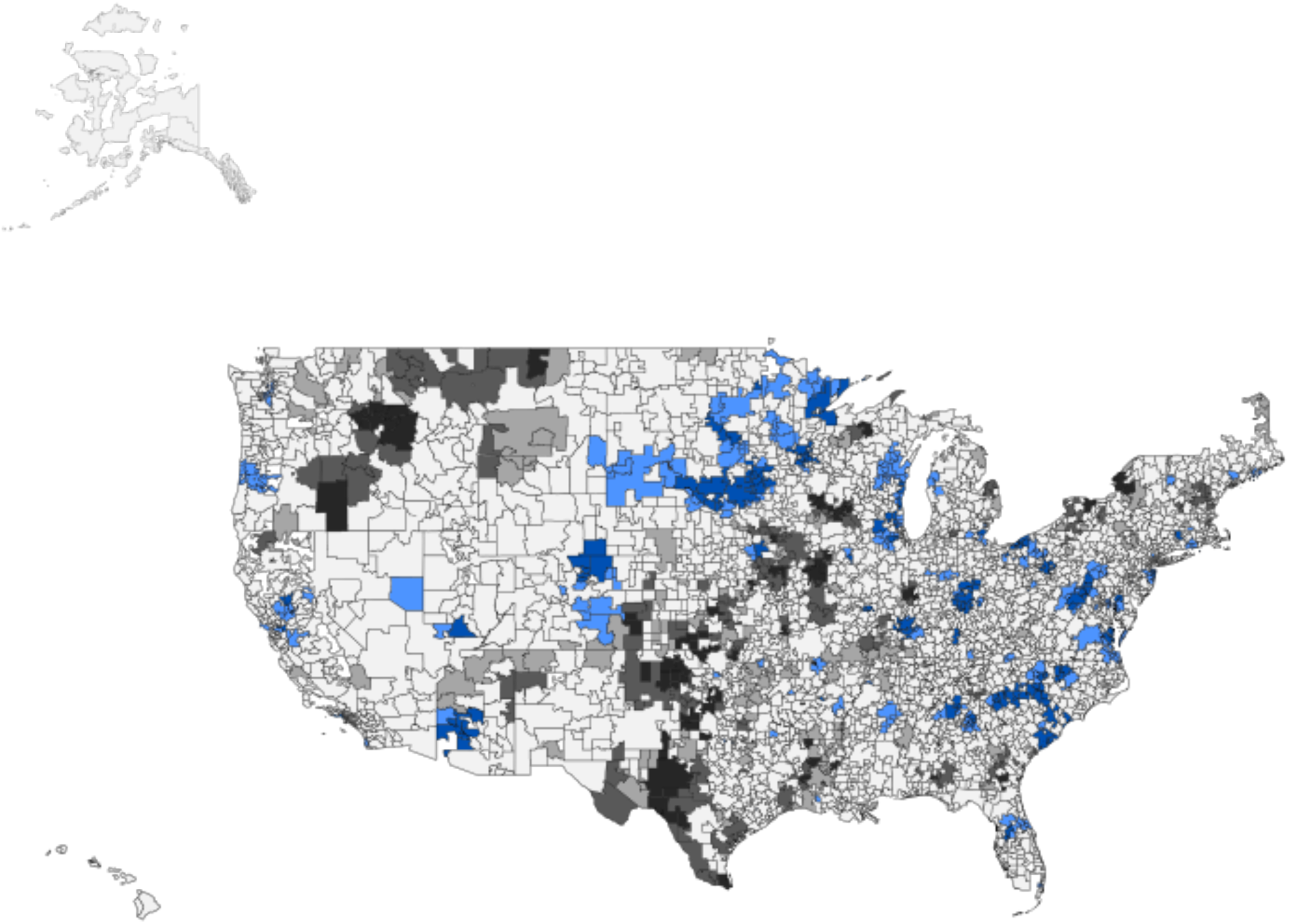
Geographic distribution of AI implementation hotspots and coldspots across U.S. hospital service areas. Hotspots (blue) indicate areas of significantly higher AI adoption, and coldspots (black) indicate areas of significantly lower adoption, based on Getis-Ord Gi* statistics. Results shown here are unadjusted

### 3.4 Predictive Modeling and Factors Associated with AI Implementation

To identify key factors associated with hospital AI implementation, we developed a random forest classification model incorporating hospital-level, geographic, and community characteristics. The model achieved strong predictive performance with a mean R² of 0.61 [0.60, 0.61] across 5-fold cross-validation. Figure 4a presents the ten most influential predictors of AI implementation based on SHAP values. The analysis revealed that both institutional and regional factors serve as important features, with seven hospital-level characteristics and three regional factors among the top predictors.

**Figure 4.**
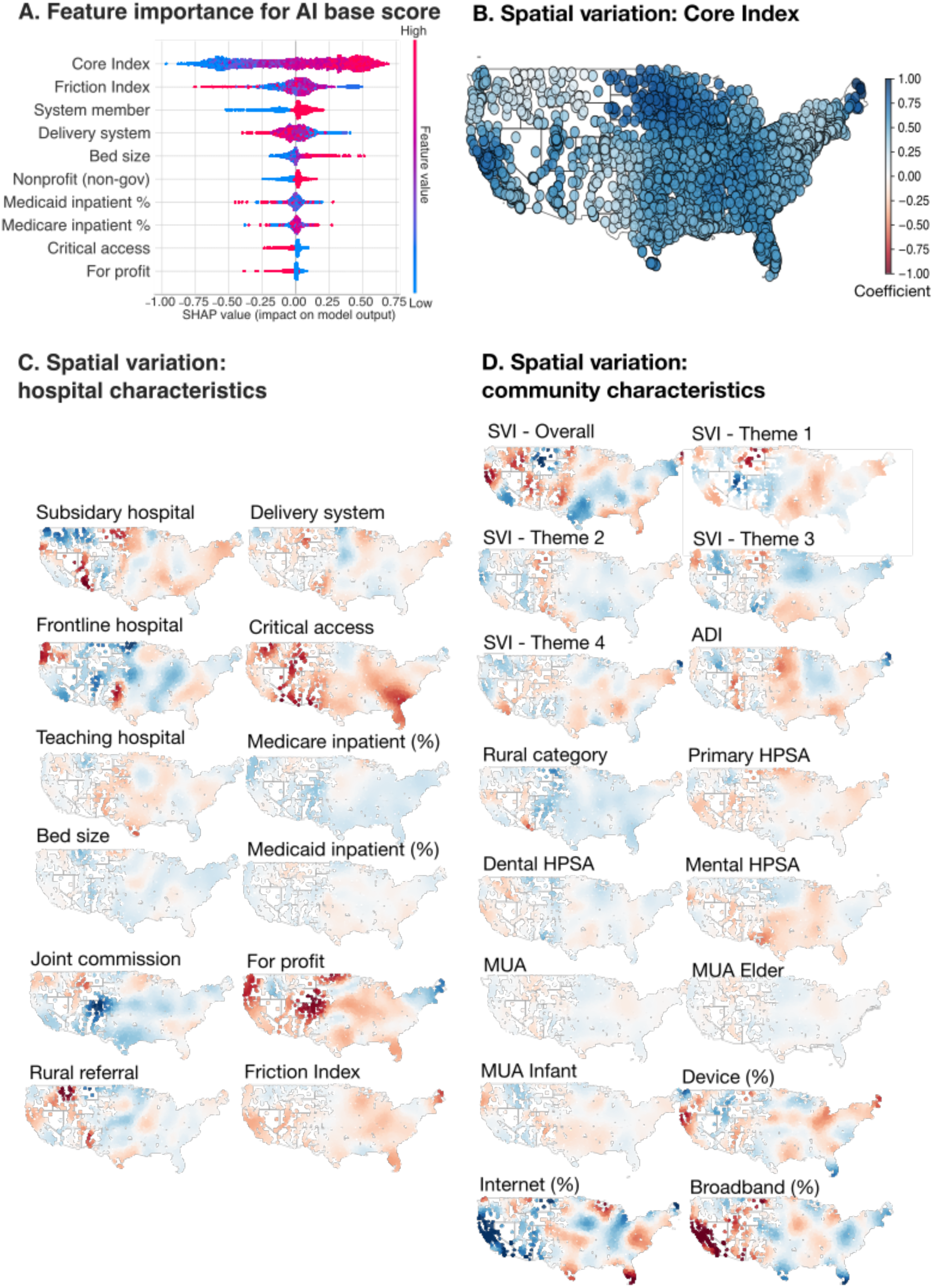
Feature Importance and Spatial Analysis of AI Implementation Factors AI, Artificial intelligence; HPSA, Health Professional Shortage Area; MUA, Medically Underserved Area; SVI, Social Vulnerability Index; ADI, Area Deprivation Index; SHAP, Shapley Additive Explanations; GWR, Geographically Weighted Regression. Panel A shows feature importance for AI base score using SHAP values, with Core Index and Friction Index demonstrating the highest predictive importance. Panel B displays the consistency of the Core Index across the US from GWR analysis. Panels C and D show regional variation across the US for hospital characteristics and community characteristics, respectively, from GWR analysis. Panel C presents geographic distribution of hospital characteristics including ownership type, teaching status, system membership, and patient population metrics. Panel D shows spatial patterns of community characteristics including socioeconomic disadvantage measures (SVI themes, ADI), healthcare access indicators (HPSA designations, MUA status), rurality, and technology infrastructure (internet and broadband access).

The five most important features were Core Index (measuring hospitals’ interoperability capabilities), Friction Index (quantifying barriers to health information exchange), system membership status, delivery system centralization model, and bed capacity. SHAP analysis demonstrated that hospitals with higher interoperability capabilities (Core Index), system membership, and larger bed capacity showed increased likelihood of AI adoption. Conversely, hospitals with greater barriers to information exchange (higher Friction Index) and more decentralized delivery systems were less likely to implement AI technologies.

### 3.5 Spatial Heterogeneity in Factors Influencing AI Implementation

Geographically weighted regression analysis revealed significant spatial heterogeneity in the relationships between hospital-level and community-level factors and AI implementation across the US (Figure 4b-d). Local regression coefficients varied by geographic region, indicating that the influence of predictor variables on AI adoption was not spatially uniform. The Core Index demonstrated the most spatial stability, maintaining positive coefficients throughout all regions. In contrast, other hospital-level factors exhibited pronounced spatial heterogeneity, with coefficients varying from negative to positive values. Community-level characteristics similarly demonstrated spatial variation in their associations with AI implementation.

### 3.6 Longitudinal Analysis of Hospital Quality Trends by AI Implementation Status

Our longitudinal analysis examined associations between AI implementation and changes in hospital quality metrics across 15 quarterly time points from 2022 to 2025. Linear mixed-effects models compared trend trajectories between hospitals without predictive models (AI-0, reference) and those with AI-based predictive models (AI-2). Of 17 quality metrics analyzed, 15 showed statistically significant trajectory differences after BH FDR correction (Table S19).

AI-adopting hospitals demonstrated favorable trends in several domains: reduced 30-day pneumonia mortality (β = −0.31 [−0.34, −0.27], t = −16.21, P < 0.0001), lower total hospital-acquired condition (HAC) score (β = −0.014 [−0.024, −0.004], t = −2.80, P = 0.006), fewer excess acute-care days after discharge for heart failure (β = −0.78 [−1.12, −0.45], t = −4.56, P < 0.0001) and pneumonia (β = −0.34 [−0.64, −0.03], t = −2.15, P = 0.036), and higher influenza vaccination coverage (β = 0.69 [0.51, 0.86], t = 7.70, P < 0.0001).

Less favorable trends were also observed: higher 30-day readmission rates for heart failure (β = 0.060 [0.040, 0.079], t = 6.03, P < 0.0001) and pneumonia (β = 0.0013 [0.0003, 0.0022], t = 2.54, P = 0.02), longer ED median times (β = 0.95 [0.64, 1.26], t = 6.00, P < 0.0001), higher rates of patients leaving ED without being seen (β = 0.060 [0.033, 0.086], t = 4.41, P < 0.0001), increased unsafe opioid prescribing (β = 0.92 [0.77, 1.06], t = 12.09, P < 0.0001), and worsened sepsis shock incidence rate (SEP-1: β = 0.90 [0.71, 1.10], t = 8.93, P < 0.0001, SEV-SEP-3HR: β = 0.35 [0.21, 0.49], t = −4.79, P < 0.0001). Residual diagnostics indicated that the two ED outcome models deviated from normality, while the others were approximately normal.

## 4. Discussion

This nationwide study of 3520 U.S. hospitals reveals the uneven landscape of healthcare AI implementation in 2023-2024 and characterizes geographic disparities with implications for health equity. AI implementation exhibits pronounced regional variation with clear spatial clustering. Hospitals in provider shortage areas or medically underserved areas consistently had lower AI implementation ratios, indicating that hospitals in high-need areas had lower rates of AI adoption compared to those in less underserved areas. Results show that the factors associated with AI implementation vary substantially across geographic space, challenging one-size-fits-all policy approaches and suggesting the need for regionally tailored strategies. The identification of specific hotspots, coldspots, and clusters provides geographic targets for stakeholders to address implementation gaps. Longitudinal analysis suggests potential associations between AI implementation and hospital quality metrics across clinical domains. Together, these findings provide system-level insights into AI integration patterns across hospital systems and identify disparities that can inform policy, implementation, and future research.

Our findings extend earlier work examining patterns of hospital AI adoption. Chen and Yan^4^ examined spatial autocorrelation in AI adoption and showed that hospitals in high-deprivation areas were less likely to implement these technologies, providing early evidence of geographic disparity. Poon et al^8^ conducted a survey of 43 nonprofit health systems to assess adoption of AI across multiple clinical use cases, perceived success, and barriers to implementation.

We build on these foundational studies in several important ways. First, while earlier work has focused on either selected health systems or on general geographic clustering, our analysis captures multiple dimensions of AI implementation, including evaluation, development, and breadth across a nationwide sample. Second, rather than simply describing the clustering patterns, we identify hotspots and coldspots, offering more actionable insights into where adoption is concentrated and where it lags. Third, our geographically weighted regression analysis shows that the factors associated with AI implementation vary substantially across geographic space, with interoperability emerging as the most consistent predictor. This spatial heterogeneity suggests the potential value of regionally tailored strategies that address local contexts and barriers rather than uniform implementation approaches.

Our findings reveal that AI adoption is especially limited in areas with healthcare acess needs. Provider shortage and medical underservice measures (HPSA and MUA) consistently showed low implementation ratios, with high-need regions far less likely to adopt AI-based predictive models. By comparison, socioeconomic community indicators, such as ADI and SVI, displayed more mixed patterns, with disparities less pronounced and in some cases approaching parity between low- and high-need regions. These differences underscore that healthcare capacity shortages and socioeconomic disadvantage capture distinct aspects of community need and may have different implications for AI adoption. Current deployment patterns show limited alignment with areas of highest healthcare need. However, this pattern may reflect realistic constraints of early-stage AI implementation, which requires substantial institutional resources and technical expertise that may be less available in underserved areas. Additionally, higher AI adoption does not necessarily indicate better outcomes, as implementation quality, clinical integration, and local appropriateness are critical factors not captured in our adoption measures. Future research should examine the complex relationships between hospital institutional characteristics, community needs, and AI implementation approaches. To illustrate this concept, Figure 5 demonstrates how hospital AI implementation status can be overlaid with community indicators such as area deprivation, revealing spatial patterns that aren’t apparent when examining either dataset alone. This type of integrated visualization could be expanded into comprehensive interactive tools that allow stakeholders to dynamically explore relationships between multiple hospital characteristics (size, ownership, teaching status, interoperability capabilities, resource capacity, quality metrics, financial performance, specialty services) and various community contexts (healthcare shortages, socioeconomic factors, digital infrastructure, population health outcomes). Such approaches could help researchers and policymakers better understand the multifaceted nature of AI adoption by examining institutional capacity alongside community contexts.

**Figure 5.**
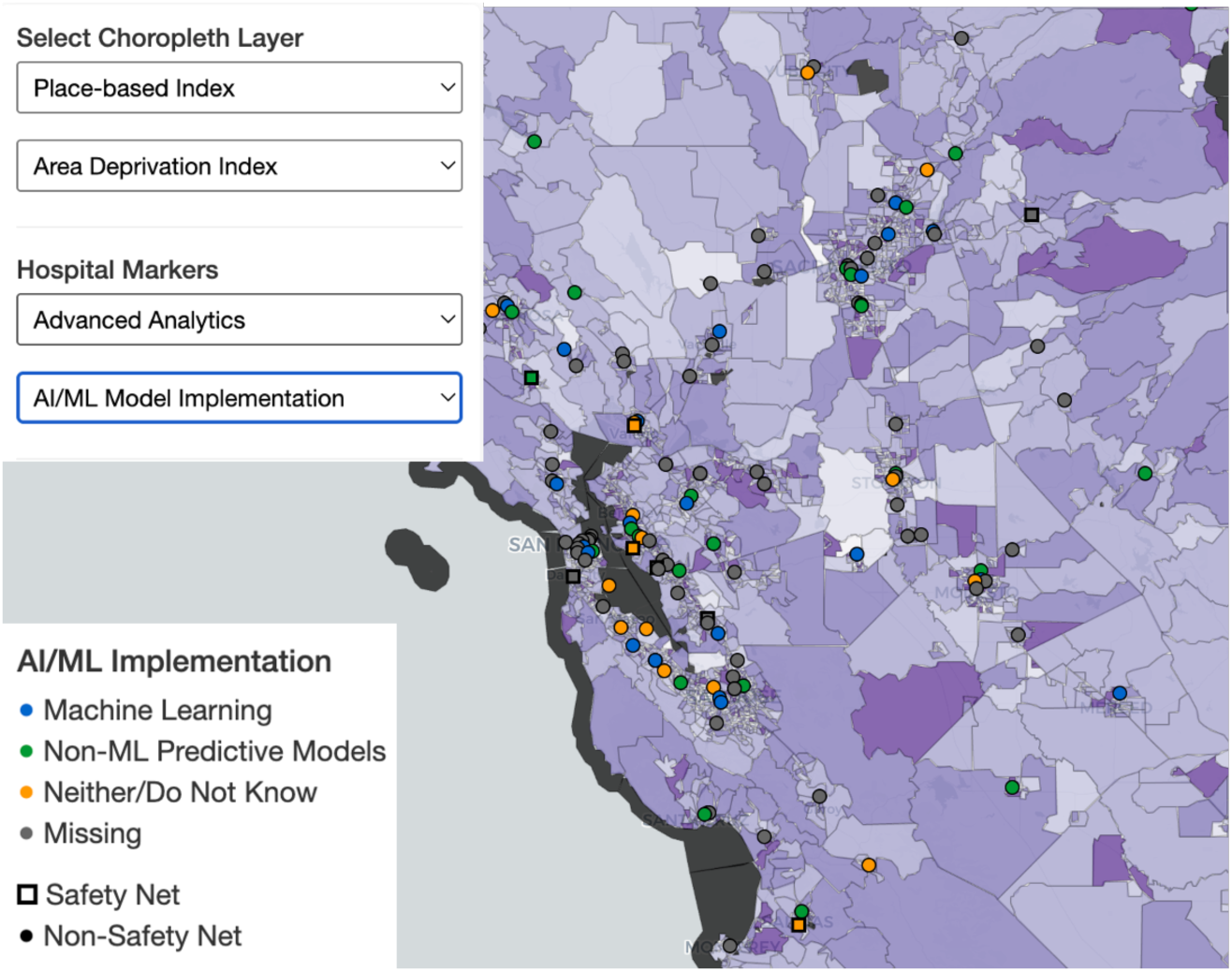
Conceptual Interactive Visualization of Hospital AI Implementation and Community Context Conceptual interactive map demonstrating how hospital AI implementation data could be integrated with community indicators to reveal spatial patterns not apparent when examining datasets independently. This mockup shows the San Francisco Bay Area using simulated data and does not represent real hospital or community characteristics. Hospitals are categorized by AI/ML implementation status: Machine Learning (blue circles), Non-ML Predictive Models (green circles), Neither/Do Not Know (orange circles), and Missing data (gray circles). Safety net status is indicated by square markers (Safety Net) versus circular markers (Non-Safety Net). The interface allows users to select different hospital characteristics informations from dropdown menus (shown: AI/ML Model Implementation) and toggle between community indices in the choropleth layer (shown: Area Deprivation Index) to examine relationships between various hospital attributes and community characteristics. This type of integrated visualization framework could be expanded into comprehensive interactive tools that allow stakeholders to dynamically explore relationships between multiple hospital characteristics and various community contexts, helping researchers and policymakers better understand the multifaceted nature of AI adoption by examining institutional capacity alongside community needs.

Our feature importance analysis showed that interoperability measures (Core Index and Friction Index) emerged as the strongest predictors of AI implementation. This finding suggests that foundational data exchange capabilities may serve as prerequisites for enabling advanced analytics implementation.^25–27^ The strong predictive performance (R²=0.61) was largely driven by these measures, with higher Core Index scores consistently associated with greater AI adoption across geographies. This spatial consistency highlights interoperability as a promising potential leverage point for addressing geographic disparities in AI adoption. However, this relationship may be confounded by underlying factors such as EHR vendor capabilities, as hospitals using market-leading platforms often have both high interoperability scores and integrated AI offerings. ^16,28^ Additionally, stronger interoperability may also reflect broader aspects of institutional readiness, including resource capacity and infrastructure, rather than serving as a direct driver of AI adoption.

Our longitudinal analysis across 15 quarterly time points from 2022 to 2025 examined associations between AI implementation and hospital performance, building on findings from our previous cross-sectional study examining county-level AI implementation and health outcomes.^11^ This study extends that work by examining hospital-level quality trajectories over time rather than single-time point county-level associations. These patterns may reflect institutional capacity differences rather than AI effects, requiring cautious interpretation given the limitations detailed in the methods and limitations sections. This exploratory analysis provides preliminary observations about potential relationships between AI adoption and hospital performance trajectories. After adjusting for outcome-specific hospital characteristics and regional factors, AI-adopting hospitals showed varied patterns across different clinical areas. Some measures suggested more favorable trajectories, including reduced mortality rates for pneumonia patients and decreased excess days in acute care after hospitalization. However, other measures showed less favorable patterns, including ED efficiency and sepsis care performance. These mixed associations underscore the complexity of early-stage AI implementation and the importance of avoiding assumptions about uniform benefits, particularly during a period when AI implementation was limited in scope and integration. A key limitation of our analysis is the lack of detailed variables linking specific AI applications to relevant quality metrics, highlighting the critical need for granular, standardized AI reporting that would enable proper assessment of implementation-outcome relationships. Future research with standardized AI implementation reporting and targeted hospital quality metrics will be essential for more definitive assessment of AI impacts on healthcare quality and patient outcomes.

There are important limitations that warrant consideration when interpreting our findings. First, our analysis is limited to hospitals that responded to the 2023 or 2024 AHA IT Supplement, which may introduce selection bias as responding hospitals may differ systematically from non-responding hospitals in their IT infrastructure, resources, or organizational characteristics. We presented descriptive statistics comparing between respondents and non-respondents to assess this potential bias (Table S1).

Second, AI implementation status was measured cross-sectionally from the 2023 or 2024 AHA IT Supplement, capturing only a single point in time. For our longitudinal quality analysis, we treated AI adoption as time-invariant due to lack of precise implementation timing, which may not reflect the true temporal relationship between adoption and quality changes. We combined responses from both survey years to maximize coverage, though this introduced minor variability from survey differences and response patterns. We present descriptive statistics by response pattern (2023 only, 2024 only, both years) to provide transparency about potential differences (Table S4).

Third, our quality metrics analysis was limited to variables with less than 50% missingness. While this threshold preserved sufficient sample size and statistical power, it may introduce selection bias and limits generalizability. The longitudinal analyses should therefore be interpreted with caution and viewed as exploratory.

Fourth, this study examined AI adoption at an early stage (2023–2024). Observed patterns may reflect initial experimentation rather than mature, integrated deployment. Future studies should reassess these relationships as adoption expands and evolves.

Fifth, despite integrating multiple datasets, unmeasured confounders likely influenced both adoption decisions and quality outcomes. In addition, our random forest and SHAP analyses assume feature independence, which may not fully hold given correlations among hospital, geographic, and community characteristics.

Sixth, our quality metric analysis could not account for specific AI use cases because the survey questions on model development and evaluation referred to predictive models in general rather than to individual AI models. This lack of specificity limits our ability to determine how particular AI applications influence targeted quality outcomes, and whether models were independently validated or externally tested. Adoption of AI does not necessarily imply better performance or improved clinical outcomes, and poorly validated models may even increase risks, particularly in high-need settings. Future research should integrate independent validation and real-world effectiveness data to clarify the impact of AI adoption on hospital quality and equity.

Seventh, our secondary implementation measures (e.g., breadth, development, evaluation scores) were exploratory and not validated constructs. Developing standardized and validated measures, as Strawley et al. did for interoperability, would require separate studies. Our aim was to provide flexible, continuous indicators for exploratory analysis, and future users may adapt parameters or scoring schemes for their own needs.

Eighth, although we distinguished between AI-based and non-AI predictive models, we could not assess their relative performance. Therefore, our analysis cannot determine whether AI model adoption translates into improved outcomes compared with traditional predictive models.”

Finally, while our findings identified significant associations, they cannot establish causal relationships between geographic factors and AI implementation or between AI adoption and quality outcomes. Addressing this gap will require longitudinal tracking of adoption, standardized assessments of implementation maturity, and analysis of specific AI use cases in relation to targeted outcomes.

## 5. Conclusion

This nationwide geospatial analysis provides critical baseline evidence of geographic disparities in hospital AI implementation during early adoption (2023-2024). Our analysis reveals significant spatial clustering in AI implementation patterns, with implementation ratios indicating lower adoption rates in areas with high healthcare access needs. We also observed mixed associations between AI adoption and hospital quality outcomes; however, our study lacked standardized measures to rigorously assess these relationships, underscoring the need for more detailed and validated metrics in future work. Geographically weighted regression demonstrated substantial regional variation in factors associated with AI implementation, with interoperability measures emerging as the most consistent predictor across geographies. These findings provide important insights for future research and policy development. AI implementation initiatives may benefit from regional approaches that consider local contexts and implementation readiness. AI implementation should be coupled with systematic evaluation and monitoring systems to assess quality, effectiveness, and potential impacts. Our approach provides a flexible framework for ongoing assessment of AI adoption as it matures and adapts as standardized metrics emerge. As AI implementation evolves beyond these early adoption patterns, continued assessment using adaptable approaches will be important for understanding how these technologies influence healthcare disparities and outcomes.

## Data Availability

Except for AHA data, all data are publicly available through the sources cited in the references. AHA data requires a subscription and can be obtained through AHA. Information on data access is available in the cited references.

## 6. Disclosures

### Author Contributions

Drs. Yeon-Mi Hwang and Madelena Y. Ng had full access to all the data in the study and take responsibility for the integrity of the data and the accuracy of the data analysis. *Concept and design: Hwang, Ng, Pillai, Hernandez-Boussard*.

*Acquisition, analysis, or interpretation of data: Hwang, Ng*.

*Drafting of the manuscript: Hwang*.

*Critical review of the manuscript for important intellectual content: All authors*.

*Statistical analysis: Hwang*.

*Obtained funding: Ng, Hernandez-Boussard*.

*Administrative, technical, or material support: Hernandez-Boussard*.

*Supervision: Hernandez-Boussard*.

### Human Ethics and Consent to Participate declarations

Not applicable.

### Data availability

This study used both publicly available and third-party proprietary datasets. The 2023 American Hospital Association (AHA) Annual Survey and 2023-2024 Information Technology (IT) Supplement were obtained from the AHA (https://www.ahadata.com/aha-data-resources) and are available for purchase under a data use agreement. These data are not publicly available and cannot be shared by the authors; interested researchers must obtain access from the AHA.

Other data sources are publicly available, including hospital quality metrics (2023-2025) from the Centers for Medicare & Medicaid Services (https://data.cms.gov/provider-data/archived-data/hospitals), the Area Deprivation Index from the University of Wisconsin Neighborhood Atlas (https://www.neighborhoodatlas.medicine.wisc.edu/), the Social Vulnerability Index from the Centers for Disease Control and Prevention (https://www.atsdr.cdc.gov/place-health/php/svi/svi-data-documentation-download.html), and Health Professional Shortage Area and Medically Underserved Area designations from the Health Resources and Services Administration (https://data.hrsa.gov/data/download). United States Census data were accessed via the U.S. Census Bureau’s public API.

Detailed information on data access and preprocessing is available in the project’s GitHub repository (https://github.com/su-boussard-lab/ai-disparities-us-hospitals).

### Code availability

Code used in this study is available at https://github.com/su-boussard-lab/ai-disparities-us-hospitals. The repository includes scripts for data collection, preprocessing, and analysis, reflecting the full workflow described in the manuscript. Access to proprietary datasets is required to fully execute the analysis.

