## Supplementary Materials for "AI Implementation in U.S. Hospitals: Regional Disparities and System-Level Implications"

**Supplementary Tables**

**Table S1.** Descriptive statistics by AHA IT Supplement response status

**Table S2.** Definitions and scoring criteria for AI implementation metrics

**Table S3.** Variable definitions

**Table S4.** Descriptive statistics by AHA IT Supplement response year

**Table S5.** Descriptive statistics by U.S. census division

**Table S6.** Descriptive statistics by U.S. state (available in Supplementary Data)

**Table S7.** Descriptive statistics by hospital AI implementation level

**Table S8.** Need indicator tertile distribution

**Table S9.** Moran’s I results by AI census division and state (available in Supplementary Data)

**Table S10.** DBSCAN clustering results by census division

**Table S11.** Hotspot/coldspot results across U.S. hospitals and hospital service areas

**Table S12.** Hotspot/coldspot results for AI implementation base score by census division and state

**Table S13.** Hotspot/coldspot results for model breadth score by census division and state

**Table S14.** Hotspot/coldspot results for model development score by census division and state

**Table S15.** Hotspot/coldspot results for model evaluation 2023 score by census division and state

**Table S16.** Hotspot/coldspot results for model evaluation 2024 score by census division and state

**Table S17.** Hotspot/coldspot results for LLM readiness score score by census division and state

**Table S18.** Hospital quality metrics and selected covariates

**Table S19.** Longitudinal analysis of AI implementation and quality outcomes

**Supplementary Figures**

**Figure S1.** Sensitivity analysis for parameter k-nearest-neighbor selection

**Figure S2.** DBSCAN parameter tuning and sensitivity analysis

**Figure S3.** Heatmap of AI implementation scores across hospitals

**Figure S4.** Hotspot/coldspot summary for base score by census division and state

**Figure S5.** Hotspot/coldspot summary for model breadth score by census division and state

**Figure S6.** Hotspot/coldspot summary for model development score by census division and state

**Figure S7.** Hotspot/coldspot summary for model evaluation score by census division and state

**Figure S8.** SHAP feature importance analysis for hospital AI implementation prediction

**Figure S9.** Geographic variation in hospital characteristics predicting hospital AI implementation

**Figure S10.** Geographic variation in community characteristics predicting hospital AI implementation

### **Supplementary Tables**

**Table S1.** Descriptive statistics by AHA IT Supplement response status.

Variable definitions are provided in Table S3. To prevent re-identification through cross-tabulation, we suppressed cells with counts below 20 along with adjacent cells as needed. Continuous variables are presented as means with standard deviations, while categorical variables are shown as frequencies with percentages. Statistical comparisons across groups were conducted using ANOVA for continuous variables and chi-square tests for categorical variables. ANOVA assumes normality within groups, homogeneity of variance, and independence of observations; chi-square tests assume independence and adequate expected cell frequencies. These assumptions were assumed to be adequately met for descriptive comparisons. All statistical tests were two-sided with significance set at p<0.05. Bonferroni correction was applied to correct for multiple comparisons.

| **AHA IT Supplement response status** | | | | | |
| --- | --- | --- | --- | --- | --- |
|  | **Missing** | **Overall** | **0** | **1** | **P-Value** |
| **n** |  | 6093 | 2533 | 3560 |  |
| **Geographic location** |  |  |  |  |  |
| **Census division, n (%)** | 0 |  |  |  | <0.0001* |
| East North Central |  | 904 (14.8) | 407 (16.1) | 497 (14.0) |  |
| East South Central |  | 488 (8.0) | 266 (10.5) | 222 (6.2) |  |
| Mid Atlantic |  | 527 (8.6) | 192 (7.6) | 335 (9.4) |  |
| Mountain |  | 554 (9.1) | 248 (9.8) | 306 (8.6) |  |
| New England |  | 244 (4.0) | 105 (4.1) | 139 (3.9) |  |
| Pacific |  | 639 (10.5) | 272 (10.7) | 367 (10.3) |  |
| South Atlantic |  | 929 (15.2) | 364 (14.4) | 565 (15.9) |  |
| West North Central |  | 766 (12.6) | 225 (8.9) | 541 (15.2) |  |
| West South Central |  | 1042 (17.1) | 454 (17.9) | 588 (16.5) |  |
| **Hospital characteristics** |  |  |  |  |  |
| **System member, n (%)** | 0 |  |  |  | <0.0001* |
| Yes |  | 4157 (68.2) | 1551 (61.2) | 2606 (73.2) |  |
| **Community hospital, n (%)** | 0 |  |  |  | <0.0001* |
| Yes |  | 5112 (83.9) | 1877 (74.1) | 3235 (90.9) |  |
| **Ownership, n (%)** | 0 |  |  |  | <0.0001* |
| Federal government |  | 207 (3.4) | 163 (6.4) | 44 (1.2) |  |
| For profit |  | 1586 (26.0) | 928 (36.6) | 658 (18.5) |  |
| Non-profit non-government |  | 3142 (51.6) | 933 (36.8) | 2209 (62.1) |  |
| Nonfederal government |  | 1158 (19.0) | 509 (20.1) | 649 (18.2) |  |
| **Delivery system, n (%)** | 0 |  |  |  | <0.0001* |
| Centralized Health System |  | 561 (9.2) | 73 (2.9) | 488 (13.7) |  |
| Centralized Physician/Insurance Health System |  | 257 (4.2) | 70 (2.8) | 187 (5.3) |  |
| Decentralized Health System |  | 801 (13.1) | 169 (6.7) | 632 (17.8) |  |
| Independent Hospital System |  | 1280 (21.0) | 665 (26.3) | 615 (17.3) |  |
| Moderately Centralized Health System |  | 1258 (20.6) | 574 (22.7) | 684 (19.2) |  |
| None |  | 1936 (31.8) | 982 (38.8) | 954 (26.8) |  |
| **Children hospital, n(%)** |  |  |  |  |  |
| Yes |  | 138 (2.3) | 63 (2.5) | 75 (2.1) | 0.37 |
| **Teaching hospital, n (%)** | 0 |  |  |  |  |
| Yes |  | 2659 (43.6) | 917 (36.2) | 1742 (48.9) | <0.0001* |
| **Major teaching hospital, n (%)** | 0 |  |  |  | <0.0001* |
| Yes |  | 281 (4.6) | 76 (3.0) | 205 (5.8) |  |
| **Minor teaching hospital, n (%)** | 0 |  |  |  | <0.0001* |
| Yes |  | 2378 (39.0) | 841 (33.2) | 1537 (43.2) |  |
| **Bed size, n (%)** | 0 |  |  |  | <0.0001* |
| 6-24 beds |  | 925 (15.2) | 445 (17.6) | 480 (13.5) |  |
| 25-49 beds |  | 1418 (23.3) | 657 (25.9) | 761 (21.4) |  |
| 50-99 beds |  | 1151 (18.9) | 516 (20.4) | 635 (17.8) |  |
| 100-199 beds |  | 1141 (18.7) | 470 (18.6) | 671 (18.8) |  |
| 200-299 beds |  | 576 (9.5) | 196 (7.7) | 380 (10.7) |  |
| 300-399 beds |  | 353 (5.8) | 108 (4.3) | 245 (6.9) |  |
| 400-499 beds |  | 187 (3.1) | 59 (2.3) | 128 (3.6) |  |
| 500 or more beds |  | 342 (5.6) | 82 (3.2) | 260 (7.3) |  |
| **Joint Commission accreditation, n (%)** | 0 |  |  |  | 0.0926 |
| Yes |  | 3965 (65.1) | 1617 (63.8) | 2348 (66.0) |  |
| **Critical access hospital, n (%)** | 0 |  |  |  | 0.0062* |
| Yes |  | 1345 (22.1) | 515 (20.3) | 830 (23.3) |  |
| **Rural referral center, n (%)** | 0 |  |  |  | <0.0001* |
| Yes |  | 726 (11.9) | 193 (7.6) | 533 (15.0) |  |
| **Operation of subsidiary hospital, n (%)** | 0 |  |  |  | <0.0001* |
| Yes |  | 584 (9.6) | 115 (4.5) | 469 (13.2) |  |
| **Frontline facility, n (%)** | 0 | 5642 (92.6) | 2452 (96.8) | 3190 (89.6) | <0.0001* |
| Yes |  | 451 (7.4) | 81 (3.2) | 370 (10.4) |  |
| **Healthcare payer mix** |  |  |  |  |  |
| **Medicare inpatient days percentage, mean (SD)** | 1 | 49.0 (23.6) | 45.4 (25.4) | 51.6 (22.0) | <0.0001* |
| **Medicaid inpatient days percentage, mean (SD)** | 1 | 18.3 (16.0) | 17.4 (16.3) | 19.0 (15.7) | <0.0001* |
| **Hospital Interoperability capability** |  |  |  |  |  |
| **Core index, mean (SD)** | 3001 | 3.5 (1.8) | nan (nan) | 3.5 (1.8) |  |
| **Friction index, mean (SD)** | 3001 | 0.8 (0.4) | nan (nan) | 0.8 (0.4) |  |
| **Geographic community characteristics** |  |  |  |  |  |
| **Hospital Service Area geographic socioeconomic context** |  |  |  |  |  |
| **Rural urban category, n (%)** | 0 |  |  |  |  |
| Metro |  | 4156 (68.2) | 1758 (69.4) | 2398 (67.4) | 0.1199 |
| Micro |  | 849 (13.9) | 327 (12.9) | 522 (14.7) |  |
| Rural |  | 1088 (17.9) | 448 (17.7) | 640 (18.0) |  |
| **National area deprivation index, mean (SD)** | 13 | 58.9 (24.6) | 60.4 (24.2) | 57.8 (24.9) | <0.0001* |
| **Overall Social Vulnerability Index, mean (SD)** | 12 | 0.6 (0.2) | 0.6 (0.2) | 0.5 (0.2) | <0.0001* |
| **Social Vulnerability Index Theme 1, mean (SD)** | 12 | 0.5 (0.2) | 0.5 (0.2) | 0.5 (0.2) | <0.0001* |
| **Social Vulnerability Index Theme 2, mean (SD)** | 7 | 0.5 (0.2) | 0.6 (0.2) | 0.5 (0.2) | <0.0001* |
| **Social Vulnerability Index Theme 3, mean (SD)** | 5 | 0.6 (0.2) | 0.6 (0.2) | 0.6 (0.2) | 0.0010* |
| **Social Vulnerability Index Theme 4, mean (SD)** | 7 | 0.5 (0.2) | 0.6 (0.2) | 0.5 (0.2) | 0.0001* |
| **County Digital Infrastructure** |  |  |  |  |  |
| **at least one device (%), mean (SD)** | 42 | 93.7 (3.4) | 93.5 (3.6) | 93.8 (3.2) | 0.0012* |
| **broadband connection (%), mean (SD)** | 42 | 87.8 (5.5) | 87.5 (5.9) | 88.0 (5.2) | 0.0010* |
| **internet connection (%), mean (SD)** | 42 | 90.6 (5.0) | 90.3 (5.3) | 90.8 (4.7) | 0.0008* |
| **Hospital Service Area Healthcare Access and provider shortage** |  |  |  |  |  |
| **Primary Health Professional Shortage Score, mean (SD)** | 0 | 8.6 (7.4) | 8.8 (7.5) | 8.5 (7.4) | 0.0694 |
| **Dental Health Professional Shortage Score, mean (SD)** | 0 | 8.1 (8.1) | 8.4 (8.2) | 7.9 (8.1) | 0.0183* |
| **Mental Health Health Professional Shortage Score, mean (SD)** | 0 | 11.8 (7.1) | 11.9 (7.2) | 11.8 (7.1) | 0.3753 |
| **Medical Underserve Area Score, mean (SD)** | 0 | 23.4 (27.7) | 24.6 (27.8) | 22.5 (27.7) | 0.0037* |
| **Elders Medical Underservice Area Score, mean (SD)** | 0 | 1.9 (5.2) | 2.0 (5.3) | 1.8 (5.1) | 0.4343 |
| **Infant Medical Underservice Area Score, mean (SD)** | 0 | 3.3 (8.5) | 3.4 (8.6) | 3.2 (8.4) | 0.4683 |

**Table S2.** Definitions and scoring criteria for AI and model implementation metrics

The AI implementation base score (0–2) is the primary measure and is reported in its original range to preserve interpretability. For visualization, it may be normalized to a 0-1 scale to align with secondary metrics. Secondary model implementation metrics include breadth, development approach, and evaluation scores. These secondary metrics were calculated by adding the base score to component scores derived from related survey items, scaled to a 0-1 range improve comparability across measures and reduce the risk of range-driven inflation. The LLM readiness score was calculated separately and does not include the base score. Point allocations for secondary measures were informed by published literature and author consensus, but they are judgment-based and not empirically validated. These measures are intended for exploratory and descriptive analysis to compare relative rankings across hospitals, rather than to interpret absolute score values as definitive measures of capability. Both the model implementation evaluation score (2024 version) and the LLM readiness score were only calculated for hospitals that responded to the 2024 AHA IT survey, as the required survey items were not available in the 2023 survey. Code is provided to implement the scoring and workflow, and users may adjust the schema as needed. We applied z-score standardization for geospatial analysis to ensure equal contribution of metrics with different native ranges, while using min-max scaling (0-1) for descriptive statistics to enhance interpretability.

| Score | Score calculation |
| --- | --- |
| AI implementation base score (Range 0-2) | **2023 version** Q25. Does your hospital use any machine learning or other predictive models that display output or recommendations (e.g., risk scores or clinical support) in your EHR or an App embedded in or launched by your EHR? (one selection only)   a. Machine Learning = 2 b. Other Non-Machine Learning Predictive Models (e.g. APACHE IV) = 1 c. Neither = 0 d. Do not know = 0 e. No response = 0   **2024 version** Q18 Does your hospital use any machine learning or other predictive models that display output or recommendations (e.g., risk scores or clinical support) in your EHR or an App embedded in or launched by your EHR?  Q18a. Machine Learning   a. Yes = 2 b. No = 0 c. Do not know = 0   Q18b Other non-machine learning predictive models (e.g. APACHE IV)   a. Yes = 1 b. No = 0 c. Do not know = 0 |
| Scoring plan explanation | Hospitals using machine learning–based predictive models receive the highest score, as these systems generally require more advanced data infrastructure, integration capability, and governance compared to non–machine learning predictive models. Non–machine learning tools (e.g., APACHE IV) still provide clinical decision support but typically offer less adaptability and complexity, so they receive a lower score  To maintain consistency with the 2023 survey format, the 2024 responses were processed as follows: if a respondent answered “Yes” to Q18a (machine learning), the score was based solely on Q18a. If the response to Q18a was “No,” “Do not know,” or left blank, Q18b (non–machine learning predictive models) was used to assign the score. |
| Model implementation breadth score (Range 0-4) | **= AI implementation base score + total use case score**  Total use case score  **2023 version Q26 & 2024 version Q19.** To which of the following uses has your hospital applied machine learning or other predictive models? (multiple selection)   a. Predicting health trajectories or risks for inpatients +0.25 b. Identify high risk outpatients to inform follow-up care +0.25 c. Monitor health +0.25 d. Recommend treatments +0.25 e. Simplify or automate billing procedures +0.25 f. Facilitate scheduling +0.25 g. Other (operational process optimization) +0.25 h. Other (clinical use cases) +0.25 i. None of the above (machine learning application) +0.25 j. Do not know (machine learning application) + 0 k. No response + 0 |
| Scoring plan explanation | This measure captures the breadth of model applications within a hospital. Respondents receive +0.25 points for each distinct use case reported, reflecting the diversity of model integration across clinical and operational functions. |
| Model implementation development score (Range 0-6.5) | **= AI implementation base score + total development score**  Total development score  **2023 version Q27 & 2024 version Q20.** Who developed the machine learning or other predictive models used at your hospitals? *(multiple selection)*   a. Our EHR developer + 1.5 b. A third-party developer + 1.0 c. Self-developed + 1.5 d. Public domain + 0.5 e. Do not know + 0 f. No response + 0 |
| Scoring plan explanation | EHR developer developed +1.5: Vendor-developed models have shown higher adoption rates than local or third-party tools. (1) They offer a high likelihood of seamless integration into existing workflows and sustained vendor support, with mature engineering and quality assurance from the platform side. Marwaha et al., (2) noted that products offered by EHR vendors are more likely to have a proven track record use allowing prospective adopter to assess performance and value through external case studies. EHR vendors may also provide configuration libraries that facilitate sharing and adaptation across health systems, enhancing integration efficiency and reducing local development burden.  Self-developed +1.5: Reflects strong in-house capability and governance. Nong et al., 2025(1) reported more frequent local post-deployment evaluation among self-developed models.  Third party developer +1: Brings specialized external expertise, but integration can be more challenging than with EHR vendor models. Transparency may be limited if algorithms are proprietary and local customization is often needed. These models are less likely to undergo local evaluation.(1)  Public domain 0.5: Offers transparency and flexibility, but places the burden of integration, validation, and ongoing maintenance on the local team. While it removes the need for initial model creation, the lack of dedicated vendor or developer support often limits implementation depth and scalability.(3,4)  EHR vendor (1.5) and self developed (1.5) get the highest weight as they tend to be better integrated and/or better governed.  Third party (1.0) is lower because of integration friction and less local evaluation.  Public domain (0.5) is lowest, reflecting ease of acquisition but high local validation burden. |
| Model implementation evaluation score 2023 version (Range 0-4) | **= AI implementation base score + total 2023 evaluation score**  Total 2023 evaluation score  **2023 version Q28 & 2024 version Q21.** What share of your machine learning or other predictive models have been evaluated using data from your hospital or health system for? (one selection)   a. Model accuracy 1) All models + 1 2) Most models + 0.75 3) Some models + 0.5 4) Few models + 0.25 5) None + 0 6) Do not know + 0 7) No response + 0 b. Model bias 1) All models + 1 2) Most models + 0.75 3) Some models + 0.5 4) Few models + 0.25 5) None + 0 6) Do not know + 0 7) No response + 0 |
| Scoring plan explanation | Evaluating models locally for both accuracy and bias is essential to ensure safe and equitable deployment in the intended patient population. Higher points are given when a larger share of models undergo these evaluations, reflecting stronger governance and quality assurance. |
| Model implementation evaluation score 2024 version (Range 0-6.25) | **= AI implementation base score + total 2024 evaluation score**  Total 2024 evaluation score  **2023 version Q28 & 2024 version Q21.** What share of your machine learning or other predictive models have been evaluated using data from your hospital or health system for? (one selection)   a. Model accuracy 1) All models + 1 2) Most models + 0.75 3) Some models + 0.5 4) Few models + 0.25 5) None + 0 6) Do not know + 0 7) No response + 0 b. Model bias 1) All models + 1 2) Most models + 0.75 3) Some models + 0.5 4) Few models + 0.25 5) None + 0 6) Do not know + 0 7) No response + 0 c. Post-implementation evaluation or monitoring 1) All models + 1 2) Most models + 0.75 3) Some models + 0.5 4) Few models + 0.25 5) None + 0 6) Do not know + 0 7) No response + 0   **2024 version Q22** Who in your hospital or health care system is accountable for evaluating models? (Multiple selection)   a. Designated Senior Executive (CMIO/CIO/Director of Technology) + 0.25 b. Specific Committee or Task Force for Machine Learning or Predictive Modeling + 0.25 c. Clinical Decision Support Committee/other non-Machine Learning or Predictive Modeling specific Committee + 0.25 d. Division/Department Leaders + 0.25 e. IT staff + 0.25 f. None of the above (evaluating models) |
| Scoring plan explanation | The 2024 version expands the evaluation score to include three practices: model accuracy evaluation, bias evaluation, and post-implementation monitoring, each scored the same way as in the 2023 version. These measures capture whether models are assessed for performance, equity, and ongoing reliability in the local population.  The 2024 survey also added a governance question. Accountability structures, such as executive oversight, ML committees, and departmental leadership, help ensure consistent evaluation and timely issue detection. Hospitals receive +0.25 points for each governance mechanism reported, reflecting the breadth of oversight without allowing governance to outweigh actual evaluation practices. No points are given if none are reported.  The 2023 version of the score was calculated for both 2023 and 2024 responding hospitals to allow cross-year comparison. The 2024 version could only be calculated for hospitals responding to the 2024 survey, as the additional items were not included in the 2023 survey. |
| LLM readiness score | **2024 Q23** Does your hospital or health system have a large language model (e.g. ChatGPT, GPT-4, Google Gemini, Nuance DAX Copilot) integrated into the electronic health record? (Single selection)   a. No + 0 b. Yes, currently using + 3 c. Plan to use in the next year +2 d. Plan to use in the next 5 years +1 e. Do not know +0 |
| Scoring plan explanation | Hospitals currently using LLMs receive the highest score, followed by those planning near-term adoption, then longer-term adoption. Hospitals not using or planning LLMs receive zero points. This ranking reflects assumed readiness based on proximity to implementation. |

**Table S3.** Variable definitions

For categorical variables, we used {} to list categories. For continuous variables, we indicated the possible range using [].

| Variable | Definition |
| --- | --- |
| Hospital AI and model implementation/readiness |  |
| AI base implementation score | Hospital AI implementation level {0: No/No Response, 1: Non-AI Predictive Model, 2: AI predictive Model} |
| Model implementation breadth score | Hospital model use case breadth level [0,1] |
| Model implementation development score | Hospital model development level [0,1] |
| Model implementation evaluation score 2023 version | Hospital model evaluation level [0,1] |
| LLM readiness score | Hospital LLM readiness level [0,1] |
| Hospital characteristics |  |
| System Member | Hospital belonging to a corporate body that owns and/or manages health provider facilities or health-related subsidiaries. {0:No/No response,1:Yes} |
| Community hospital | All nonfederal, short-term general, and special hospitals whose facilities and services are available to the public {0:No/No response, 1:Yes} |
| Ownership type | Control code. Type of authority responsible for establishing policy concerning overall operation of the hospital {federal government, nonfederal government, not-for-profit nongovernment, for-profit} |
| Delivery system | Groups of health systems that share common strategic/structural features. Health system is assigned to one of five categories based on how much they differentiate and centralize their hospital service, physician arrangements, and provider-based insurance products. Hospitals with insufficient data were not available to determine a cluster assignment {1: Centralized health system, 2: Centralized physician/insurance health system, 3: Moderately centralized health system, 4: Decentralized health system, 5: Independent Hospital System} |
| Children hospital | Hospital with primary service designated to children {0: No/No response, 1: Yes} |
| Teaching hospital | Includes both major and minor teaching hospitals. Major teaching hospitals are defined as all hospitals with Council of Teaching Hospitals designation. Minor teaching hospitals are defined as hospitals that either serve as participating sites for one or more Accreditation Council for Graduate Medical Education accredited programs or report medical school affiliation to the American Medical Association.{0: No/No response, 1: Yes} |
| Joint commission accreditation | Accreditation by The Joint Commission {0: No/No response, 1: Yes} |
| Critical access hospital | Critical access hospital designation {0: No/No response, 1: Yes} |
| Rural referral center | Rural referral center designation {0:No/No response, 1: Yes} |
| Operation of subsidiary hospital | Hospital operates subsidiary operation {0:No/No response, 1: Yes} |
| Frontline facility | Frontline facility {0:No/No response, 1:Yes} |
| Bed size | Number of hospital beds set-up and staffed for use {1: 6-24 beds, 2: 25-49 beds, 3: 50-99 beds, 4: 100-199 beds, 5: 200-299 beds, 6: 300-399 beds, 7: 400-499 beds, 8: 500 or more beds} |
| Medicare inpatient days percentage | Proportion of inpatient days where a Medicaid Managed Care Plan is the source of payment |
| Medicaid inpatient days percentage | Proportion of inpatient days where a Medicaid Managed Care Plan is the source of payment |
| Core index | Index developed by Strawley et al., 2025(5) that measures adoption of foundational interoperability capabilities. Original methodology scales to 0-100; current analysis uses raw scores [0-7.125], where 0 = no interoperability and 7.125 = full interoperability). These scores were calculated based on 2023 responses. |
| Friction index | Index developed by Strawley et al., 2025(5) that measures barriers to interoperability. Original methodology scales to 0-100; current analysis uses raw scores [0-2.74], where 0 = no barriers and 2.74 = maximum barriers to interoperability. These scores were calculated based on 2023 responses. |
| Geographic community characteristics |  |
| Rural urban category | Hospitals are categorized based on the core-based statistical area type {1: rural, 2: micropolitan, 3: metropolitan} |
| Area deprivation index | Place-based index that ranks neighborhoods by adverse social exposome in a region of interest considering factors related to income, education, employment, and housing quality. [1-100], where 1= least disadvantaged and 100=most disadvantaged |
| Overall social vulnerability index | Place-based index developed by CDC that identifies and quantifies communities experiencing social vulnerability [0-1], where 0=area with lowest vulnerability and 1= area with highest vulnerability. |
| Social vulnerability index theme 1 | Social vulnerability index focuses on socioeconomic status (poverty, employment, income, educational attainment) [0-1], where 0=area with lowest vulnerability and 1= area with highest vulnerability. |
| Social vulnerability index theme 2 | Social vulnerability index focuses on household composition and disability (age distribution, disability, family structure) [0-1], where 0=area with lowest vulnerability and 1= area with highest vulnerability. |
| Social vulnerability index theme 3 | Social vulnerability index focuses on minority status and language (racial and ethnic minority status and English language proficiency) [0-1], where 0=area with lowest vulnerability and 1= area with highest vulnerability. |
| Social vulnerability index theme 4 | Social vulnerability index focuses on housing type and transportation (housing, housing density, and access to transportation) [0-1], where 0=area with lowest vulnerability and 1= area with highest vulnerability. |
| County digital infrastructure |  |
| Device access | Percentage of households with at least one computer or device in county [0-100] |
| Broadband connection | Percentage of households with high-speed internet [0-100] |
| Internet connection | Percentage of households with any internet access [0-100] |
| Hospital service area healthcare access and provider shortage |  |
| Primary health professional shortage score | Numerical rating assigned to designated areas to indicate the severity of primary care provider shortage [0-25], where 0 = no HPSA designation and 25 = most severe shortage). Hospitals in hospital service areas without HPSA designation were assigned a score of 0. |
| Dental health professional shortage score | Numerical rating assigned to designated areas to indicate the severity of dental care provider shortage [0-26], where 0 = no HPSA designation and 26 = most severe shortage). Hospitals in hospital service areas without HPSA designation were assigned a score of 0. |
| Mental health professional shortage score | Numerical rating assigned to designated areas to indicate the severity of mental care provider shortage [0-25], where 0 = no HPSA designation and 25 = most severe shortage). Hospitals in hospital service areas without HPSA designation were assigned a score of 0. |
| Medical Underserved Area Score | Numerical rating assigned to designated area to indicate the medical underservice [0-100]. |
| Elders medical underservice area score | Index of Medical Underservice (IMU) weighted value corresponding to the percentage of population in the service area which is age 65 or older [0,20.2] |
| Infant medical underservice area score | Infant mortality rate index of medical underservice score [0,26] |

**Table S4.** Descriptive statistics by AHA IT Supplement response year

Variable definitions are provided in Table S3. To prevent re-identification through cross-tabulation, we suppressed cells with counts below 20 along with adjacent cells as needed. Continuous variables are presented as means with standard deviations, while categorical variables are shown as frequencies with percentages. Statistical comparisons across groups were conducted using ANOVA for continuous variables and chi-square tests for categorical variables. ANOVA assumes normality within groups, homogeneity of variance, and independence of observations; chi-square tests assume independence and adequate expected cell frequencies. These assumptions were assumed to be adequately met for descriptive comparisons. All statistical tests were two-sided with significance set at P<0.05. Bonferroni correction was applied to correct for multiple comparisons.

|  | **Missing** | **Overall** | **2023** **only** | **2024** **only** | **both** | **P-Value** |
| --- | --- | --- | --- | --- | --- | --- |
| **n** |  | 3560 | 937 | 468 | 2155 |  |
| **AI implementation level** |  |  |  |  |  |  |
| **AI implementation score, mean (SD)** | 211 | 1.2 (0.9) | 1.0 (0.9) | 1.2 (1.0) | 1.3 (0.9) | <0.0001* |
| **AI implementation category, n (%)** |  |  |  |  |  |  |
| AI/ML model |  | 1738 (48.8) | 334 (35.6) | 225 (48.1) | 1179 (54.7) | <0.0001* |
| non-AI/ML predictive model |  | 568 (16.0) | 178 (19.0) | 26 (5.6) | 364 (16.9) |  |
| Do not know/Neither |  | 1043 (29.3) | 307 (32.8) | 159 (34.0) | 577 (26.8) |  |
| No response |  | 211 (5.9) | 118 (12.6) | 58 (12.4) | 35 (1.6) |  |
| **Secondary Model Implementation Measures** |  |  |  |  |  |  |
| **Model breadth implementation standardized, mean (SD)** | 213 | 0.5 (0.4) | 0.4 (0.3) | 0.4 (0.4) | 0.5 (0.3) | <0.0001* |
| **Model development score standardized, mean (SD)** | 211 | 0.5 (0.3) | 0.4 (0.3) | 0.4 (0.4) | 0.5 (0.3) | <0.0001* |
| **Model evaluation score 2023 version standardized, mean (SD)** | 211 | 0.5 (0.4) | 0.4 (0.4) | 0.5 (0.4) | 0.6 (0.4) | <0.0001* |
| **Model evaluation score 2024 version standardized, mean (SD)^a^** | 1030 | 0.5 (0.4) | nan (nan) | 0.4 (0.4) | 0.5 (0.4) |  |
| **LLM readiness score** |  |  |  |  |  |  |
| **LLM readiness score standardized, mean (SD)^a^** | 1327 | 0.6 (0.4) | nan (nan) | 0.4 (0.4) | 0.6 (0.4) |  |
| **Predictive Model Use Case** |  |  |  |  |  |  |
| **Predicting health trajectories or risks for inpatients, n (%)** |  |  |  |  |  |  |
| Yes |  | 2178 (61.2) | 482 (51.4) | 205 (43.8) | 1491 (69.2) | <0.0001* |
| **Identify high risk outpatients to inform follow-up care, n (%)** |  |  |  |  |  |  |
| Yes |  | 2093 (58.8) | 471 (50.3) | 200 (42.7) | 1422 (66.0) | <0.0001* |
| **Monitor health, n (%)** |  |  |  |  |  |  |
| Yes |  | 854 (24.0) | 213 (22.7) | 93 (19.9) | 548 (25.4) | <0.0001* |
| **Recommend treatments, n (%)** |  |  |  |  |  |  |
| Yes |  | 1083 (30.4) | 197 (21.0) | 137 (29.3) | 749 (34.8) | <0.0001* |
| **Simplify or automate billing procedures, n (%)** |  |  |  |  |  |  |
| Yes |  | 1314 (36.9) | 187 (20.0) | 147 (31.4) | 980 (45.5) | <0.0001* |
| **Facilitate scheduling, n (%)** |  |  |  |  |  |  |
| Yes |  | 1447 (40.6) | 196 (20.9) | 201 (42.9) | 1050 (48.7) | <0.0001* |
| **Other operational process optimization, n (%)** |  |  |  |  |  |  |
| Yes |  | 540 (15.2) | 77 (8.2) | 28 (6.0) | 435 (20.2) | <0.0001* |
| **Other clinical use case, n (%)** |  |  |  |  |  |  |
| Yes |  | 584 (16.4) | 71 (7.6) | 38 (8.1) | 475 (22.0) | <0.0001* |
| **Model Developer** |  |  |  |  |  |  |
| **EHR developer, n (%)** |  |  |  |  |  |  |
| Yes |  | 2024 (56.9) | 470 (50.2) | 226 (48.3) | 1328 (61.6) | <0.0001* |
| **Third-party developer, n (%)** |  | 1245 (35.0) | 248 (26.5) | 110 (23.5) | 887 (41.2) | <0.0001* |
| Yes |  | 1245 (35.0) | 248 (26.5) | 110 (23.5) | 887 (41.2) | <0.0001* |
| **Self-development, n (%)** |  |  |  |  |  |  |
| Yes |  | 1192 (33.5) | 191 (20.4) | 126 (26.9) | 875 (40.6) | <0.0001* |
| **Public domain, n (%)** |  |  |  |  |  |  |
| Yes |  | 89 (2.5) | 21 (2.2) | 2 (0.4) | 66 (3.1) | <0.0001* |
| **Model Evaluation** |  |  |  |  |  |  |
| **Model accuracy, n (%)** |  |  |  |  |  |  |
| All models |  | 1039 (29.2) | 235 (25.1) | 121 (25.9) | 683 (31.7) | <0.0001* |
| Most models |  | 563 (15.8) | 94 (10.0) | 30 (6.4) | 439 (20.4) |  |
| Some models |  | 152 (4.3) | 28 (3.0) | 9 (1.9) | 115 (5.3) |  |
| Few models |  | 108 (3.0) | 8 (0.9) | 25 (5.3) | 75 (3.5) |  |
| Do not know/None |  | 991 (27.8) | 96 (10.2) | 208 (44.4) | 687 (31.9) |  |
| No response |  | 707 (19.9) | 476 (50.8) | 75 (16.0) | 156 (7.2) |  |
| **Model bias, n (%)** |  |  |  |  |  |  |
| All models |  | 786 (22.1) | 197 (21.0) | 111 (23.7) | 478 (22.2) | <0.0001* |
| Most models |  | 495 (13.9) | 58 (6.2) | 29 (6.2) | 408 (18.9) |  |
| Some models |  | 299 (8.4) | 90 (9.6) | 15 (3.2) | 194 (9.0) |  |
| Few models |  | 151 (4.2) | 27 (2.9) | 19 (4.1) | 105 (4.9) |  |
| Do not know/None |  | 1159 (32.6) | 136 (14.5) | 212 (45.3) | 811 (37.6) |  |
| No response |  | 670 (18.8) | 429 (45.8) | 82 (17.5) | 159 (7.4) |  |
| **Post-implementation/monitoring, n (%)^a^** |  |  |  |  |  |  |
| All models |  | 643 (18.1) | 0 (0.0) | 120 (25.6) | 523 (24.3) | <0.0001* |
| Most models |  | 366 (10.3) | 0 (0.0) | 25 (5.3) | 341 (15.8) |  |
| Some models |  | 195 (5.5) | 0 (0.0) | 12 (2.6) | 183 (8.5) |  |
| Few models |  | 182 (5.1) | 0 (0.0) | 26 (5.6) | 156 (7.2) |  |
| Do not know/None |  | 896 (25.2) | 0 (0.0) | 207 (44.2) | 689 (32.0) |  |
| No response |  | 1278 (35.9) | 937 (100.0) | 78 (16.7) | 263 (12.2) |  |
| **Model evaluation responsible personnel^a^** |  |  |  |  |  |  |
| ***Designated Senior Executive (CMIO/CIO/Director of Technology), n (%)*** |  |  |  |  |  |  |
| Yes |  | 1144 (32.1) | 0 (0.0) | 194 (41.5) | 950 (44.1) | <0.0001* |
| **Specific Committee or Task Force for Machine Learning or Predictive Modeling, n(%)** |  |  |  |  |  |  |
| Yes |  | 1210 (34.0) | 0 (0.0) | 159 (34.0) | 1051 (48.8) | <0.0001* |
| **Clinical Decision Support Committee/other non-Machine Learning or Predictive Modeling specific Committee, n(%)** |  |  |  |  |  |  |
| Yes |  | 1062 (29.8) | 0 (0.0) | 175 (37.4) | 887 (41.2) | <0.0001* |
| **Division/Department Leaders, n(%)** |  |  |  |  |  |  |
| Yes |  | 1118 (31.4) | 0 (0.0) | 173 (37.0) | 945 (43.9) | <0.0001* |
| **IT staff, n(%)** |  |  |  |  |  |  |
| Yes |  | 885 (24.9) | 0 (0.0) | 160 (34.2) | 725 (33.6) | <0.0001* |
| **LLM integration in EHR** |  |  |  |  |  |  |
| **LLM integration in EHR** |  | 813 (22.8) | 0 (0.0) | 116 (24.8) | 697 (32.3) | <0.0001* |
| Yes, currently using |  | 585 (16.4) | 0 (0.0) | 47 (10.0) | 538 (25.0) |  |
| Plan to use in the next year |  | 149 (4.2) | 0 (0.0) | 28 (6.0) | 121 (5.6) |  |
| Plan to use in the next 5 years |  | 686 (19.3) | 0 (0.0) | 180 (38.5) | 506 (23.5) |  |
| Do not know/No |  | 1197 (33.6) | 937 (100.0) | 58 (12.4) | 202 (9.4) |  |
| None |  | 130 (3.7) | 0 (0.0) | 39 (8.3) | 91 (4.2) |  |
| **Geographic location** |  |  |  |  |  |  |
| **Census division, n (%)** |  | 497 (14.0) | 112 (12.0) | 68 (14.5) | 317 (14.7) | <0.0001* |
| East North Central |  | 222 (6.2) | 64 (6.8) | 30 (6.4) | 128 (5.9) |  |
| East South Central |  | 335 (9.4) | 93 (9.9) | 25 (5.3) | 217 (10.1) |  |
| Mid Atlantic |  | 306 (8.6) | 103 (11.0) | 30 (6.4) | 173 (8.0) |  |
| Mountain |  | 139 (3.9) | 50 (5.3) | 9 (1.9) | 80 (3.7) |  |
| New England |  | 367 (10.3) | 114 (12.2) | 62 (13.2) | 191 (8.9) |  |
| Pacific |  | 565 (15.9) | 150 (16.0) | 68 (14.5) | 347 (16.1) |  |
| South Atlantic |  | 541 (15.2) | 91 (9.7) | 47 (10.0) | 403 (18.7) |  |
| West North Central |  | 588 (16.5) | 160 (17.1) | 129 (27.6) | 299 (13.9) |  |
| **Hospital characteristics** |  |  |  |  |  |  |
| **System member, n (%)** |  |  |  |  |  |  |
| Yes |  | 2606 (73.2) | 649 (69.3) | 330 (70.5) | 1627 (75.5) | 0.0006* |
| **Community hospital, n (%)** |  |  |  |  |  |  |
| Yes |  | 3235 (90.9) | 830 (88.6) | 407 (87.0) | 1998 (92.7) | <0.0001* |
| **Ownership, n (%)** |  |  |  |  |  |  |
| Federal government |  | 44 (1.2) | 19 (2.0) | 8 (1.7) | 17 (0.8) | <0.0001* |
| For profit |  | 658 (18.5) | 271 (28.9) | 105 (22.4) | 282 (13.1) |  |
| Non-profit non-government |  | 2209 (62.1) | 486 (51.9) | 272 (58.1) | 1451 (67.3) |  |
| Nonfederal government |  | 649 (18.2) | 161 (17.2) | 83 (17.7) | 405 (18.8) |  |
| **Delivery system, n (%)** |  |  |  |  |  |  |
| Centralized Health System |  | 488 (13.7) | 96 (10.2) | 15 (3.2) | 377 (17.5) | <0.0001* |
| Centralized Physician/Insurance Health System |  | 187 (5.3) | 33 (3.5) | 11 (2.4) | 143 (6.6) |  |
| Decentralized Health System |  | 632 (17.8) | 84 (9.0) | 115 (24.6) | 433 (20.1) |  |
| Independent Hospital System |  | 615 (17.3) | 285 (30.4) | 81 (17.3) | 249 (11.6) |  |
| Moderately Centralized Health System |  | 684 (19.2) | 151 (16.1) | 108 (23.1) | 425 (19.7) |  |
| None |  | 954 (26.8) | 288 (30.7) | 138 (29.5) | 528 (24.5) |  |
| **Children hospital, n(%)** |  |  |  |  |  |  |
| Yes |  | 75 (2.1) | NA | NA | 46 (2.1) | 0.98 |
| **Teaching hospital, n (%)** |  |  |  |  |  |  |
| Yes |  | 1742 (48.9) | 391 (41.7) | 195 (41.7) | 1156 (53.6) | <0.0001* |
| **Major teaching hospital, n (%)** |  |  |  |  |  |  |
| Yes |  | 205 (5.8) | 35 (3.7) | 11 (2.4) | 159 (7.4) | <0.0001* |
| **Minor teaching hospital, n (%)** |  |  |  |  |  |  |
| Yes |  | 1537 (43.2) | 356 (38.0) | 184 (39.3) | 997 (46.3) | <0.0001* |
| **Bed size, n (%)** |  |  |  |  |  |  |
| 6-24 beds |  | 480 (13.5) | 123 (13.1) | 78 (16.7) | 279 (12.9) | <0.0001* |
| 25-49 beds |  | 761 (21.4) | 219 (23.4) | 118 (25.2) | 424 (19.7) |  |
| 50-99 beds |  | 635 (17.8) | 232 (24.8) | 88 (18.8) | 315 (14.6) |  |
| 100-199 beds |  | 671 (18.8) | 171 (18.2) | 69 (14.7) | 431 (20.0) |  |
| 200-299 beds |  | 380 (10.7) | 74 (7.9) | 55 (11.8) | 251 (11.6) |  |
| 300-399 beds |  | 245 (6.9) | 46 (4.9) | 24 (5.1) | 175 (8.1) |  |
| 400-499 beds |  | 128 (3.6) | 23 (2.5) | 17 (3.6) | 88 (4.1) |  |
| 500 or more beds |  | 260 (7.3) | 49 (5.2) | 19 (4.1) | 192 (8.9) |  |
| **Joint Commission accreditation, n (%)** |  |  |  |  |  |  |
| Yes |  | 2348 (66.0) | 631 (67.3) | 284 (60.7) | 1433 (66.5) | 0.0321* |
| **Critical access hospital, n (%)** |  |  |  |  |  |  |
| Yes |  | 830 (23.3) | 196 (20.9) | 108 (23.1) | 526 (24.4) | 0.1071 |
| **Rural referral center, n (%)** |  |  |  |  |  |  |
| Yes |  | 533 (15.0) | 103 (11.0) | 61 (13.0) | 369 (17.1) | <0.0001* |
| **Operation of subsidiary hospital, n (%)** |  |  |  |  |  |  |
| Yes |  | 469 (13.2) | 74 (7.9) | 43 (9.2) | 352 (16.3) | <0.0001* |
| **Frontline facility, n (%)** |  |  |  |  |  |  |
| Yes |  | 370 (10.4) | 76 (8.1) | 31 (6.6) | 263 (12.2) | <0.0001* |
| **Healthcare payer mix** |  |  |  |  |  |  |
| **Medicare inpatient days percentage, mean (SD)** | 1 | 51.6 (22.0) | 51.8 (22.7) | 50.6 (22.9) | 51.7 (21.4) | 0.5442 |
| **Medicaid inpatient days percentage, mean (SD)** | 1 | 19.0 (15.7) | 18.2 (15.4) | 18.3 (15.1) | 19.6 (15.9) | 0.0514 |
| **Hospital Interoperability capability** |  |  |  |  |  |  |
| **Core index, mean (SD)** | 468 | 3.5 (1.8) | 3.1 (1.9) | nan (nan) | 3.7 (1.7) |  |
| **Friction index, mean (SD)** | 468 | 0.8 (0.4) | 0.7 (0.5) | nan (nan) | 0.9 (0.4) |  |
| **Geographic community characteristics** |  |  |  |  |  |  |
| **Hospital Service Area geographic socioeconomic context** |  |  |  |  |  |  |
| **Rural urban category, n (%)** |  |  |  |  |  |  |
| Metro |  | 2398 (67.4) | 652 (69.6) | 309 (66.0) | 1437 (66.7) | 0.1795 |
| Micro |  | 522 (14.7) | 130 (13.9) | 61 (13.0) | 331 (15.4) |  |
| Rural |  | 640 (18.0) | 155 (16.5) | 98 (20.9) | 387 (18.0) |  |
| **National area deprivation index, mean (SD)** | 4 | 57.8 (24.9) | 56.6 (25.0) | 60.3 (24.6) | 57.8 (24.8) | 0.0331* |
| **Overall Social Vulnerability Index, mean (SD)** | 5 | 0.5 (0.2) | 0.5 (0.2) | 0.6 (0.2) | 0.5 (0.2) | 0.13 |
| **Social Vulnerability Index Theme 1, mean (SD)** | 5 | 0.5 (0.2) | 0.5 (0.2) | 0.5 (0.2) | 0.5 (0.2) | 0.0101* |
| **Social Vulnerability Index Theme 2, mean (SD)** | 3 | -0.2 (25.1) | 0.5 (0.2) | -2.7 (51.6) | 0.1 (21.5) | 0.0037* |
| **Social Vulnerability Index Theme 3, mean (SD)** | 1 | 0.6 (0.2) | 0.6 (0.2) | 0.6 (0.2) | 0.6 (0.2) | 0.0433* |
| **Social Vulnerability Index Theme 4, mean (SD)** | 3 | 0.5 (0.2) | 0.5 (0.2) | 0.6 (0.2) | 0.5 (0.2) | 0.0002* |
| **County Digital Infrastructure** |  |  |  |  |  |  |
| **at least one device (%), mean (SD)** | 30 | 93.8 (3.2) | 94.0 (3.3) | 93.7 (3.3) | 93.7 (3.1) | 0.05 |
| **broadband connection (%), mean (SD)** | 30 | 88.0 (5.2) | 88.1 (5.4) | 87.7 (5.6) | 88.0 (5.0) | 0.4375 |
| **internet connection (%), mean (SD)** | 30 | 90.8 (4.7) | 91.0 (4.8) | 90.5 (5.2) | 90.7 (4.5) | 0.1316 |
| **Hospital Service Area Healthcare Access and provider shortage** |  |  |  |  |  |  |
| **Primary Health Professional Shortage Score, mean (SD)** | 0 | 8.5 (7.4) | 8.2 (7.3) | 8.7 (7.6) | 8.5 (7.5) | 0.3752 |
| **Dental Health Professional Shortage Score, mean (SD)** | 0 | 7.9 (8.1) | 7.9 (8.0) | 7.3 (8.0) | 8.0 (8.1) | 0.2808 |
| **Mental Health Health Professional Shortage Score, mean (SD)** | 0 | 11.8 (7.1) | 11.5 (7.1) | 12.0 (6.9) | 11.8 (7.2) | 0.3798 |
| **Medical Underserve Area Score, mean (SD)** | 0 | 22.5 (27.7) | 21.3 (27.3) | 21.9 (27.8) | 23.2 (27.8) | 0.2004 |
| **Elders Medical Underservice Area Score, mean (SD)** | 0 | 1.8 (5.1) | 1.8 (5.1) | 2.3 (5.7) | 1.8 (5.0) | 0.1393 |
| **Infant Medical Underservice Area Score, mean (SD)** | 0 | 3.2 (8.4) | 3.1 (8.3) | 3.9 (9.1) | 3.1 (8.4) | 0.2109 |

**Table S5.** Descriptive statistics by Census Division

Variable definitions are provided in Table S3. To prevent re-identification through cross-tabulation, we suppressed cells with counts below 20 along with adjacent cells as needed. Continuous variables are presented as means with standard deviations, while categorical variables are shown as frequencies with percentages. Statistical comparisons across groups were conducted using ANOVA for continuous variables and chi-square tests for categorical variables. ANOVA assumes normality within groups, homogeneity of variance, and independence of observations; chi-square tests assume independence and adequate expected cell frequencies. These assumptions were assumed to be adequately met for descriptive comparisons. All statistical tests were two-sided with significance set at p<0.05. Bonferroni correction was applied to correct for multiple comparisons.

| **Census Region** |  |  | **Northeast** | | **South** | | | **Midwest** | | **West** | | **P** |
| --- | --- | --- | --- | --- | --- | --- | --- | --- | --- | --- | --- | --- |
| **Census Division** | **Missing** | **Overall** | **NE** | **MA** | **SA** | **ESC** | **WSC** | **ENC** | **WNC** | **MTN** | **PAC** |  |
|  |  |  | CT,  ME,  MA,  NH,  RI,  VT | NJ, NY, PA | DE, DC, FL, GA, MD, NC, SC, VA, WV | AL, KY, MS, TN | AR, LA, OK, TX | IL, IN, MI, OH, WI | IA, KS, MN, MO, NE, ND, SD | AZ, CO, ID, MT, NV, NM, UT, WY | AK, CA, HI, OR, WA |  |
| **n** |  | 3560 | 139 | 335 | 565 | 222 | 588 | 497 | 541 | 306 | 367 |  |
| **Hospital characteristics** |  |  |  |  |  |  |  |  |  |  |  |  |
| **Healthcare system structure and organizational characteristics** |  |  |  |  |  |  |  |  |  |  |  |  |
| **System member, n (%)** |  |  |  |  |  |  |  |  |  |  |  |  |
| Yes |  | 2606 (73.2) | 102 (73.4) | 260 (77.6) | 487 (86.2) | 181 (81.5) | 373 (63.4) | 372 (74.8) | 345 (63.8) | 211 (69.0) | 275 (74.9) | <0.0001* |
| **Community hospital, n (%)** |  |  |  |  |  |  |  |  |  |  |  |  |
| Yes |  | 3235 (90.9) | 123 (88.5) | 280 (83.6) | 523 (92.6) | 195 (87.8) | 519 (88.3) | 455 (91.5) | 512 (94.6) | 284 (92.8) | 344 (93.7) | <0.0001* |
| **Ownership type, n (%)** |  |  |  |  |  |  |  |  |  |  |  |  |
| Federal-government |  | 44 (1.2) | NA | NA | NA | NA | NA | NA | NA | NA | NA | <0.0001* |
| Nonfederal-government |  | 649 (18.2) | NA | 50 (14.9) | 86 (15.2) | 51 (23.0) | 143 (24.3) | 47 (9.5) | 150 (27.7) | 46 (15.0) | 67 (18.3) |  |
| nonprofit |  | 2209 (62.1) | 117 (84.2) | 250 (74.6) | 342 (60.5) | 113 (50.9) | 201 (34.2) | 394 (79.3) | 338 (62.5) | 193 (63.1) | 261 (71.1) |  |
| for profit |  | 658 (18.5) | NA | NA | 128 (22.7) | 54 (24.3) | 237 (40.3) | 50 (10.1) | 50 (9.2) | 62 (20.3) | 34 (9.3) |  |
| **Delivery system, n (%)** |  |  |  |  |  |  |  |  |  |  |  |  |
| Centralized Health System |  | 488 (13.7) | 18 (12.9) | 80 (23.9) | 118 (20.9) | 28 (12.6) | 19 (3.2) | 78 (15.7) | 40 (7.4) | 62 (20.3) | 45 (12.3) | <0.0001* |
| Centralized Physician/Insurance Health System |  | 187 (5.3) | NA | 41 (12.2) | NA | NA | 41 (7.0) | 37 (7.4) | NA | 31 (10.1) | 13 (3.5) |  |
| Moderately Centralized Health System |  | 684 (19.2) | 39 (28.1) | 51 (15.2) | 124 (21.9) | 66 (29.7) | 112 (19.0) | 123 (24.7) | 80 (14.8) | NA | 69 (18.8) |  |
| Decentralized Health System |  | 632 (17.8) | 22 (15.8) | NA | 110 (19.5) | 22 (9.9) | 69 (11.7) | 64 (12.9) | 170 (31.4) | 55 (18.0) | 110 (30.0) |  |
| Independent Hospital System |  | 615 (17.3) | NA | 78 (23.3) | 124 (21.9) | 61 (27.5) | 132 (22.4) | 70 (14.1) | 51 (9.4) | 43 (14.1) | 38 (10.4) |  |
| No/No Response |  | 954 (26.8) | 37 (26.6) | 75 (22.4) | 78 (13.8) | 41 (18.5) | 215 (36.6) | 125 (25.2) | 196 (36.2) | 95 (31.0) | 92 (25.1) |  |
| **Children hospital, n(%)** |  |  |  |  |  |  |  |  |  |  |  |  |
| Yes |  | 75 (2.1) | NA | NA | NA | NA | NA | NA | NA | NA | NA | 0.39 |
| **Teaching hospital , n (%)** |  |  |  |  |  |  |  |  |  |  |  |  |
| Yes |  | 1742 (48.9) | 94 (67.6) | 247 (73.7) | 319 (56.5) | 93 (41.9) | 218 (37.1) | 254 (51.1) | 149 (27.5) | 133 (43.5) | 235 (64.0) | <0.0001* |
| **Major teaching hospital, n (%)** |  |  |  |  |  |  |  |  |  |  |  |  |
| Yes |  | 205 (5.8) | NA | 36 (10.7) | 42 (7.4) | NA | NA | 31 (6.2) | NA | NA | NA | <0.0001* |
| **Minor teaching hospital, n (%)** |  |  |  |  |  |  |  |  |  |  |  |  |
| Yes |  | 1537 (43.2) | 75 (54.0) | 211 (63.0) | 277 (49.0) | 82 (36.9) | 198 (33.7) | 223 (44.9) | 135 (25.0) | 119 (38.9) | 217 (59.1) | <0.0001* |
| **Joint commision accreditation, n(%)** |  |  |  |  |  |  |  |  |  |  |  |  |
| Yes |  | 2348 (66.0) | 110 (79.1) | 271 (80.9) | 448 (79.3) | 153 (68.9) | 349 (59.4) | 337 (67.8) | 205 (37.9) | 190 (62.1) | 285 (77.7) | <0.0001* |
| **Critical access hospital, n (%)** |  |  |  |  |  |  |  |  |  |  |  |  |
| Yes |  | 830 (23.3) | 23 (16.5) | NA | 61 (10.8) | 31 (14.0) | 114 (19.4) | 117 (23.5) | 295 (54.5) | 100 (32.7) | 72 (19.6) | <0.0001* |
| **Rural referral center, n (%)** |  |  |  |  |  |  |  |  |  |  |  |  |
| Yes |  | 533 (15.0) | 34 (24.5) | 66 (19.7) | 98 (17.3) | 29 (13.1) | 55 (9.4) | 103 (20.7) | 38 (7.0) | 46 (15.0) | 64 (17.4) | <0.0001* |
| **Operation of subsidiary hospital, n (%)** |  |  |  |  |  |  |  |  |  |  |  |  |
| Yes |  | 469 (13.2) | 30 (21.6) | 70 (20.9) | 80 (14.2) | NA | 61 (10.4) | 83 (16.7) | 55 (10.2) | NA | 50 (13.6) | <0.0001* |
| **Frontline facility, n (%)** |  |  |  |  |  |  |  |  |  |  |  |  |
| Yes |  | 370 (10.4) | NA | 57 (17.0) | 65 (11.5) | NA | 52 (8.8) | 63 (12.7) | 40 (7.4) | 25 (8.2) | 37 (10.1) | 0.0003* |
| **Hospital size, n(%)** |  |  |  |  |  |  |  |  |  |  |  |  |
| **Bedsize, n (%)** |  |  |  |  |  |  |  |  |  |  |  |  |
| 6-24 beds |  | 480 (13.5) | NA | NA | 34 (6.0) | NA | 130 (22.1) | 56 (11.3) | 142 (26.2) | 53 (17.3) | 31 (8.4) | <0.0001* |
| 25-49 beds |  | 761 (21.4) | 27 (19.4) | 22 (6.6) | 81 (14.3) | 63 (28.4) | 150 (25.5) | 125 (25.2) | 150 (27.7) | 91 (29.7) | 52 (14.2) |  |
| 50-99 beds |  | 635 (17.8) | 28 (20.1) | 56 (16.7) | 109 (19.3) | 43 (19.4) | 111 (18.9) | 85 (17.1) | 100 (18.5) | 53 (17.3) | 50 (13.6) |  |
| 100-199 beds |  | 671 (18.8) | 35 (25.2) | 87 (26.0) | 129 (22.8) | 49 (22.1) | 70 (11.9) | 99 (19.9) | 77 (14.2) | 38 (12.4) | 87 (23.7) |  |
| 200-299 beds |  | 380 (10.7) | 20 (14.4) | 48 (14.3) | 86 (15.2) | NA | 49 (8.3) | 54 (10.9) | 25 (4.6) | 25 (8.2) | 55 (15.0) |  |
| 300-399 beds |  | 245 (6.9) | NA | 45 (13.4) | 40 (7.1) | NA | 30 (5.1) | 28 (5.6) | NA | 21 (6.9) | 43 (11.7) |  |
| 400-499 beds |  | 128 (3.6) | NA | NA | 25 (4.4) | NA | NA | 22 (4.4) | NA | NA | 26 (7.1) |  |
| 500 or more beds |  | 260 (7.3) | NA | 49 (14.6) | 61 (10.8) | NA | 36 (6.1) | 28 (5.6) | NA | NA | 23 (6.3) |  |
| **Healthcare payer mix** |  |  |  |  |  |  |  |  |  |  |  |  |
| **Medicare inpatient days percentage, mean (SD)** | 1 | 51.6 (22.0) | 51.7 (20.6) | 47.0 (22.7) | 52.8 (18.5) | 50.5 (22.7) | 52.5 (24.6) | 55.2 (20.3) | 54.4 (25.1) | 48.9 (20.5) | 46.5 (18.5) | <0.0001* |
| **Medicaid inpatient days percentage, mean (SD)** | 1 | 19.0 (15.7) | 20.2 (15.0) | 21.3 (16.5) | 17.7 (13.0) | 18.0 (15.3) | 14.2 (15.4) | 18.3 (13.3) | 18.6 (16.5) | 20.5 (14.7) | 27.4 (18.2) | <0.0001* |
| **Hospital Interoperability capability** |  |  |  |  |  |  |  |  |  |  |  |  |
| **Core index, mean (SD)** | 468 | 3.5 (1.8) | 4.0 (1.8) | 3.8 (1.8) | 4.0 (1.6) | 3.3 (1.8) | 2.9 (2.0) | 3.8 (1.8) | 3.4 (1.7) | 3.4 (1.6) | 3.6 (1.8) | <0.0001* |
| **Friction index, mean (SD)** | 468 | 0.8 (0.4) | 0.8 (0.4) | 0.8 (0.4) | 0.9 (0.4) | 0.7 (0.4) | 0.7 (0.4) | 0.8 (0.4) | 0.8 (0.4) | 0.8 (0.4) | 1.0 (0.5) | <0.0001* |
| **Geographic community characteristics** |  |  |  |  |  |  |  |  |  |  |  |  |
| **Hospital Service Area geographic socioeconomic context** |  |  |  |  |  |  |  |  |  |  |  |  |
| **Rural urban category, n (%)** |  |  |  |  |  |  |  |  |  |  |  |  |
| Metro |  | 2398 (67.4) | 100 (71.9) | 282 (84.2) | 448 (79.3) | 134 (60.4) | 401 (68.2) | 333 (67.0) | 210 (38.8) | 191 (62.4) | 299 (81.5) | <0.0001* |
| Micro |  | 522 (14.7) | 23 (16.5) | 35 (10.4) | 54 (9.6) | 47 (21.2) | 78 (13.3) | 97 (19.5) | 95 (17.6) | 48 (15.7) | 45 (12.3) |  |
| Rural |  | 640 (18.0) | 16 (11.5) | 18 (5.4) | 63 (11.2) | 41 (18.5) | 109 (18.5) | 67 (13.5) | 236 (43.6) | 67 (21.9) | 23 (6.3) |  |
| **National Area Deprivation Index, mean (SD)** | 4 | 57.8 (24.9) | 37.8 (20.7) | 43.3 (26.0) | 57.5 (23.1) | 76.0 (15.8) | 70.4 (18.0) | 66.4 (17.1) | 72.0 (13.7) | 45.8 (20.0) | 25.3 (19.0) | <0.0001* |
| **Overall Social Vulnerability Index, mean (SD)** | 5 | 0.5 (0.2) | 0.4 (0.2) | 0.5 (0.2) | 0.6 (0.2) | 0.6 (0.2) | 0.7 (0.2) | 0.5 (0.2) | 0.4 (0.2) | 0.5 (0.2) | 0.7 (0.2) | <0.0001* |
| **Social Vulnerability Index Theme 1, mean (SD)** | 5 | 0.5 (0.2) | 0.4 (0.2) | 0.4 (0.2) | 0.6 (0.2) | 0.6 (0.2) | 0.7 (0.2) | 0.4 (0.2) | 0.4 (0.2) | 0.5 (0.2) | 0.6 (0.2) | <0.0001* |
| **Social Vulnerability Index Theme 2, mean (SD)** | 3 | 0.5 (0.2) | 0.4 (0.2) | 0.5 (0.2) | 0.6 (0.2) | 0.5 (0.2) | 0.7 (0.2) | 0.5 (0.2) | 0.4 (0.2) | 0.5 (0.2) | 0.6 (0.2) | <0.0001* |
| **Social Vulnerability Index Theme 3, mean (SD)** | 1 | 0.6 (0.2) | 0.5 (0.2) | 0.5 (0.2) | 0.7 (0.2) | 0.6 (0.2) | 0.7 (0.2) | 0.4 (0.2) | 0.4 (0.2) | 0.6 (0.2) | 0.8 (0.2) | <0.0001* |
| **Social Vulnerability Index Theme 4, mean (SD)** | 3 | 0.5 (0.2) | 0.5 (0.2) | 0.6 (0.2) | 0.6 (0.2) | 0.5 (0.2) | 0.6 (0.2) | 0.5 (0.2) | 0.4 (0.2) | 0.5 (0.2) | 0.7 (0.2) | <0.0001* |
| **County Digital Infrastructure** |  |  |  |  |  |  |  |  |  |  |  |  |
| **at lease one device (%), mean (SD)** | 30 | 93.8 (3.2) | 94.3 (1.9) | 93.6 (2.3) | 94.0 (3.5) | 92.3 (3.5) | 93.8 (3.3) | 93.3 (2.7) | 92.9 (2.9) | 94.7 (4.2) | 95.9 (1.6) | <0.0001* |
| **broadband connection (%), mean (SD)** | 30 | 88.0 (5.2) | 89.8 (3.6) | 89.4 (3.4) | 87.9 (5.4) | 84.9 (6.5) | 86.8 (5.8) | 87.9 (3.7) | 86.7 (4.4) | 88.7 (6.7) | 91.5 (3.0) | <0.0001* |
| **internet connection (%), mean (SD)** | 30 | 90.8 (4.7) | 92.8 (2.5) | 91.7 (2.9) | 90.8 (4.9) | 88.0 (5.4) | 89.4 (5.3) | 90.7 (3.5) | 89.8 (4.0) | 91.7 (6.2) | 93.9 (2.6) | <0.0001* |
| **Hospital Service Area Healthcare Access and provider shortage** |  |  |  |  |  |  |  |  |  |  |  |  |
| **Primary Health Professional Shortage Score, mean (SD)** | 30 | 93.8 (3.2) | 94.3 (1.9) | 93.6 (2.3) | 94.0 (3.5) | 92.3 (3.5) | 93.8 (3.3) | 93.3 (2.7) | 92.9 (2.9) | 94.7 (4.2) | 95.9 (1.6) | <0.0001* |
| **Dental Health Professional Shortage Score, mean (SD)** | 30 | 88.0 (5.2) | 89.8 (3.6) | 89.4 (3.4) | 87.9 (5.4) | 84.9 (6.5) | 86.8 (5.8) | 87.9 (3.7) | 86.7 (4.4) | 88.7 (6.7) | 91.5 (3.0) | <0.0001* |
| **Mental Health Health Professional Shortage Score, mean (SD)** | 30 | 90.8 (4.7) | 92.8 (2.5) | 91.7 (2.9) | 90.8 (4.9) | 88.0 (5.4) | 89.4 (5.3) | 90.7 (3.5) | 89.8 (4.0) | 91.7 (6.2) | 93.9 (2.6) | <0.0001* |
| **Medical Underserve Area Score, mean (SD)** | 30 | 93.8 (3.2) | 94.3 (1.9) | 93.6 (2.3) | 94.0 (3.5) | 92.3 (3.5) | 93.8 (3.3) | 93.3 (2.7) | 92.9 (2.9) | 94.7 (4.2) | 95.9 (1.6) | <0.0001* |
| **Elders Medical Underservice Area Score, mean (SD)** | 30 | 88.0 (5.2) | 89.8 (3.6) | 89.4 (3.4) | 87.9 (5.4) | 84.9 (6.5) | 86.8 (5.8) | 87.9 (3.7) | 86.7 (4.4) | 88.7 (6.7) | 91.5 (3.0) | <0.0001* |
| **Infant Medical Underservice Area Score, mean (SD)** | 30 | 90.8 (4.7) | 92.8 (2.5) | 91.7 (2.9) | 90.8 (4.9) | 88.0 (5.4) | 89.4 (5.3) | 90.7 (3.5) | 89.8 (4.0) | 91.7 (6.2) | 93.9 (2.6) | <0.0001* |

**Table S6.** Descriptive statistics by U.S. state (Available in Supplementary Data)

Variable definitions are provided in Table S3. To prevent re-identification through cross-tabulation, we suppressed cells with counts below 20 along with adjacent cells as needed. Continuous variables are presented as means with standard deviations, while categorical variables are shown as frequencies with percentages. Statistical comparisons across groups were conducted using ANOVA for continuous variables and chi-square tests for categorical variables. ANOVA assumes normality within groups, homogeneity of variance, and independence of observations; chi-square tests assume independence and adequate expected cell frequencies. These assumptions were assumed to be adequately met for descriptive comparisons. All statistical tests were two-sided with significance set at p<0.05. Bonferroni correction was applied to correct for multiple comparisons.

**Table S7.** Descriptive statistics by hospital AI implementation level

Variable definitions are provided in Table S2. To prevent re-identification through cross-tabulation, we suppressed cells with counts below 10 along with adjacent cells as needed. Continuous variables are presented as means with standard deviations, while categorical variables are shown as frequencies with percentages. Statistical comparisons across groups were conducted using ANOVA for continuous variables and chi-square tests for categorical variables. ANOVA assumes normality within groups, homogeneity of variance, and independence of observations; chi-square tests assume independence and adequate expected cell frequencies. These assumptions were assumed to be adequately met for descriptive comparisons. All statistical tests were two-sided with significance set at p<0.05. No adjustment for multiple comparisons was applied to descriptive statistics, as these were exploratory comparisons to characterize the sample rather than formal hypothesis tests.

|  |  | **AI implementation status** | | |  |
| --- | --- | --- | --- | --- | --- |
|  | **Missing** | **No predictive model** | **Non-AI predictive model** | **AI-based predictive model** | **P-Value** |
| **n** |  | 1043 | 568 | 1738 |  |
| **Secondary Predictive Model Implementation Measures** |  |  |  |  |  |
| **Model breadth implementation, mean (SD)** | 213 | 0.0 (0.0) | 0.5 (0.1) | 0.8 (0.1) | <0.0001 |
| **Model development score, mean (SD)** | 211 | 0.0 (0.0) | 0.5 (0.2) | 0.7 (0.2) | <0.0001 |
| **Model evaluation score 2023 version, mean (SD)** | 211 | 0.0 (0.0) | 0.5 (0.2) | 0.8 (0.2) | <0.0001 |
| **Model evaluation score 2024 version, mean (SD)** | 1030 | 0.0 (0.0) | 0.5 (0.2) | 0.7 (0.2) | <0.0001 |
| **LLM readiness score** |  |  |  |  |  |
| **LLM readiness score, mean (SD)** | 1327 | 0.2 (0.3) | 0.6 (0.3) | 0.7 (0.4) | <0.0001 |
| **Predictive Model Use Case** |  |  |  |  |  |
| **Predicting health trajectories or risks for inpatients, n (%)** |  |  |  |  |  |
| Yes |  | 0 (0) | 508 (89.4) | 1628 (93.7) | <0.0001 |
| **Identify high risk outpatients to inform follow-up care, n (%)** |  |  |  |  |  |
| Yes |  | 0 (0) | 476 (83.8) | 1559 (89.7) | <0.0001 |
| **Monitor health, n (%)** |  |  |  |  |  |
| Yes |  | 0 (0) | 170 (29.9) | 650 (37.4) | <0.0001 |
| **Recommend treatments, n (%)** |  |  |  |  |  |
| Yes |  | 0 (0) | 180 (31.7) | 861 (49.5) | <0.0001 |
| **Simplify or automate billing procedures, n (%)** |  |  |  |  |  |
| Yes |  | 0 (0) | 342 (60.2) | 923 (53.1) | <0.0001 |
| **Facilitate scheduling, n (%)** |  |  |  |  |  |
| Yes |  | 0 (0) | 319 (56.2) | 1087 (62.5) | <0.0001 |
| **Other operational process optimization, n (%)** |  |  |  |  |  |
| Yes |  | 0 (0) | 137 (24.1) | 373 (21.5) | <0.0001 |
| **Other clinical use case, n (%)** |  |  |  |  |  |
| Yes |  | 0 (0) | 164 (28.9) | 411 (23.6) | <0.0001 |
| **Predictive Model Developer** |  |  |  |  |  |
| **EHR developer, n (%)** |  |  |  |  |  |
| Yes |  | 0 (0) | 350 (61.6) | 1586 (91.3) | <0.0001 |
| **Third-party developer, n (%)** |  |  |  |  |  |
| Yes |  | 0 (0) | 389 (68.5) | 797 (45.9) | <0.0001 |
| **Self-development, n (%)** |  |  |  |  |  |
| Yes |  | 0 (0) | 248 (43.7) | 884 (50.9) | <0.0001 |
| **Public domain, n (%)** |  |  |  |  |  |
| Yes |  | 0 (0) | NA | 65 (3.7) | <0.0001 |
| **Predictive Model Evaluation** |  |  |  |  |  |
| **Model accuracy, n (%)** |  |  |  |  |  |
| All models |  | 0 (0) | 73 (12.9) | 956 (55.0) | <0.0001 |
| Most models |  | 0 (0) | 227 (40.0) | 323 (18.6) |  |
| Some models |  | 0 (0) | 60 (10.6) | 85 (4.9) |  |
| Few models |  | 0 (0) | 22 (3.9) | NA |  |
| Do not know/None |  | 581 (55.7) | 125 (22.0) | 282 (16.2) |  |
| No response |  | 427 (40.9) | 61 (10.7) | NA |  |
| **Model bias, n (%)** |  |  |  |  |  |
| All models |  | 0 (0) | 49 (8.6) | 730 (42.0) | <0.0001 |
| Most models |  | 0 (0) | 186 (32.7) | 296 (17.0) |  |
| Some models |  | 0 (0) | 113 (19.9) | 178 (10.2) |  |
| Few models |  | 0 (0) | NA | 118 (6.8) |  |
| Do not know/None |  | 581 (55.7) | 181 (31.9) | 395 (22.7) |  |
| No response |  | 427 (40.9) | NA | 21 (1.2) |  |
| **Post-implementation, n (%)** |  |  |  |  |  |
| All models |  | 0 (0) | NA | 600 (34.5) | <0.0001 |
| Most models |  | 0 (0) | 171 (30.1) | 182 (10.5) |  |
| Some models |  | 0 (0) | 48 (8.5) | 140 (8.1) |  |
| Few models |  | 0 (0) | NA | 167 (9.6) |  |
| Do not know/None |  | 581 (55.7) | 125 (22.0) | 282 (16.2) |  |
| No response |  | 448 (43.0) | 213 (37.5) | 406 (23.4) |  |
| **Model evaluation responsible personnel^a^** |  |  |  |  |  |
| ***Designated Senior Executive (CMIO/CIO/Director of Technology), n (%)*** |  |  |  |  |  |
| Yes |  | 200 (19.2) | 243 (42.8) | 701 (40.3) | <0.0001 |
| **Specific Committee or Task Force for Machine Learning or Predictive Modeling, n(%)** |  |  |  |  |  |
| Yes |  | 70 (6.7) | 239 (42.1) | 901 (51.8) | <0.0001 |
| **Clinical Decision Support Committee/other non-Machine Learning or Predictive Modeling specific Committee, n(%)** |  |  |  |  |  |
| Yes |  | 124 (11.9) | 272 (47.9) | 666 (38.3) | <0.0001 |
| **Division/Department Leaders, n(%)** |  |  |  |  |  |
| Yes |  | 110 (10.5) | 221 (38.9) | 787 (45.3) | <0.0001 |
| **IT staff, n(%)** |  |  |  |  |  |
| Yes |  | 158 (15.1) | 87 (15.3) | 640 (36.8) |  |
| **Hospital characteristics** |  |  |  |  |  |
| **System member, n (%)** |  |  |  |  |  |
| Yes |  | 474 (45.4) | 483 (85.0) | 1547 (89.0) | <0.0001 |
| **Community hospital, n (%)** |  |  |  |  |  |
| Yes |  | 852 (81.7) | 549 (96.7) | 1690 (97.2) | <0.0001 |
| **Ownership type, n (%)** |  |  |  |  |  |
| nonfederal government |  | 362 (34.7) | 43 (7.6) | 185 (10.6) | <0.0001 |
| nonprofit |  | 438 (42.0) | 362 (63.7) | 1336 (76.9) |  |
| for profit |  | 226 (21.7) | 157 (27.6) | 209 (12.0) |  |
| **Delivery system, n (%)** |  |  |  |  |  |
| Centralized Health System |  | NA | 66 (11.6) | 399 (23.0) | <0.0001 |
| Centralized Physician/Insurance Health System |  | NA | 41 (7.2) | 125 (7.2) |  |
| Moderately Centralized Health System |  | 161 (15.4) | 88 (15.5) | 396 (22.8) |  |
| Decentralized Health System |  | 74 (7.1) | 235 (41.4) | 316 (18.2) |  |
| Independent Hospital System |  | 205 (19.7) | 53 (9.3) | 311 (17.9) |  |
| No/No Response |  | 569 (54.6) | 85 (15.0) | 191 (11.0) |  |
| **Children hospital, n(%)** |  |  |  |  |  |
| Yes |  | 29 (2.8) | NA | 29 (1.7) | 0.095 |
| **Teaching hospital , n (%)** |  |  |  |  |  |
| Yes |  | 368 (35.3) | 361 (63.6) | 942 (54.2) | <0.0001 |
| **Major teaching hospital, n (%)** |  |  |  |  |  |
| Yes |  | 21 (2.0) | 23 (4.0) | 157 (9.0) | <0.0001 |
| **Minor teaching hospital, n (%)** |  |  |  |  |  |
| Yes |  | 347 (33.3) | 338 (59.5) | 785 (45.2) | <0.0001 |
| **Bed size, n (%)** |  |  |  |  |  |
| 6-24 beds |  | 210 (20.1) | 60 (10.6) | 169 (9.7) | <0.0001 |
| 25-49 beds |  | 315 (30.2) | 80 (14.1) | 314 (18.1) |  |
| 50-99 beds |  | 174 (16.7) | 82 (14.4) | 326 (18.8) |  |
| 100-199 beds |  | 199 (19.1) | 123 (21.7) | 317 (18.2) |  |
| 200-299 beds |  | 82 (7.9) | 76 (13.4) | 208 (12.0) |  |
| 300-399 beds |  | 36 (3.5) | 56 (9.9) | 145 (8.3) |  |
| 400-499 beds |  | 10 (1.0) | 37 (6.5) | 78 (4.5) |  |
| 500 or more beds |  | 17 (1.6) | 54 (9.5) | 181 (10.4) |  |
| **Joint commission accreditation, n(%)** |  |  |  |  |  |
| Yes |  | 508 (48.7) | 472 (83.1) | 1257 (72.3) | <0.0001 |
| **Critical access hospital, n (%)** |  |  |  |  |  |
| Yes |  | 377 (36.1) | 90 (15.8) | 307 (17.7) | <0.0001 |
| **Rural referral center, n (%)** |  |  |  |  |  |
| Yes |  | 65 (6.2) | 110 (19.4) | 348 (20.0) | <0.0001 |
| **Operation of subsidiary hospital, n (%)** |  |  |  |  |  |
| Yes |  | 96 (9.2) | 74 (13.0) | 283 (16.3) | <0.0001 |
| **Frontline facility, n (%)** |  |  |  |  |  |
| Yes |  | 35 (3.4) | 76 (13.4) | 252 (14.5) | <0.0001 |
| **Medicare inpatient days percentage, mean (SD)** | 1 | 47.5 (26.6) | 53.8 (16.7) | 54.7 (18.5) | <0.0001 |
| **Medicaid inpatient days percentage, mean (SD)** | 1 | 18.7 (18.4) | 18.8 (12.5) | 19.2 (14.3) | 0.64 |
| **Core index, mean (SD)** | 468 | 2.0 (1.4) | 4.2 (1.3) | 4.5 (1.3) | <0.0001 |
| **Friction index, mean (SD)** | 468 | 0.7 (0.4) | 1.0 (0.4) | 0.9 (0.4) | <0.0001 |
| **Rural urban category, n (%)** |  |  |  |  |  |
| Metro |  | 552 (52.9) | 431 (75.9) | 1291 (74.3) | <0.0001 |
| Micro |  | 179 (17.2) | 81 (14.3) | 233 (13.4) |  |
| Rural |  | 312 (29.9) | 56 (9.9) | 214 (12.3) |  |
| **National Area Deprivation Index, mean (SD)** | 4 | 63.5 (24.5) | 52.9 (25.1) | 55.3 (24.3) | <0.0001 |
| **Overall Social Vulnerability Index, mean (SD)** | 5 | 0.5 (0.2) | 0.6 (0.2) | 0.5 (0.2) | 0.008 |
| **Social Vulnerability Index Theme 1, mean (SD)** | 5 | 0.5 (0.2) | 0.5 (0.2) | 0.5 (0.2) | <0.0001 |
| **Social Vulnerability Index Theme 2, mean (SD)** | 3 | 0.5 (0.2) | 0.5 (0.2) | 0.5 (0.2) | 0.33 |
| **Social Vulnerability Index Theme 3, mean (SD)** | 1 | 0.6 (0.3) | 0.6 (0.2) | 0.6 (0.2) | <0.0001 |
| **Social Vulnerability Index Theme 4, mean (SD)** | 3 | 0.5 (0.2) | 0.6 (0.2) | 0.5 (0.2) | <0.0001 |
| **at lease one device (%), mean (SD)** | 30 | 92.9 (3.6) | 94.4 (2.9) | 94.2 (2.9) | <0.0001 |
| **broadband connection (%), mean (SD)** | 30 | 86.4 (5.9) | 89.0 (4.4) | 88.7 (4.7) | <0.0001 |
| **internet connection (%), mean (SD)** | 30 | 89.4 (5.4) | 91.7 (4.1) | 91.4 (4.2) | <0.0001 |
| **Primary Health Professional Shortage Score, mean (SD)** | 0 | 9.8 (7.0) | 8.6 (7.5) | 7.5 (7.5) | <0.0001 |
| **Dental Health Professional Shortage Score, mean (SD)** | 0 | 8.6 (7.9) | 8.4 (8.2) | 7.2 (8.1) | <0.0001 |
| **Mental Health Health Professional Shortage Score, mean (SD)** | 0 | 13.0 (6.6) | 12.0 (6.9) | 10.8 (7.4) | <0.0001 |
| **Medical Underserve Area Score, mean (SD)** | 0 | 25.8 (28.3) | 21.9 (27.5) | 20.6 (27.2) | <0.0001 |
| **Elders Medical Underservice Area Score, mean (SD)** | 0 | 2.4 (5.7) | 1.6 (4.8) | 1.6 (4.8) | <0.0001 |
| **Infant Medical Underservice Area Score, mean (SD)** | 0 | 4.3 (9.5) | 3.0 (8.2) | 2.6 (7.7) | <0.0001 |

**Table S8.** Need indicator tertile distribution

| **Measure** | **Low Need Range** | **Medium Need Range** | **High Need Range** | **Low Need N** | **Medium Need N** | **High Need N** |
| --- | --- | --- | --- | --- | --- | --- |
| **Primary HPSA Score** | 0.000 - 0.000 | 0.000 - 14.000 | 14.000 - 23.000 | 1187 | 1186 | 1187 |
| **Mental HPSA Score** | 0.000 - 12.000 | 12.000 - 17.000 | 17.000 - 23.000 | 1187 | 1186 | 1187 |
| **Dental HPSA Score** | 0.000 - 0.000 | 0.000 - 15.000 | 15.000 - 25.000 | 1187 | 1186 | 1187 |
| **MUA Overall Score** | 0.000 - 0.000 | 0.000 - 50.100 | 50.200 - 91.900 | 1187 | 1186 | 1187 |
| **MUA Elder Score** | 0.000 - 0.000 | 0.000 - 0.000 | 0.000 - 20.200 | 1187 | 1186 | 1187 |
| **MUA Infant Score** | 0.000 - 0.000 | 0.000 - 0.000 | 0.000 - 26.000 | 1187 | 1186 | 1187 |
| **Area Deprivation Index** | 1.00 - 48.50 | 49.00 - 74.00 | 74.50 - 100.00 | 1187 | 1206 | 1163 |
| **Social Vulnerability Index** | 0.03 - 0.43 | 0.43 - 0.66 | 0.66 - 0.99 | 1185 | 1186 | 1184 |
| **SVI Theme 1 (Socioeconomic)** | 0.00 - 0.38 | 0.38 - 0.63 | 0.63 - 0.99 | 1186 | 1184 | 1185 |
| **SVI Theme 2 (Household)** | 0.01 - 0.44 | 0.44 - 0.61 | 0.61 - 1.00 | 1186 | 1187 | 1184 |
| **SVI Theme 3 (Minority)** | 0.00 - 0.44 | 0.44 - 0.73 | 0.73 - 0.99 | 1187 | 1191 | 1181 |
| **SVI Theme 4 (Housing/Transport)** | 0.00 - 0.45 | 0.45 - 0.63 | 0.63 - 1.00 | 1187 | 1185 | 1185 |

**Table S9.** Moran’s I results by AI census division and state (available in Supplementary Data)

Census division abbreviations: NE=New England, MA=Mid Atlantic, SA=South Atlantic, ENC=East North Central, WNC=West North Central, ESC=East South Central, WSC=West South Central, MTN=Mountain, PAC=Pacific. State abbreviations: AL=Alabama, AK=Alaska, AZ=Arizona, AR=Arkansas, CA=California, CO=Colorado, CT=Connecticut, DE=Delaware, FL=Florida, GA=Georgia, HI=Hawaii, ID=Idaho, IL=Illinois, IN=Indiana, IA=Iowa, KS=Kansas, KY=Kentucky, LA=Louisiana, ME=Maine, MD=Maryland, MA=Massachusetts, MI=Michigan, MN=Minnesota, MS=Mississippi, MO=Missouri, MT=Montana, NE=Nebraska, NV=Nevada, NH=New Hampshire, NJ=New Jersey, NM=New Mexico, NY=New York, NC=North Carolina, ND=North Dakota, OH=Ohio, OK=Oklahoma, OR=Oregon, PA=Pennsylvania, RI=Rhode Island, SC=South Carolina, SD=South Dakota, TN=Tennessee, TX=Texas, UT=Utah, VT=Vermont, VA=Virginia, WA=Washington, WV=West Virginia, WI=Wisconsin, WY=Wyoming; States with fewer than 20 hospitals were suppressed. Census divisions and states are listed in alphabetical order.

**Table S10.** DBSCAN clustering results by census division

Hospitals were clustered using the DBSCAN algorithm based on six AI implementation indicators (AI base implementation score, model implementation breadth, development, evaluation in 2023 and 2024, and LLM readiness). Results are stratified by U.S. census division.

| **AI base Implementation score** | | | | | | |
| --- | --- | --- | --- | --- | --- | --- |
| **Division** | **Clusters (n)** | **Total hospitals (n)** | **Average cluster size** | **Average score** | **Minimum Score** | **Maximum score** |
| **TOTAL** | 272 | 3328 | 12.2 | -0.26 | -1.25 | 0.95 |
| **East North Central** | 38 | 464 | 12.2 | -0.47 | -1.25 | 0.95 |
| **East South Central** | 40 | 186 | 4.6 | -0.12 | -1.25 | 0.95 |
| **Mid Atlantic** | 35 | 308 | 8.8 | -0.24 | -1.25 | 0.95 |
| **Mountain** | 38 | 279 | 7.3 | -0.56 | -1.25 | 0.95 |
| **New England** | 17 | 108 | 6.4 | -0.02 | -1.25 | 0.95 |
| **Pacific** | 13 | 356 | 27.4 | -0.15 | -1.25 | 0.95 |
| **South Atlantic** | 25 | 547 | 21.9 | -0.33 | -1.25 | 0.95 |
| **West North Central** | 34 | 512 | 15.1 | -0.12 | -1.25 | 0.95 |
| **West South Central** | 32 | 568 | 17.8 | -0.08 | -1.25 | 0.95 |
| **Model implementation breadth score** | | | | | | |
| **Division** | **Clusters (n)** | **Total hospitals (n)** | **Average cluster size** | **Average score** | **Minimum Score** | **Maximum score** |
| **TOTAL** | 277 | 3292 | 11.9 | -0.18 | -1.26 | 1.51 |
| **East North Central** | 42 | 461 | 11 | -0.28 | -1.26 | 1.51 |
| **East South Central** | 37 | 183 | 4.9 | -0.24 | -1.26 | 1.34 |
| **Mid Atlantic** | 31 | 301 | 9.7 | -0.35 | -1.26 | 1.51 |
| **Mountain** | 38 | 274 | 7.2 | -0.45 | -1.26 | 1.51 |
| **New England** | 20 | 108 | 5.4 | 0.11 | -1.26 | 1.51 |
| **Pacific** | 10 | 353 | 35.3 | -0.02 | -1.26 | 1.34 |
| **South Atlantic** | 26 | 545 | 21 | -0.12 | -1.26 | 1.51 |
| **West North Central** | 39 | 501 | 12.8 | -0.03 | -1.26 | 1.51 |
| **West South Central** | 34 | 566 | 16.6 | 0.03 | -1.26 | 1.51 |
| **Model implementation development score** | | | | | | |
| **Division** | **Clusters (n)** | **Total hospitals (n)** | **Average cluster size** | **Average score** | **Minimum Score** | **Maximum score** |
| **TOTAL** | 310 | 3287 | 10.6 | -0.1 | -1.22 | 1.64 |
| **East North Central** | 42 | 458 | 10.9 | -0.14 | -1.22 | 1.64 |
| **East South Central** | 49 | 185 | 3.8 | -0.08 | -1.22 | 1.42 |
| **Mid Atlantic** | 32 | 294 | 9.2 | -0.32 | -1.22 | 1.64 |
| **Mountain** | 44 | 275 | 6.2 | -0.41 | -1.22 | 1.42 |
| **New England** | 23 | 107 | 4.7 | 0.01 | -1.22 | 1.42 |
| **Pacific** | 10 | 356 | 35.6 | -0.09 | -1.22 | 1.42 |
| **South Atlantic** | 33 | 544 | 16.5 | 0.09 | -1.22 | 1.64 |
| **West North Central** | 42 | 508 | 12.1 | 0.1 | -1.22 | 1.64 |
| **West South Central** | 35 | 560 | 16 | -0.04 | -1.22 | 1.42 |
| **Model implementation evaluation 2023 score** | | | | | | |
| **Division** | **Clusters (n)** | **Total hospitals (n)** | **Average cluster size** | **Average score** | **Minimum Score** | **Maximum score** |
| **TOTAL** | 302 | 3252 | 10.8 | -0.09 | -1.2 | 1.27 |
| **East North Central** | 51 | 455 | 8.9 | -0.19 | -1.2 | 1.27 |
| **East South Central** | 38 | 180 | 4.7 | -0.03 | -1.2 | 1.27 |
| **Mid Atlantic** | 33 | 297 | 9 | -0.18 | -1.2 | 1.27 |
| **Mountain** | 41 | 276 | 6.7 | -0.37 | -1.2 | 1.27 |
| **New England** | 21 | 101 | 4.8 | 0.12 | -1.2 | 1.27 |
| **Pacific** | 9 | 351 | 39 | -0.21 | -1.2 | 1.27 |
| **South Atlantic** | 30 | 536 | 17.9 | -0.22 | -1.2 | 1.27 |
| **West North Central** | 41 | 497 | 12.1 | -0.03 | -1.2 | 1.27 |
| **West South Central** | 38 | 559 | 14.7 | 0.33 | -1.2 | 1.27 |
| **Model implementation evaluation 2024 score** | | | | | | |
| **Division** | **Clusters (n)** | **Total hospitals (n)** | **Average cluster size** | **Average score** | **Minimum Score** | **Maximum score** |
| **TOTAL** | 255 | 2323 | 9.1 | -0.14 | -1.27 | 1.45 |
| **East North Central** | 41 | 343 | 8.4 | -0.3 | -1.27 | 1.45 |
| **East South Central** | 22 | 123 | 5.6 | -0.11 | -1.27 | 1.45 |
| **Mid Atlantic** | 26 | 205 | 7.9 | -0.1 | -1.27 | 1.45 |
| **Mountain** | 33 | 170 | 5.2 | -0.37 | -1.27 | 1.45 |
| **New England** | 19 | 55 | 2.9 | 0.17 | -1.27 | 1.45 |
| **Pacific** | 9 | 242 | 26.9 | -0.11 | -1.27 | 1.34 |
| **South Atlantic** | 28 | 393 | 14 | 0.24 | -1.27 | 1.45 |
| **West North Central** | 37 | 401 | 10.8 | -0.08 | -1.27 | 1.34 |
| **West South Central** | 40 | 391 | 9.8 | -0.32 | -1.27 | 1.45 |
| **LLM readiness score** | | | | | | |
| **Division** | **Clusters (n)** | **Total hospitals (n)** | **Average cluster size** | **Average score** | **Minimum Score** | **Maximum score** |
| **TOTAL** | 252 | 1951 | 7.7 | -0.23 | -1.35 | 1.05 |
| **East North Central** | 43 | 320 | 7.4 | -0.31 | -1.35 | 1.05 |
| **East South Central** | 22 | 105 | 4.8 | 0.04 | -1.35 | 1.05 |
| **Mid Atlantic** | 26 | 186 | 7.2 | -0.24 | -1.35 | 1.05 |
| **Mountain** | 23 | 116 | 5 | -0.48 | -1.35 | 1.05 |
| **New England** | 18 | 55 | 3.1 | 0.03 | -1.35 | 1.05 |
| **Pacific** | 11 | 228 | 20.7 | -0.09 | -1.35 | 1.05 |
| **South Atlantic** | 24 | 370 | 15.4 | -0.18 | -1.35 | 1.05 |
| **West North Central** | 34 | 228 | 6.7 | -0.26 | -1.35 | 1.05 |
| **West South Central** | 51 | 343 | 6.7 | -0.28 | -1.35 | 1.05 |

**Table S11.** Hotspot/coldspot results across U.S. hospitals and hospital service areas

Geospatial clusters of AI implementation were identified using Getis-Ord Gi* statistics. Results are shown for hospitals and hospital service areas, with significance determined after multiple testing correction using Benjamini–Hochberg false discovery rate (FDR) and Storey’s q-value methods

|  |  | **Hotspot** | | | **Coldspot** | | |  |
| --- | --- | --- | --- | --- | --- | --- | --- | --- |
|  | **Adjustment method** | **90%** | **95%** | **99%** | **90%** | **95%** | **99%** | **Not significant** |
| **AI implementation base score** | | | | | | | | |
| **Hospital** | Unadjusted | 357 | 300 | 0 | 254 | 232 | 86 | 2331 |
|  | Storey | 208 | 0 | 0 | 164 | 40 | 0 | 3148 |
|  | BH | 0 | 0 | 0 | 40 | 0 | 0 | 3520 |
| **HSA** | Unadjusted | 219 | 159 | 0 | 171 | 162 | 75 | 1405 |
|  | Storey | 117 | 0 | 0 | 126 | 37 | 0 | 1911 |
|  | BH | 0 | 0 | 0 | 37 | 0 | 0 | 2154 |
| **Model implementation breadth score** | | | | | | | | |
| **Hospital** | Unadjusted | 149 | 250 | 60 | 136 | 205 | 130 | 2628 |
|  | Storey | 143 | 9 | 0 | 165 | 40 | 0 | 3201 |
|  | BH | 9 | 0 | 0 | 40 | 0 | 0 | 3509 |
| **HSA** | Unadjusted | 91 | 128 | 34 | 94 | 129 | 110 | 1605 |
|  | Storey | 78 | 5 | 0 | 126 | 37 | 0 | 1945 |
|  | BH | 5 | 0 | 0 | 37 | 0 | 0 | 2149 |
| **Model implementation development score** | | | | | | | | |
| **Hospital** | Unadjusted | 194 | 173 | 129 | 167 | 192 | 141 | 2564 |
|  | Storey | 140 | 57 | 0 | 162 | 48 | 0 | 3153 |
|  | BH | 45 | 33 | 0 | 58 | 0 | 0 | 3424 |
| **HSA** | Unadjusted | 112 | 103 | 70 | 117 | 125 | 119 | 1545 |
|  | Storey | 76 | 33 | 0 | 115 | 45 | 0 | 1922 |
|  | BH | 32 | 14 | 0 | 53 | 0 | 0 | 2092 |
| **Model implementation evaluation 2023 score** | | | | | | | | |
| **Hospital** | Unadjusted | 178 | 234 | 95 | 150 | 192 | 133 | 2578 |
|  | Storey | 160 | 0 | 0 | 164 | 0 | 0 | 3236 |
|  | BH | 28 | 0 | 0 | 40 | 0 | 0 | 3492 |
| **HSA** | Unadjusted | 102 | 125 | 48 | 97 | 136 | 111 | 1572 |
|  | Storey | 83 | 0 | 0 | 133 | 0 | 0 | 1975 |
|  | BH | 13 | 0 | 0 | 37 | 0 | 0 | 2141 |
| **Model implementation evaluation 2024 score** | | | | | | | | |
| **Hospital** | Unadjusted | 146 | 153 | 100 | 85 | 205 | 110 | 1824 |
|  | Storey | 133 | 74 | 0 | 190 | 61 | 0 | 2165 |
|  | BH | 77 | 0 | 0 | 67 | 0 | 0 | 2479 |
| **HSA** | Unadjusted | 91 | 100 | 55 | 55 | 134 | 91 | 1665 |
|  | Storey | 83 | 37 | 0 | 132 | 52 | 0 | 1887 |
|  | BH | 40 | 0 | 0 | 58 | 0 | 0 | 2093 |
| **LLM readiness score** | | | | | | | | |
| **Hospital** | Unadjusted | 116 | 90 | 80 | 95 | 123 | 117 | 1612 |
|  | Storey | 90 | 80 | 0 | 61 | 117 | 0 | 1885 |
|  | BH | 80 | 0 | 0 | 95 | 22 | 0 | 2036 |
| **HSA** | Unadjusted | 72 | 58 | 54 | 73 | 89 | 89 | 1756 |
|  | Storey | 58 | 54 | 0 | 38 | 89 | 0 | 1952 |
|  | BH | 54 | 0 | 0 | 70 | 19 | 0 | 2048 |

**Table S12.** Hotspot/coldspot results for AI implementation base score by census division and state

Census division abbreviations: NE=New England, MA=Mid Atlantic, SA=South Atlantic, ENC=East North Central, WNC=West North Central, ESC=East South Central, WSC=West South Central, MTN=Mountain, PAC=Pacific. State abbreviations: AL=Alabama, AK=Alaska, AZ=Arizona, AR=Arkansas, CA=California, CO=Colorado, CT=Connecticut, DE=Delaware, FL=Florida, GA=Georgia, HI=Hawaii, ID=Idaho, IL=Illinois, IN=Indiana, IA=Iowa, KS=Kansas, KY=Kentucky, LA=Louisiana, ME=Maine, MD=Maryland, MA=Massachusetts, MI=Michigan, MN=Minnesota, MS=Mississippi, MO=Missouri, MT=Montana, NE=Nebraska, NV=Nevada, NH=New Hampshire, NJ=New Jersey, NM=New Mexico, NY=New York, NC=North Carolina, ND=North Dakota, OH=Ohio, OK=Oklahoma, OR=Oregon, PA=Pennsylvania, RI=Rhode Island, SC=South Carolina, SD=South Dakota, TN=Tennessee, TX=Texas, UT=Utah, VT=Vermont, VA=Virginia, WA=Washington, WV=West Virginia, WI=Wisconsin, WY=Wyoming. States with less than 20 hospitals were suppressed.

| **AI implementation base score hotspot analysis** | | | | | |
| --- | --- | --- | --- | --- | --- |
|  | **Hotspot %** | **Coldspot %** | **Net %** | **Total** | **Mean Z** |
| **Census division** |  |  |  |  |  |
| SA | 29.6 | 3 | 26.5 | 565 | 0.72 |
| ENC | 27.8 | 7.4 | 20.3 | 497 | 0.52 |
| MA | 18.5 | 10.7 | 7.8 | 335 | 0.23 |
| MNT | 21.9 | 19.6 | 2.3 | 306 | 0.15 |
| NE | 9.4 | 15.1 | -5.8 | 139 | 0.13 |
| PAC | 14.2 | 9.3 | 4.9 | 367 | 0.06 |
| ESC | 9.5 | 11.7 | -2.3 | 222 | -0.03 |
| WNC | 19.6 | 26.4 | -6.8 | 541 | -0.2 |
| WSC | 5.4 | 33.3 | -27.9 | 588 | -0.72 |
| **State** |  |  |  |  |  |
| SC | 70.2 | 0 | 70.2 | 47 | 1.78 |
| SD | 68.2 | 0 | 68.2 | 44 | 1.67 |
| MN | 57.1 | 0 | 57.1 | 91 | 1.48 |
| OH | 45 | 0 | 45 | 120 | 1.21 |
| CT | 23.3 | 0 | 23.3 | 30 | 1.19 |
| AZ | 59.6 | 15.8 | 43.9 | 57 | 1.1 |
| VA | 37.3 | 0 | 37.3 | 75 | 1.07 |
| NC | 42.7 | 2.2 | 40.4 | 89 | 1.03 |
| NJ | 30.3 | 1.5 | 28.8 | 66 | 0.77 |
| UT | 9.1 | 0 | 9.1 | 44 | 0.74 |
| MD | 16.7 | 0 | 16.7 | 48 | 0.67 |
| NV | 10.3 | 0 | 10.3 | 29 | 0.6 |
| WI | 44.9 | 9 | 35.9 | 78 | 0.58 |
| FL | 16.9 | 0 | 16.9 | 154 | 0.56 |
| CO | 33.8 | 9.5 | 24.3 | 74 | 0.53 |
| PA | 28.5 | 8.8 | 19.7 | 137 | 0.51 |
| IL | 23.4 | 8.8 | 14.6 | 137 | 0.39 |
| ND | 22.2 | 7.4 | 14.8 | 27 | 0.39 |
| KY | 17 | 13.2 | 3.8 | 53 | 0.35 |
| MA | 2 | 0 | 2 | 49 | 0.34 |
| GA | 28.7 | 13.8 | 14.9 | 94 | 0.29 |
| IN | 13.5 | 8.1 | 5.4 | 74 | 0.25 |
| MS | 11.3 | 5.7 | 5.7 | 53 | 0.2 |
| WV | 17.1 | 4.9 | 12.2 | 41 | 0.2 |
| CA | 14.5 | 7.9 | 6.6 | 242 | 0.14 |
| WA | 18.5 | 14.8 | 3.7 | 54 | 0.03 |
| MI | 8 | 13.6 | -5.7 | 88 | -0.07 |
| AR | 14.3 | 20 | -5.7 | 70 | -0.09 |
| OR | 16.3 | 16.3 | 0 | 43 | -0.19 |
| NE | 15.1 | 15.1 | 0 | 53 | -0.22 |
| TN | 1.4 | 8.6 | -7.1 | 70 | -0.3 |
| AL | 10.9 | 21.7 | -10.9 | 46 | -0.31 |
| NY | 2.3 | 17.4 | -15.2 | 132 | -0.32 |
| LA | 2.3 | 31.8 | -29.5 | 88 | -0.62 |
| NM | 0 | 33.3 | -33.3 | 24 | -0.66 |
| MO | 0.8 | 26.5 | -25.8 | 132 | -0.66 |
| TX | 4.5 | 33.9 | -29.4 | 354 | -0.79 |
| OK | 5.3 | 44.7 | -39.5 | 76 | -1.07 |
| IA | 7.3 | 46.8 | -39.4 | 109 | -1.11 |
| ID | 0 | 29.6 | -29.6 | 27 | -1.26 |
| KS | 1.2 | 55.3 | -54.1 | 85 | -1.29 |
| NH | 0 | 60.9 | -60.9 | 23 | -1.47 |
| MT | 0 | 69.4 | -69.4 | 36 | -1.53 |

**Table S13.** Hotspot/coldspot results for Model implementation breadth score by census division and state

Census division abbreviations: NE=New England, MA=Mid Atlantic, SA=South Atlantic, ENC=East North Central, WNC=West North Central, ESC=East South Central, WSC=West South Central, MTN=Mountain, PAC=Pacific. State abbreviations: AL=Alabama, AK=Alaska, AZ=Arizona, AR=Arkansas, CA=California, CO=Colorado, CT=Connecticut, DE=Delaware, FL=Florida, GA=Georgia, HI=Hawaii, ID=Idaho, IL=Illinois, IN=Indiana, IA=Iowa, KS=Kansas, KY=Kentucky, LA=Louisiana, ME=Maine, MD=Maryland, MA=Massachusetts, MI=Michigan, MN=Minnesota, MS=Mississippi, MO=Missouri, MT=Montana, NE=Nebraska, NV=Nevada, NH=New Hampshire, NJ=New Jersey, NM=New Mexico, NY=New York, NC=North Carolina, ND=North Dakota, OH=Ohio, OK=Oklahoma, OR=Oregon, PA=Pennsylvania, RI=Rhode Island, SC=South Carolina, SD=South Dakota, TN=Tennessee, TX=Texas, UT=Utah, VT=Vermont, VA=Virginia, WA=Washington, WV=West Virginia, WI=Wisconsin, WY=Wyoming. States with less than 20 hospitals were suppressed.

| **Model implementation breadth score hotspot analysis** | | | | | |
| --- | --- | --- | --- | --- | --- |
|  | **Hotspot %** | **Coldspot %** | **Net %** | **Total** | **Mean Z** |
| **Census division** |  |  |  |  |  |
| SA | 25.3 | 2.1 | 23.2 | 565 | 0.85 |
| ENC | 19.7 | 5.8 | 13.9 | 497 | 0.57 |
| MA | 11.4 | 7.2 | 4.2 | 334 | 0.28 |
| PAC | 12.8 | 6.5 | 6.3 | 367 | 0.25 |
| MNT | 19 | 17.4 | 1.6 | 305 | 0.1 |
| ESC | 4.1 | 11.3 | -7.2 | 222 | -0.12 |
| NE | 0 | 12.9 | -12.9 | 139 | -0.24 |
| WNC | 8.9 | 23.5 | -14.6 | 541 | -0.34 |
| **State** |  |  |  |  |  |
| NC | 41.6 | 1.1 | 40.4 | 89 | 1.48 |
| MN | 35.2 | 0 | 35.2 | 91 | 1.35 |
| SC | 48.9 | 0 | 48.9 | 47 | 1.29 |
| VA | 36 | 0 | 36 | 75 | 1.24 |
| SD | 15.9 | 0 | 15.9 | 44 | 1.05 |
| OH | 20 | 0 | 20 | 120 | 0.96 |
| NJ | 24.2 | 1.5 | 22.7 | 66 | 0.94 |
| FL | 20.8 | 0 | 20.8 | 154 | 0.93 |
| AZ | 57.9 | 15.8 | 42.1 | 57 | 0.84 |
| UT | 6.8 | 2.3 | 4.5 | 44 | 0.75 |
| MD | 4.2 | 0 | 4.2 | 48 | 0.67 |
| CT | 0 | 0 | 0 | 30 | 0.63 |
| CO | 26 | 9.6 | 16.4 | 73 | 0.58 |
| WI | 29.5 | 9 | 20.5 | 78 | 0.55 |
| PA | 14 | 1.5 | 12.5 | 136 | 0.52 |
| IL | 21.9 | 5.8 | 16.1 | 137 | 0.45 |
| ND | 18.5 | 3.7 | 14.8 | 27 | 0.43 |
| NV | 6.9 | 0 | 6.9 | 29 | 0.43 |
| MI | 13.6 | 10.2 | 3.4 | 88 | 0.4 |
| IN | 12.2 | 6.8 | 5.4 | 74 | 0.4 |
| CA | 14.5 | 5.8 | 8.7 | 242 | 0.34 |
| KY | 15.1 | 11.3 | 3.8 | 53 | 0.31 |
| WA | 18.5 | 7.4 | 11.1 | 54 | 0.26 |
| GA | 20.2 | 10.6 | 9.6 | 94 | 0.24 |
| MA | 0 | 0 | 0 | 49 | 0.03 |
| WV | 7.3 | 2.4 | 4.9 | 41 | -0.01 |
| OR | 4.7 | 14 | -9.3 | 43 | -0.04 |
| MS | 1.9 | 5.7 | -3.8 | 53 | -0.08 |
| AR | 8.6 | 24.3 | -15.7 | 70 | -0.19 |
| TN | 0 | 8.6 | -8.6 | 70 | -0.23 |
| NY | 2.3 | 15.9 | -13.6 | 132 | -0.31 |
| NE | 3.8 | 11.3 | -7.5 | 53 | -0.42 |
| AL | 0 | 21.7 | -21.7 | 46 | -0.47 |
| NM | 0 | 33.3 | -33.3 | 24 | -0.51 |
| MO | 0 | 22.7 | -22.7 | 132 | -0.66 |
| TX | 2.8 | 27.7 | -24.9 | 354 | -0.82 |
| LA | 0 | 21.6 | -21.6 | 88 | -0.91 |
| OK | 1.3 | 32.9 | -31.6 | 76 | -0.97 |
| ID | 0 | 18.5 | -18.5 | 27 | -1.19 |
| KS | 0 | 44.7 | -44.7 | 85 | -1.31 |
| IA | 1.8 | 47.7 | -45.9 | 109 | -1.34 |
| NH | 0 | 47.8 | -47.8 | 23 | -1.43 |
| MT | 0 | 55.6 | -55.6 | 36 | -1.58 |

**Table S14.** Hotspot/coldspot results for model implementation development score by census division and state

Census division abbreviations: NE=New England, MA=Mid Atlantic, SA=South Atlantic, ENC=East North Central, WNC=West North Central, ESC=East South Central, WSC=West South Central, MTN=Mountain, PAC=Pacific. State abbreviations: AL=Alabama, AK=Alaska, AZ=Arizona, AR=Arkansas, CA=California, CO=Colorado, CT=Connecticut, DE=Delaware, FL=Florida, GA=Georgia, HI=Hawaii, ID=Idaho, IL=Illinois, IN=Indiana, IA=Iowa, KS=Kansas, KY=Kentucky, LA=Louisiana, ME=Maine, MD=Maryland, MA=Massachusetts, MI=Michigan, MN=Minnesota, MS=Mississippi, MO=Missouri, MT=Montana, NE=Nebraska, NV=Nevada, NH=New Hampshire, NJ=New Jersey, NM=New Mexico, NY=New York, NC=North Carolina, ND=North Dakota, OH=Ohio, OK=Oklahoma, OR=Oregon, PA=Pennsylvania, RI=Rhode Island, SC=South Carolina, SD=South Dakota, TN=Tennessee, TX=Texas, UT=Utah, VT=Vermont, VA=Virginia, WA=Washington, WV=West Virginia, WI=Wisconsin, WY=Wyoming; States with less than 20 hospitals were suppressed.

| **AI implementation development score hotspot analysis** | | | | | |
| --- | --- | --- | --- | --- | --- |
|  | **Hotspot %** | **Coldspot %** | **Net %** | **Total** | **Mean Z** |
| **Census division** |  |  |  |  |  |
| SA | 21.9 | 2.3 | 19.6 | 565 | 0.75 |
| PAC | 24.5 | 4.6 | 19.9 | 367 | 0.68 |
| ENC | 21.1 | 5.6 | 15.5 | 497 | 0.56 |
| MA | 23 | 7.5 | 15.5 | 335 | 0.29 |
| MNT | 15.4 | 12.4 | 2.9 | 306 | 0.03 |
| ESC | 4.1 | 10.8 | -6.8 | 222 | -0.15 |
| NE | 0 | 10.8 | -10.8 | 139 | -0.31 |
| WNC | 11.8 | 23.1 | -11.3 | 541 | -0.32 |
| WSC | 1.7 | 27.2 | -25.5 | 588 | -0.82 |
| **State** |  |  |  |  |  |
| NC | 52.8 | 1.1 | 51.7 | 89 | 1.94 |
| UT | 52.3 | 0 | 52.3 | 44 | 1.6 |
| MN | 41.8 | 0 | 41.8 | 91 | 1.5 |
| NJ | 39.4 | 1.5 | 37.9 | 66 | 1.21 |
| SC | 36.2 | 0 | 36.2 | 47 | 1.06 |
| VA | 10.7 | 0 | 10.7 | 75 | 0.95 |
| OH | 19.2 | 0 | 19.2 | 120 | 0.85 |
| CA | 30.2 | 2.9 | 27.3 | 242 | 0.84 |
| PA | 37.2 | 2.9 | 34.3 | 137 | 0.82 |
| ND | 29.6 | 0 | 29.6 | 27 | 0.73 |
| SD | 6.8 | 0 | 6.8 | 44 | 0.67 |
| MD | 18.8 | 0 | 18.8 | 48 | 0.63 |
| FL | 14.9 | 0.6 | 14.3 | 154 | 0.61 |
| IL | 25.5 | 5.1 | 20.4 | 137 | 0.58 |
| WA | 20.4 | 7.4 | 13 | 54 | 0.58 |
| WI | 24.4 | 10.3 | 14.1 | 78 | 0.5 |
| IN | 21.6 | 5.4 | 16.2 | 74 | 0.49 |
| CO | 23 | 6.8 | 16.2 | 74 | 0.44 |
| CT | 0 | 0 | 0 | 30 | 0.4 |
| OR | 7 | 14 | -7 | 43 | 0.34 |
| MI | 13.6 | 10.2 | 3.4 | 88 | 0.26 |
| AZ | 5.3 | 15.8 | -10.5 | 57 | 0.26 |
| KY | 7.5 | 11.3 | -3.8 | 53 | 0.22 |
| GA | 19.1 | 10.6 | 8.5 | 94 | 0.12 |
| WV | 4.9 | 2.4 | 2.4 | 41 | -0.07 |
| NV | 10.3 | 0 | 10.3 | 29 | -0.11 |
| TN | 1.4 | 8.6 | -7.1 | 70 | -0.14 |
| MS | 0 | 3.8 | -3.8 | 53 | -0.16 |
| MA | 0 | 2 | -2 | 49 | -0.18 |
| AR | 8.6 | 15.7 | -7.1 | 70 | -0.25 |
| NE | 13.2 | 13.2 | 0 | 53 | -0.46 |
| MO | 4.5 | 25.8 | -21.2 | 132 | -0.49 |
| LA | 2.3 | 25 | -22.7 | 88 | -0.57 |
| AL | 8.7 | 21.7 | -13 | 46 | -0.59 |
| NM | 0 | 16.7 | -16.7 | 24 | -0.6 |
| NY | 0 | 15.2 | -15.2 | 132 | -0.72 |
| TX | 0.3 | 29.4 | -29.1 | 354 | -0.96 |
| OK | 1.3 | 30.3 | -28.9 | 76 | -0.97 |
| ID | 0 | 22.2 | -22.2 | 27 | -1.17 |
| KS | 0 | 38.8 | -38.8 | 85 | -1.26 |
| MT | 0 | 30.6 | -30.6 | 36 | -1.31 |
| NH | 0 | 34.8 | -34.8 | 23 | -1.38 |
| IA | 1.8 | 46.8 | -45.0 | 109 | -1.51 |

**Table S15.** Hotspot/coldspot results for model implementation evaluation score by census division and state

Census division abbreviations: NE=New England, MA=Mid Atlantic, SA=South Atlantic, ENC=East North Central, WNC=West North Central, ESC=East South Central, WSC=West South Central, MTN=Mountain, PAC=Pacific. State abbreviations: AL=Alabama, AK=Alaska, AZ=Arizona, AR=Arkansas, CA=California, CO=Colorado, CT=Connecticut, DE=Delaware, FL=Florida, GA=Georgia, HI=Hawaii, ID=Idaho, IL=Illinois, IN=Indiana, IA=Iowa, KS=Kansas, KY=Kentucky, LA=Louisiana, ME=Maine, MD=Maryland, MA=Massachusetts, MI=Michigan, MN=Minnesota, MS=Mississippi, MO=Missouri, MT=Montana, NE=Nebraska, NV=Nevada, NH=New Hampshire, NJ=New Jersey, NM=New Mexico, NY=New York, NC=North Carolina, ND=North Dakota, OH=Ohio, OK=Oklahoma, OR=Oregon, PA=Pennsylvania, RI=Rhode Island, SC=South Carolina, SD=South Dakota, TN=Tennessee, TX=Texas, UT=Utah, VT=Vermont, VA=Virginia, WA=Washington, WV=West Virginia, WI=Wisconsin, WY=Wyoming; States with less than 20 hospitals were suppressed.

| **AI implementation evaluation 2023 score hotspot analysis** | | | | | |
| --- | --- | --- | --- | --- | --- |
|  | **Hotspot %** | **Coldspot %** | **Net %** | **Total** | **Mean Z** |
| **Census division** |  |  |  |  |  |
| SA | 24.4 | 2.5 | 21.9 | 565 | 0.86 |
| MA | 16.4 | 6.9 | 9.6 | 335 | 0.41 |
| ENC | 17.9 | 6.8 | 11.1 | 497 | 0.35 |
| MNT | 21.9 | 18 | 3.9 | 306 | 0.21 |
| ESC | 14 | 6.8 | 7.2 | 222 | 0.17 |
| PAC | 9.8 | 7.6 | 2.2 | 367 | 0.07 |
| NE | 4.3 | 14.4 | -10.1 | 139 | -0.14 |
| WNC | 10.2 | 23.7 | -13.5 | 541 | -0.36 |
| WSC | 4.8 | 26.4 | -21.6 | 588 | -0.66 |
| **State** |  |  |  |  |  |
| SC | 61.7 | 0 | 61.7 | 47 | 1.98 |
| UT | 54.5 | 0 | 54.5 | 44 | 1.49 |
| NC | 46.1 | 1.1 | 44.9 | 89 | 1.43 |
| MN | 29.7 | 0 | 29.7 | 91 | 1.31 |
| CT | 13.3 | 0 | 13.3 | 30 | 1.09 |
| NJ | 25.8 | 1.5 | 24.2 | 66 | 0.95 |
| ND | 29.6 | 0 | 29.6 | 27 | 0.92 |
| FL | 20.1 | 1.9 | 18.2 | 154 | 0.9 |
| SD | 22.7 | 0 | 22.7 | 44 | 0.84 |
| VA | 5.3 | 0 | 5.3 | 75 | 0.78 |
| CO | 40.5 | 8.1 | 32.4 | 74 | 0.76 |
| OH | 27.5 | 0.8 | 26.7 | 120 | 0.68 |
| PA | 24.1 | 2.9 | 21.2 | 137 | 0.66 |
| KY | 22.6 | 11.3 | 11.3 | 53 | 0.6 |
| NV | 13.8 | 0 | 13.8 | 29 | 0.51 |
| WV | 12.2 | 0 | 12.2 | 41 | 0.49 |
| MS | 15.1 | 3.8 | 11.3 | 53 | 0.46 |
| AZ | 12.3 | 15.8 | -3.5 | 57 | 0.43 |
| GA | 25.5 | 9.6 | 16 | 94 | 0.37 |
| MD | 8.3 | 2.1 | 6.2 | 48 | 0.35 |
| WI | 25.6 | 9 | 16.7 | 78 | 0.35 |
| IL | 17.5 | 8 | 9.5 | 137 | 0.34 |
| IN | 8.1 | 6.8 | 1.4 | 74 | 0.28 |
| MA | 0 | 0 | 0 | 49 | 0.21 |
| CA | 12 | 6.2 | 5.8 | 242 | 0.2 |
| WA | 13 | 14.8 | -1.9 | 54 | 0.02 |
| MI | 6.8 | 11.4 | -4.5 | 88 | -0.02 |
| TN | 14.3 | 1.4 | 12.9 | 70 | -0.03 |
| NY | 3.8 | 13.6 | -9.8 | 132 | -0.12 |
| NE | 15.1 | 13.2 | 1.9 | 53 | -0.23 |
| AL | 2.2 | 13 | -10.9 | 46 | -0.34 |
| OR | 0 | 11.6 | -11.6 | 43 | -0.38 |
| ID | 0 | 18.5 | -18.5 | 27 | -0.4 |
| AR | 5.7 | 18.6 | -12.9 | 70 | -0.41 |
| LA | 2.3 | 14.8 | -12.5 | 88 | -0.51 |
| TX | 5.9 | 28 | -22 | 354 | -0.61 |
| NM | 0 | 33.3 | -33.3 | 24 | -0.63 |
| MO | 0 | 25.8 | -25.8 | 132 | -0.86 |
| KS | 0 | 45.9 | -45.9 | 85 | -1.28 |
| IA | 1.8 | 44 | -42.2 | 109 | -1.28 |
| OK | 1.3 | 39.5 | -38.2 | 76 | -1.3 |
| NH | 0 | 47.8 | -47.8 | 23 | -1.47 |
| MT | 0 | 61.1 | -61.1 | 36 | -1.7 |

**Table S16.** Hotspot/coldspot results for model implementation evaluation 2024 score by census division and state

Census division abbreviations: NE=New England, MA=Mid Atlantic, SA=South Atlantic, ENC=East North Central, WNC=West North Central, ESC=East South Central, WSC=West South Central, MTN=Mountain, PAC=Pacific. State abbreviations: AL=Alabama, AK=Alaska, AZ=Arizona, AR=Arkansas, CA=California, CO=Colorado, CT=Connecticut, DE=Delaware, FL=Florida, GA=Georgia, HI=Hawaii, ID=Idaho, IL=Illinois, IN=Indiana, IA=Iowa, KS=Kansas, KY=Kentucky, LA=Louisiana, ME=Maine, MD=Maryland, MA=Massachusetts, MI=Michigan, MN=Minnesota, MS=Mississippi, MO=Missouri, MT=Montana, NE=Nebraska, NV=Nevada, NH=New Hampshire, NJ=New Jersey, NM=New Mexico, NY=New York, NC=North Carolina, ND=North Dakota, OH=Ohio, OK=Oklahoma, OR=Oregon, PA=Pennsylvania, RI=Rhode Island, SC=South Carolina, SD=South Dakota, TN=Tennessee, TX=Texas, UT=Utah, VT=Vermont, VA=Virginia, WA=Washington, WV=West Virginia, WI=Wisconsin, WY=Wyoming; States with less than 20 hospitals were suppressed.

| **AI implementation evaluation 2024 score hotspot analysis** | | | | | |
| --- | --- | --- | --- | --- | --- |
|  | **Hotspot %** | **Coldspot %** | **Net %** | **Total** | **Mean Z** |
| **Census division** |  |  |  |  |  |
| SA | 33.3 | 0 | 33.3 | 415 | 1.19 |
| ESC | 17.7 | 10.1 | 7.6 | 158 | 0.46 |
| MA | 19.4 | 8.3 | 11.2 | 242 | 0.45 |
| ENC | 13.8 | 10.9 | 2.9 | 385 | 0.28 |
| PAC | 9.9 | 8.3 | 1.6 | 253 | 0.16 |
| MNT | 18.2 | 18.7 | -0.5 | 203 | -0.01 |
| NE | 10.1 | 21.3 | -11.2 | 89 | -0.11 |
| WSC | 5.6 | 25.9 | -20.3 | 428 | -0.67 |
| WNC | 8.7 | 29.3 | -20.7 | 450 | -0.68 |
| **State** |  |  |  |  |  |
| NC | 55.6 | 0 | 55.6 | 63 | 1.75 |
| SC | 56.8 | 0 | 56.8 | 37 | 1.67 |
| CT | 32 | 0 | 32 | 25 | 1.38 |
| WA | 60.9 | 21.7 | 39.1 | 23 | 1.22 |
| MS | 24.4 | 0 | 24.4 | 41 | 1.16 |
| FL | 23.3 | 0 | 23.3 | 116 | 1.13 |
| ND | 37.5 | 0 | 37.5 | 24 | 1.11 |
| VA | 37.3 | 0 | 37.3 | 51 | 1.1 |
| PA | 43.2 | 4.1 | 39.2 | 74 | 1.04 |
| MD | 19.4 | 0 | 19.4 | 36 | 1.03 |
| NJ | 23.7 | 1.7 | 22 | 59 | 0.95 |
| MN | 23.3 | 1.4 | 21.9 | 73 | 0.94 |
| WV | 25.8 | 0 | 25.8 | 31 | 0.94 |
| GA | 29.6 | 0 | 29.6 | 71 | 0.88 |
| CO | 37.9 | 3.4 | 34.5 | 58 | 0.85 |
| OH | 30.9 | 2.1 | 28.9 | 97 | 0.7 |
| KY | 22.5 | 10 | 12.5 | 40 | 0.67 |
| AZ | 23.7 | 10.5 | 13.2 | 38 | 0.45 |
| WI | 6.7 | 15 | -8.3 | 60 | 0.22 |
| IN | 3.7 | 13 | -9.3 | 54 | 0.22 |
| TN | 14 | 8 | 6 | 50 | 0.17 |
| CA | 5.8 | 5.3 | 0.5 | 189 | 0.16 |
| SD | 8.8 | 0 | 8.8 | 34 | 0.11 |
| IL | 9.8 | 12.5 | -2.7 | 112 | 0.09 |
| MI | 9.7 | 16.1 | -6.5 | 62 | 0.08 |
| NE | 17.1 | 2.4 | 14.6 | 41 | 0.08 |
| LA | 16.9 | 18.6 | -1.7 | 59 | -0.03 |
| MA | 0 | 7.7 | -7.7 | 26 | -0.1 |
| NY | 0.9 | 14.7 | -13.8 | 109 | -0.22 |
| AL | 7.4 | 29.6 | -22.2 | 27 | -0.36 |
| OR | 0 | 27.3 | -27.3 | 22 | -0.54 |
| AR | 1.9 | 17 | -15.1 | 53 | -0.69 |
| TX | 4.9 | 26.3 | -21.4 | 266 | -0.71 |
| OK | 0 | 42 | -42 | 50 | -1.18 |
| KS | 0 | 44.6 | -44.6 | 65 | -1.28 |
| IA | 3.4 | 50.6 | -47.2 | 89 | -1.52 |
| MO | 0 | 45.2 | -45.2 | 124 | -1.52 |
| MT | 0 | 87 | -87 | 23 | -2.36 |

**Table S17.** Hotspot/coldspot results for LLM readiness score by census division and state.

Census division abbreviations: NE=New England, MA=Mid Atlantic, SA=South Atlantic, ENC=East North Central, WNC=West North Central, ESC=East South Central, WSC=West South Central, MTN=Mountain, PAC=Pacific. State abbreviations: AL=Alabama, AK=Alaska, AZ=Arizona, AR=Arkansas, CA=California, CO=Colorado, CT=Connecticut, DE=Delaware, FL=Florida, GA=Georgia, HI=Hawaii, ID=Idaho, IL=Illinois, IN=Indiana, IA=Iowa, KS=Kansas, KY=Kentucky, LA=Louisiana, ME=Maine, MD=Maryland, MA=Massachusetts, MI=Michigan, MN=Minnesota, MS=Mississippi, MO=Missouri, MT=Montana, NE=Nebraska, NV=Nevada, NH=New Hampshire, NJ=New Jersey, NM=New Mexico, NY=New York, NC=North Carolina, ND=North Dakota, OH=Ohio, OK=Oklahoma, OR=Oregon, PA=Pennsylvania, RI=Rhode Island, SC=South Carolina, SD=South Dakota, TN=Tennessee, TX=Texas, UT=Utah, VT=Vermont, VA=Virginia, WA=Washington, WV=West Virginia, WI=Wisconsin, WY=Wyoming; States with less than 20 hospitals were suppressed.

| **LLM readiness score hotspot analysis** | | | | | |
| --- | --- | --- | --- | --- | --- |
|  | **Hotspot %** | **Coldspot %** | **Net %** | **Total** | **Mean Z** |
| **Census division** |  |  |  |  |  |
| SA | 22.9 | 1 | 21.9 | 397 | 1.04 |
| ENC | 20.2 | 4.6 | 15.7 | 351 | 0.69 |
| NE | 5.7 | 5.7 | 0 | 88 | 0.25 |
| ESC | 6.5 | 11.6 | -5.1 | 138 | 0.12 |
| MA | 14.3 | 14.7 | -0.4 | 224 | -0.17 |
| WNC | 12 | 27.2 | -15.2 | 283 | -0.22 |
| PAC | 9.4 | 18.3 | -8.9 | 235 | -0.23 |
| WSC | 5.8 | 27.5 | -21.7 | 364 | -0.67 |
| MNT | 1.3 | 24.8 | -23.5 | 153 | -0.68 |
| **State** |  |  |  |  |  |
| NC | 53.6 | 0 | 53.6 | 56 | 1.8 |
| SD | 58.8 | 0 | 58.8 | 34 | 1.78 |
| VA | 48 | 0 | 48 | 50 | 1.74 |
| SC | 47.2 | 0 | 47.2 | 36 | 1.65 |
| WI | 56.1 | 7 | 49.1 | 57 | 1.29 |
| IN | 14.6 | 0 | 14.6 | 48 | 0.94 |
| PA | 43.9 | 4.5 | 39.4 | 66 | 0.94 |
| MD | 0 | 0 | 0 | 35 | 0.86 |
| FL | 8.7 | 0 | 8.7 | 115 | 0.78 |
| MA | 7.7 | 0 | 7.7 | 26 | 0.77 |
| GA | 13.4 | 1.5 | 11.9 | 67 | 0.72 |
| IL | 20 | 8 | 12 | 100 | 0.61 |
| MS | 9.4 | 3.1 | 6.2 | 32 | 0.57 |
| MI | 0 | 1.9 | -1.9 | 52 | 0.53 |
| CT | 8 | 0 | 8 | 25 | 0.48 |
| TN | 0 | 0 | 0 | 49 | 0.38 |
| OH | 12.8 | 3.2 | 9.6 | 94 | 0.36 |
| MN | 9.7 | 12.5 | -2.8 | 72 | 0.32 |
| LA | 17.6 | 21.6 | -3.9 | 51 | 0.08 |
| CO | 0 | 8.3 | -8.3 | 48 | 0.05 |
| AL | 9.5 | 19 | -9.5 | 21 | -0.12 |
| CA | 12.4 | 16.3 | -3.9 | 178 | -0.13 |
| NJ | 0 | 3.7 | -3.7 | 54 | -0.24 |
| WV | 3.6 | 10.7 | -7.1 | 28 | -0.27 |
| AR | 2.1 | 8.5 | -6.4 | 47 | -0.33 |
| OK | 10.3 | 35.9 | -25.6 | 39 | -0.42 |
| OR | 0 | 25 | -25 | 20 | -0.46 |
| KY | 11.1 | 30.6 | -19.4 | 36 | -0.49 |
| IA | 10.6 | 33.3 | -22.7 | 66 | -0.5 |
| NY | 2.9 | 26.9 | -24 | 104 | -0.85 |
| NE | 0 | 32.4 | -32.4 | 37 | -0.89 |
| TX | 3.1 | 31.3 | -28.2 | 227 | -0.95 |
| ND | 0 | 40 | -40 | 20 | -1.07 |
| KS | 0 | 48.1 | -48.1 | 54 | -1.1 |
| WA | 0 | 42.9 | -42.9 | 21 | -1.23 |
| MT | 0 | 55 | -55 | 20 | -1.54 |

**Table S18.** Hospital quality metrics definition and selected covariates

Confounders were selected to avoid high-dimensional modeling: top predictors of AI adoption were included to adjust for systematic differences, and LASSO regression identified outcome-specific predictors. Mediators like interoperability were excluded.

| **Outcome** | **Outcome Description** | **Selected covariates** |
| --- | --- | --- |
| **Death_Complication_MORT_30_PN** | Death rate for pneumonia patients | 'non_profit_nongovernment',  'rural_urban_type',  'bedsize',  'national_adi_median',  'delivery_system',  'system_member',  'mean_primary_hpss' |
| **HAC_Reduction_Total HAC Score** | summary measure to evaluate hospital performance on hospital-acquired conditions | 'medicaid_ipd_percentage',  'Bedsize',  'Device_Percent',  'Delivery_system',  'system_member',  'for_profit',  'mean_primary_hpss' |
| **Readmission_READM-30-HF-HRRP** | Excess readmission ratio for heart failure patients | 'non_profit_nongovernment',  'bedsize',  'Internet_Percent',  'delivery_system',  'Broadband_Percent',  'system_member',  'teaching_hospital',  'for_profit' |
| **Readmission_READM-30-PN-HRRP** | Excess readmission ratio for pneumonia patients | 'medicaid_ipd_percentage',  'bedsize',  'Device_Percent',  'Internet_Percent',  'delivery_system',  ‘system_member',  'svi_theme3_median',  'medicare_ipd_percentage' |
| **Medicare_Spending_MSPB-1** | Spending per Hospital Patient with Medicare | 'non_profit_nongovernment',  'rural_urban_type',  'bedsize',  'Internet_Percent',  'delivery_system',  'federal_government',  'system_member' |
| **Timely_Care_HCP_COVID_19** | Percentage of healthcare personnel who are up to date with COVID-19 vaccinations | 'rural_urban_type',  'bedsize',  'delivery_system',  'mean_mua_score',  'System_member',  'Mean_mua_elders_score',  'teaching_hospital',  'mean_mental_hpss' |
| **Timely_Care_IMM_3** | Healthcare workers given influenza vaccination | 'non_profit_nongovernment',  'bedsize',  'national_adi_median',  'delivery_system',  'system_member',  'for_profit' |
| **Timely_Care_OP_18b** | Average (median) time patients spent in the emergency department before leaving from the visit | 'bedsize',  'national_adi_median',  'delivery_system',  'system_member',  'medicare_ipd_percentage',  'critical_access',  'for_profit' |
| **Timely_Care_OP_22** | Percentage of patients who left the emergency department before being seen | 'rural_urban_type',  'bedsize',  'delivery_system',  'system_member',  'Critical_access',  'for_profit',  'mean_primary_hpss' |
| **Timely_Care_SAFE_USE_OF_OPIOIDS** | Percentage of patients who were prescribed 2 or more opioids or an opioid and benzodiazepine concurrently at discharge | 'bedsize',  'delivery_system',  'mean_mua_score',  'system_member',  'medicare_ipd_percentage',  'teaching_hospital',  'critical_access' |
| **Timely_Care_SEP_1** | Severe Sepsis and Septic Shock | 'medicaid_ipd_percentage',  'bedsize',  'Device_Percent',  'delivery_system',  'System_member',  'for_profit' |
| **Timely_Care_SEV_SEP_3HR** | Septic Shock 3 Hour | 'medicaid_ipd_percentage',  'bedsize',  'delivery_system',  'system_member',  'medicare_ipd_percentage',  'critical_access',  'for_profit' |
| **Unplanned_Visits_EDAC_30_HF** | Hospital return days for heart failure patients | 'non_profit_nongovernment',  'medicaid_ipd_percentage',  'bedsize',  'delivery_system',  'mean_mua_score',  'system_member',  'teaching_hospital',  'mean_primary_hpss' |
| **Unplanned_Visits_EDAC_30_PN** | Hospital return days for pneumonia patients | 'Medicaid_ipd_percentage',  'bedsize',  'delivery_system',  'system_member',  'svi_theme3_median',  'medicare_ipd_percentage',  'community_hospital' |
| **Unplanned_Visits_OP_32** | Rate of unplanned hospital visits after an outpatient colonoscopy | 'bedsize',  'national_adi_median',  'delivery_system',  'system_member',  'mean_mua_infant_score',  'mean_mental_hpss',  'mean_primary_hpss' |
| **Unplanned_Visits_READM_30_HF** | Rate of readmission for heart failure patients | 'medicaid_ipd_percentage',  'bedsize',  'national_adi_median',  'delivery_system',  'system_member',  'svi_theme3_median',  'medicare_ipd_percentage' |
| **Unplanned_Visits_READM_30_PN** | Rate of readmission for pneumonia patients | 'bedsize',  'Device_Percent',  'delivery_system',  'system_member',  'svi_theme3_median',  'medicare_ipd_percentage',  'for_profit' |

**Table S19.** Longitudinal analysis of AI implementation and quality outcomes

Linear mixed-effects models were used to assess quality metric trajectories over 14 quarters (2022–2025), adjusting for selected covariates. Differential slopes were estimated by AI implementation level (AI-0: no models; AI-1: non-AI models; AI-2: AI models), with AI-0 as the reference. False discovery rate (FDR) correction was applied for multiple testing. Statistically significant outcomes are shown in bold; the direction (+/–) indicates favorability (e.g., +, reduced mortality; –, increased mortality).

| **Outcome** | **Estimated slope** | | | **A2-A0 Slope difference** | | | | **A1-A0 Slope difference** | | | |
| --- | --- | --- | --- | --- | --- | --- | --- | --- | --- | --- | --- |
|  | *A0 (ref)* | *A1* | *A2* | *A2-A0* | *Adj P* | *t-stat* | *Direction* | *A1-A0* | *Adj P* | *t-stat* | *Direction* |
| MORT-30-PN | 0.77 | 0.50 | 0.46 | -0.31  [-0.34, -0.27] | **<0.0001** | -16.21 | + | -0.27  [-0.32, -0.22] | **<0.0001** | -11.48 | + |
| Total HAC Score | 0.029 | -0.050 | 0.015 | -0.014  [-0.024,  -0.004] | **0.006** | -2.80 | + | -0.077  [-0.089,  -0.065] | **<0.0001** | -12.51 | + |
| READM-30-HF-HRRP | -0.0017 | 0.0013 | 0.00084 | 0.0022 [0.0014,  0.0035] | **<0.0001** | 4.70 | - | 0.0030  [0.0014, 0.0035] | **<0.0001** | 4.75 | - |
| READM-30-PN-HRRP | -0.0008 | -0.0007 | 0.0004 | 0.0013  [0.0003, 0.0022] | **0.02** | 2.54 | - | 0.0006  [-0.0011, 0.0012] | 0.97 | 0.10 | - |
| MSPB-1 | 0.00070 | 0.00049 | 0.00033 | -0.00037  [-0.00138, 0.00064] | 0.51 | -0.71 | + | -0.00021  [-0.00142, 0.00101] | 0.74 | -0.33 | + |
| HCP-COVID-19 | -31.30 | -32.70 | -33.24 | -1.94  [-2.78, -1.10] | **<0.0001** | -4.53 | - | -1.40  [-2.43, -0.37] | **0.01** | -2.66 | - |
| IMM-3 | -2.13 | -1.87 | -1.44 | 0.69  [0.51, 0.86] | **<0.0001** | 7.70 | + | 0.26  [0.038, 0.48] | **0.03** | 2.29 | + |
| OP-18b | 2.11 | 2.86 | 3.06 | 0.95  [0.64, 1.26] | **<0.0001** | 6.00 | - | 0.75  [0.35, 1.14] | **0.0004** | 3.71 | - |
| OP-22 | 0.21 | 0.20 | 0.27 | 0.060  [0.033, 0.086] | **<0.0001** | 4.41 | - | -0.015  [-0.048, 0.018] | 0.46 | -0.88 | + |
| SAFE-USE-OF-OPIOIDS | -1.62 | -1.16 | -0.70 | 0.92  [0.77, 1.06] | **<0.0001** | 12.09 | - | 0.46  [0.27, 0.64] | **<0.0001** | 4.95 | - |
| SEP-1 | 1.20 | 1.27 | 2.10 | 0.90  [0.71, 1.10] | **<0.0001** | 8.93 | - | 0.069  [-0.17, 0.31] | 0.65 | 0.56 | - |
| SEV-SEP-3HR | 0.53 | 0.14 | -0.88 | 0.35  [0.21, 0.49] | **<0.0001** | 4.79 | - | -0.39  [-0.56, -0.22] | **<0.0001** | -4.41 | + |
| EDAC-30-HF | 0.40 | -0.13 | -0.39 | -0.78  [-1.12, -0.45] | **<0.0001** | -4.56 | + | -0.52  [-0.93, -0.12] | **0.02** | -2.51 | + |
| EDAC-30-PN | -0.029 | -0.095 | -0.364 | -0.34  [-0.64, -0.03] | **0.036** | -2.15 | + | -0.067  [-0.448, 0.314] | 0.74 | -0.34 | + |
| OP-32 | -0.52 | -0.54 | -0.47 | 0.043  [0.019, 0.065] | **<0.0001** | 3.67 | - | -0.022  [-0.050, 0.005] | 0.17 | -1.56 | + |
| READM-30-HF | -0.71 | -0.64 | -0.65 | 0.060  [0.040, 0.079] | **<0.0001** | 6.03 | - | 0.066  [0.042, 0.089] | **<0.0001** | 5.47 | - |
| READM-30-PN | -0.19 | -0.20 | -0.20 | -0.005  [-0.020, 0.009] | 0.51 | -0.66 | + | -0.009  [-0.027, 0.009] | 0.45 | -0.91 | - |

### **Supplementary Figures**

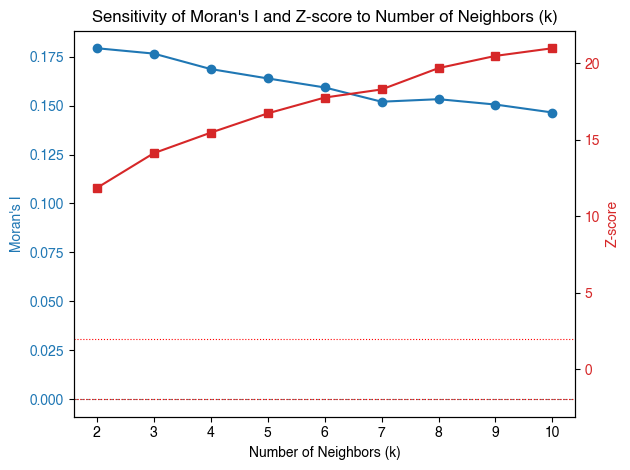

**Figure S1.** Sensitivity analysis for parameter k-nearest-neighbor selection

Moran's I decreased with increasing k, indicating weaker spatial clustering at larger neighborhood sizes. Whereas, Z-scores increased, reflecting stronger statistical power. We selected *k* = 6 to optimize this trade-off, yielding significant spatial autocorrelation while preserving detection of localized clustering patterns.

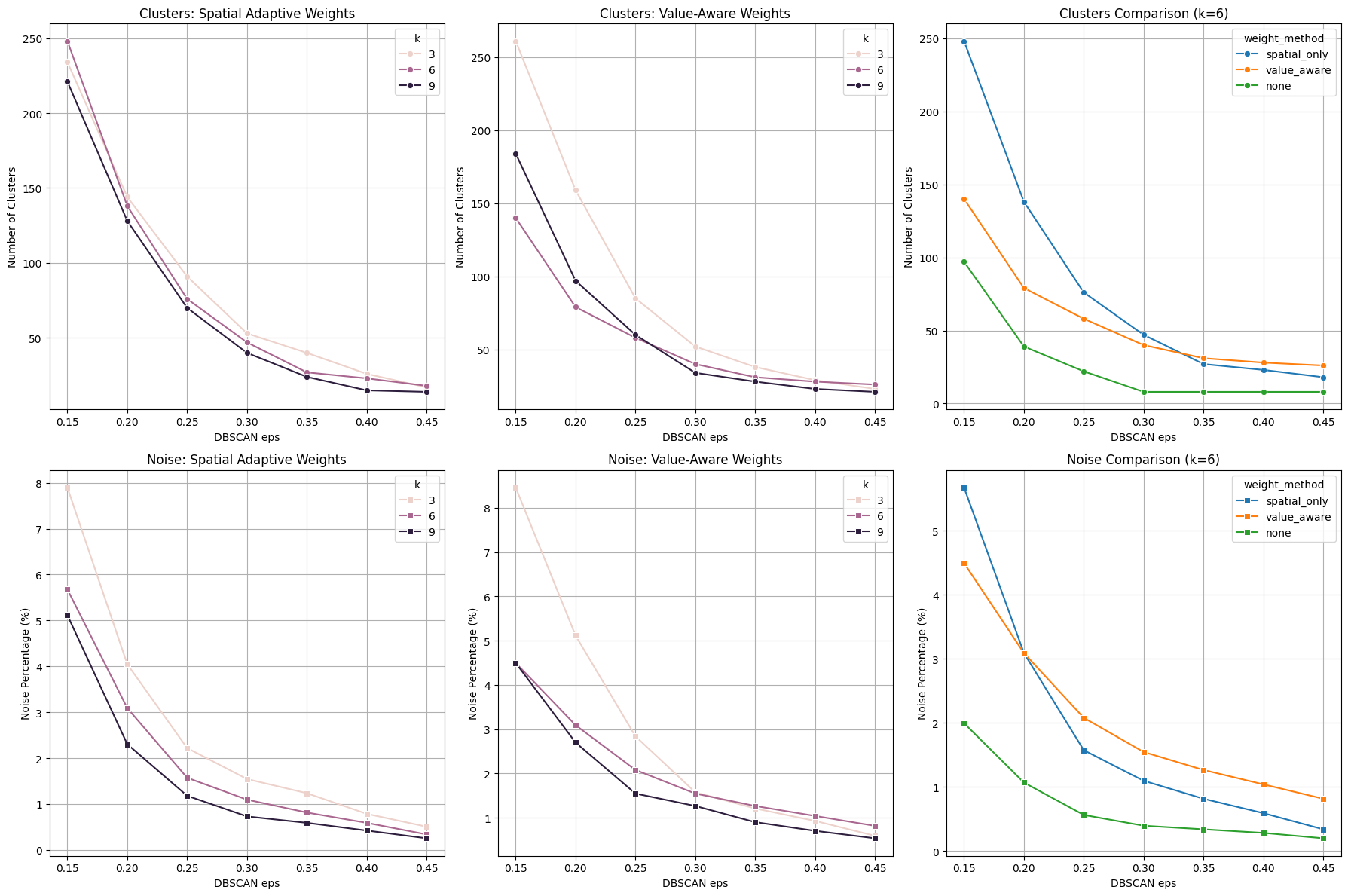

**Figure S2.** DBSCAN parameter tuning and sensitivity analysis

We selected eps=0.35 and k=6 based on parameter optimization balancing cluster granularity and noise reduction. This configuration yielded approximately 50 meaningful clusters with minimal noise (<2%), capturing spatial patterns without over-fragmentation while maintaining consistency with our spatial autocorrelation analysis (k=6).

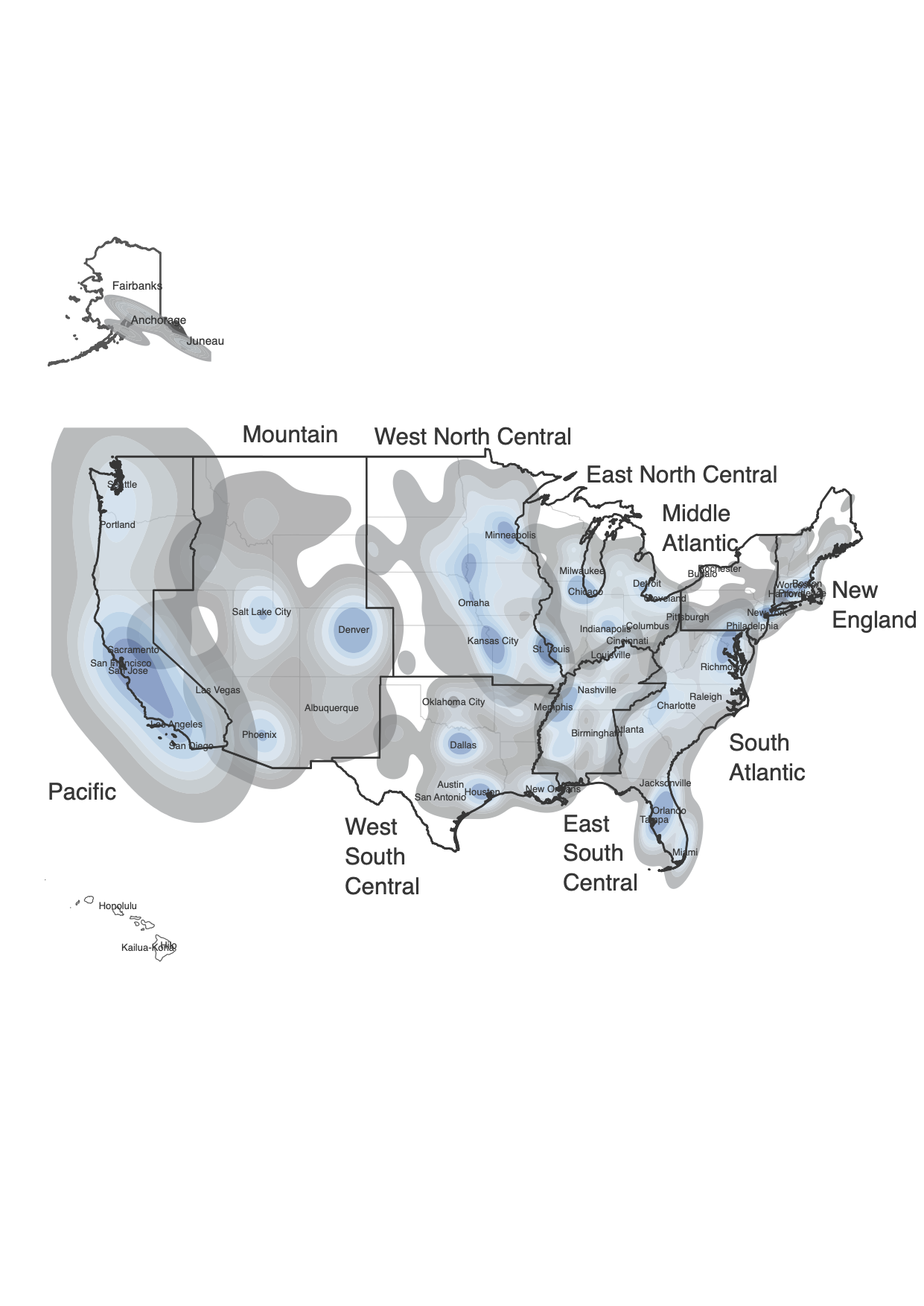

**Figure S3.** Heatmap of AI implementation scores across hospitals

Heat map shows the spatial concentration of hospitals with AI implementation, with darker shading indicating higher concentrations. Census division boundaries are outlined in black. The map reveals pronounced regional clustering, with notable concentrations in metropolitan areas and coastal regions, while some inland and rural areas show lower AI adoption density.

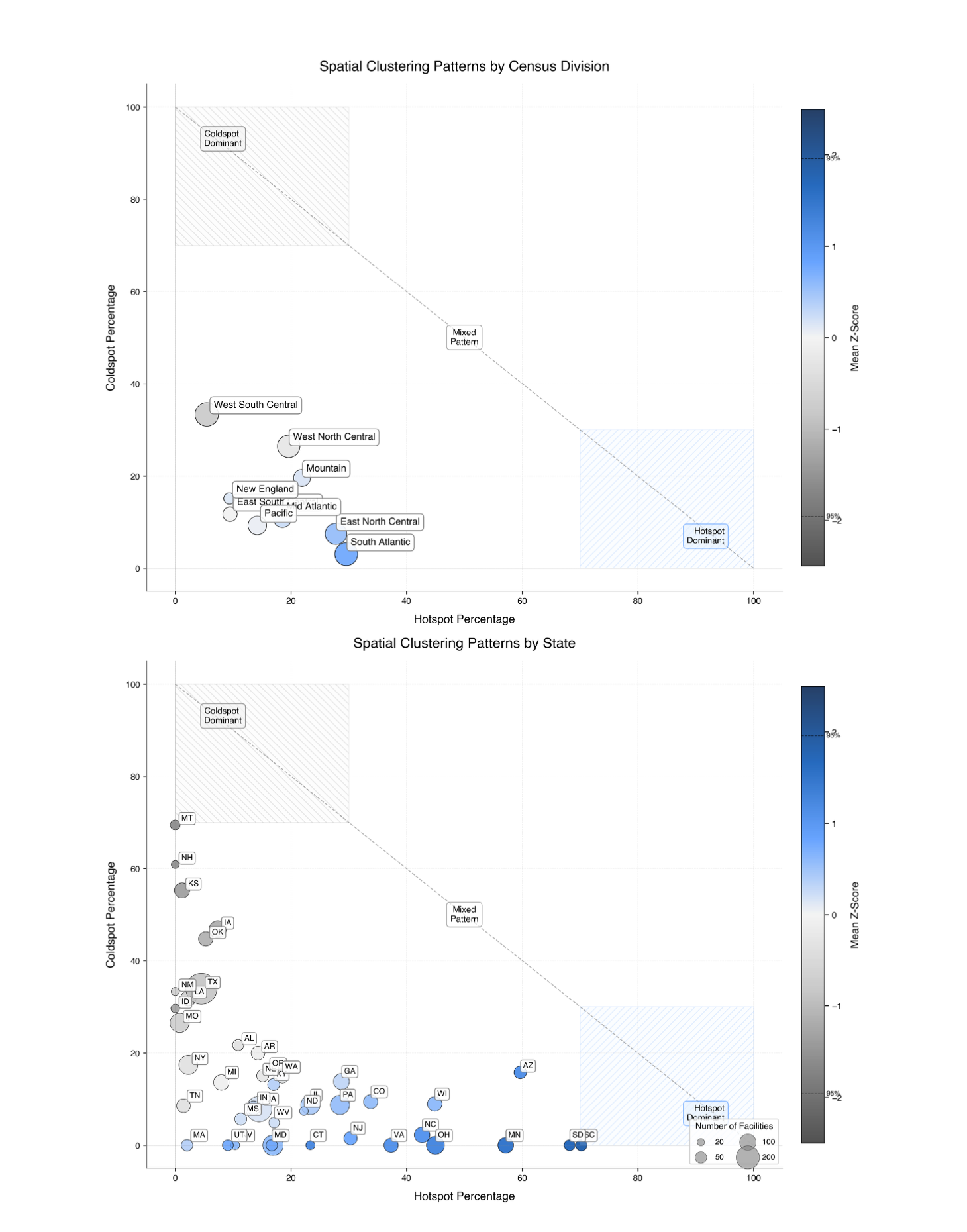

**Figure S4.** Hotspot/coldspot summary for AI implementation base score by census division and state

Scatter plots show the relationship between hotspot percentage (x-axis) and coldspot percentage (y-axis) for AI implementation base score. Circle size represents the number of hospitals, and color intensity indicates mean Z-score from Getis-Ord Gi* analysis. The diagonal line separates regions into coldspot-dominant (upper left), hotspot-dominant (lower right), and mixed pattern areas.

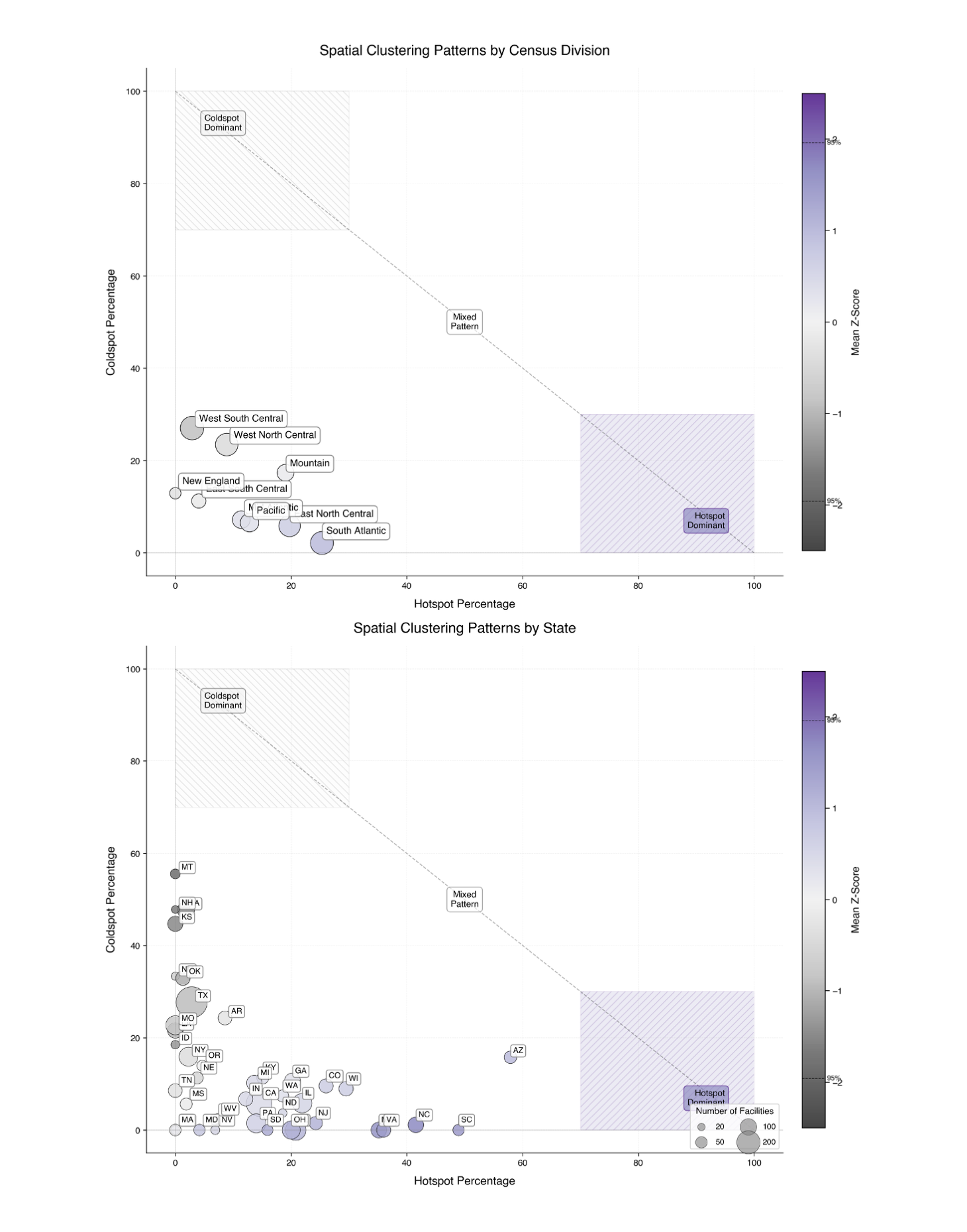

**Figure S5.** Hotspot/coldspot summary for model breadth score by census division and state

Scatter plots show the relationship between hotspot percentage (x-axis) and coldspot percentage (y-axis) for model implementation breadth score. Circle size represents the number of hospitals, and color intensity indicates mean Z-score from Getis-Ord Gi* analysis. The diagonal line separates regions into coldspot-dominant (upper left), hotspot-dominant (lower right), and mixed pattern areas.

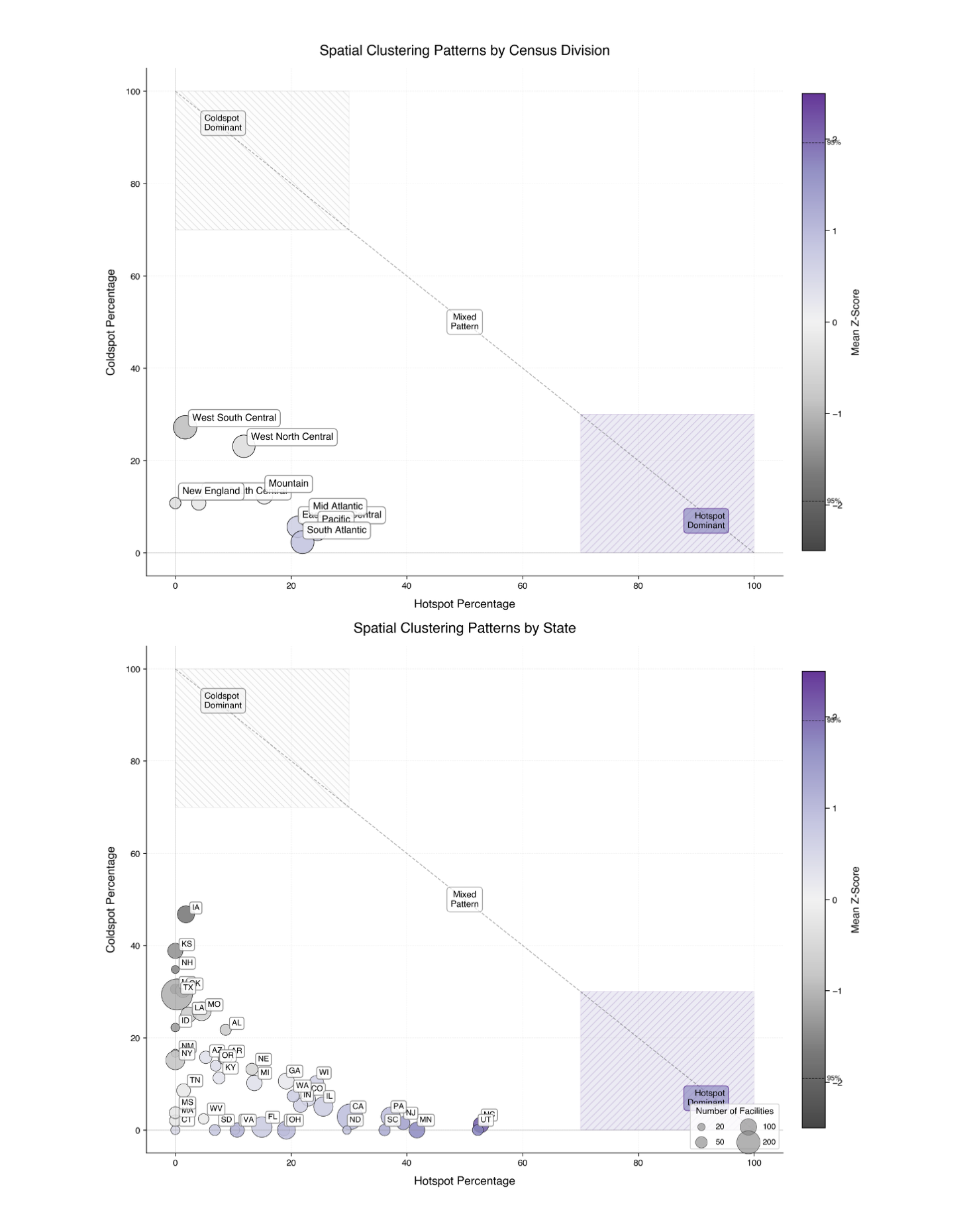

**Figure S6** Hotspot/coldspot summary for model development score by census division and state

Scatter plots show the relationship between hotspot percentage (x-axis) and coldspot percentage (y-axis) for model implementation development score. Circle size represents the number of hospitals, and color intensity indicates mean Z-score from Getis-Ord Gi* analysis. The diagonal line separates regions into coldspot-dominant (upper left), hotspot-dominant (lower right), and mixed pattern areas.

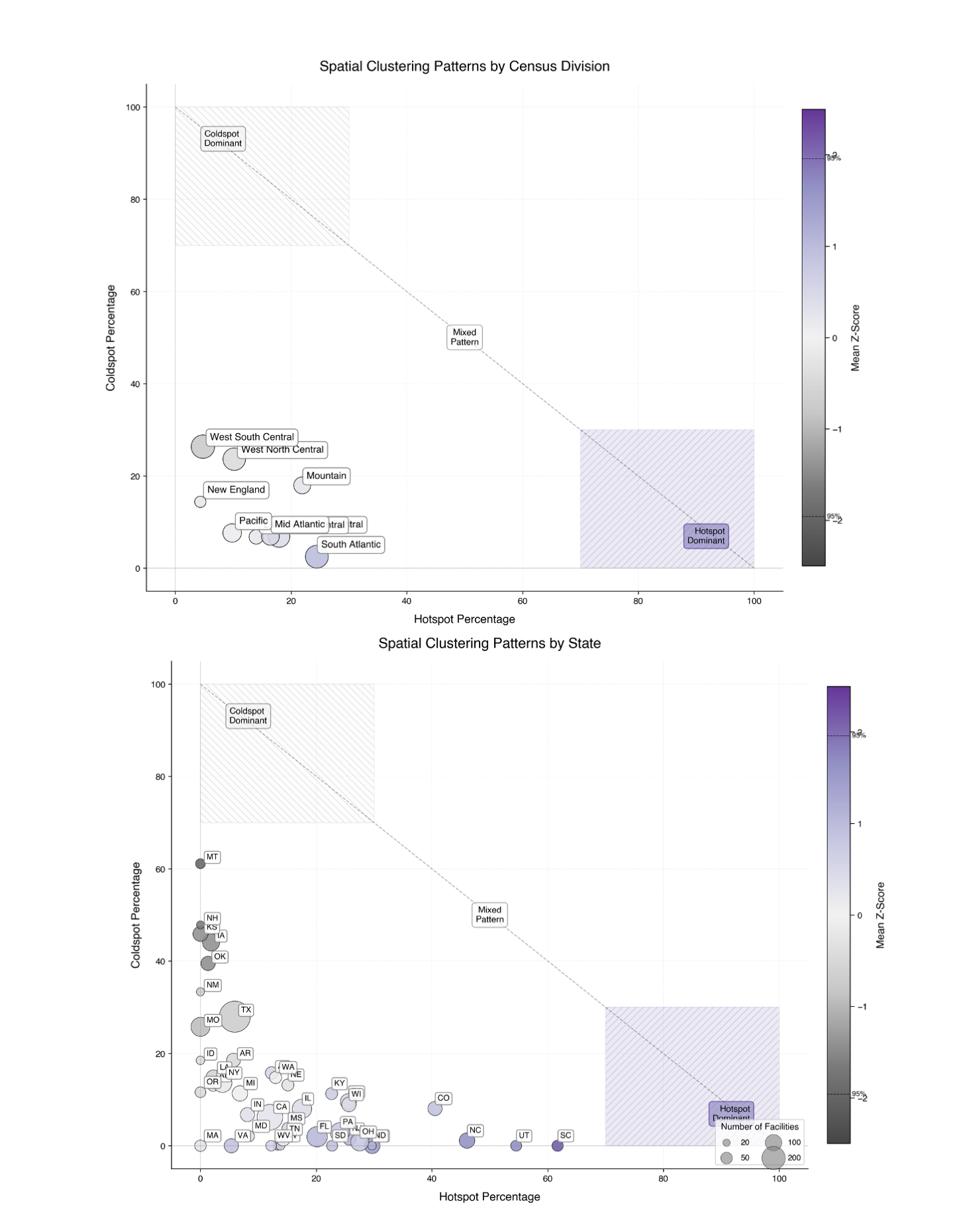

**Figure S7** Hotspot/coldspot summary for model evaluation 2023 score by census division and state

Scatter plots show the relationship between hotspot percentage (x-axis) and coldspot percentage (y-axis) for model implementation evaluation 2023 score. Circle size represents the number of hospitals, and color intensity indicates mean Z-score from Getis-Ord Gi* analysis. The diagonal line separates regions into coldspot-dominant (upper left), hotspot-dominant (lower right), and mixed pattern areas.

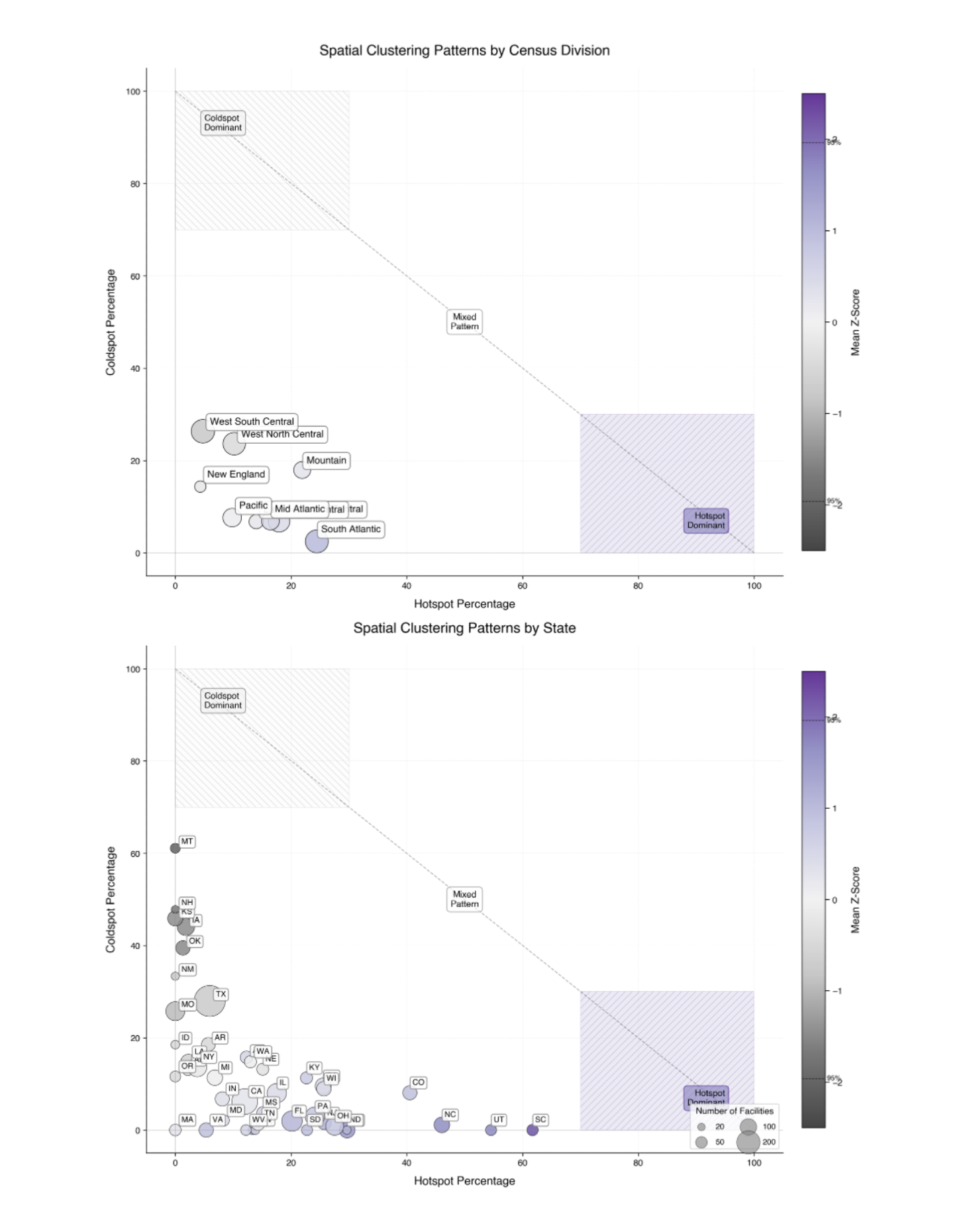

**Figure S8** Hotspot/coldspot summary for model evaluation 2024 score by census division and state

Scatter plots show the relationship between hotspot percentage (x-axis) and coldspot percentage (y-axis) for model implementation evaluation 2024 score. Circle size represents the number of hospitals, and color intensity indicates mean Z-score from Getis-Ord Gi* analysis. The diagonal line separates regions into coldspot-dominant (upper left), hotspot-dominant (lower right), and mixed pattern areas.

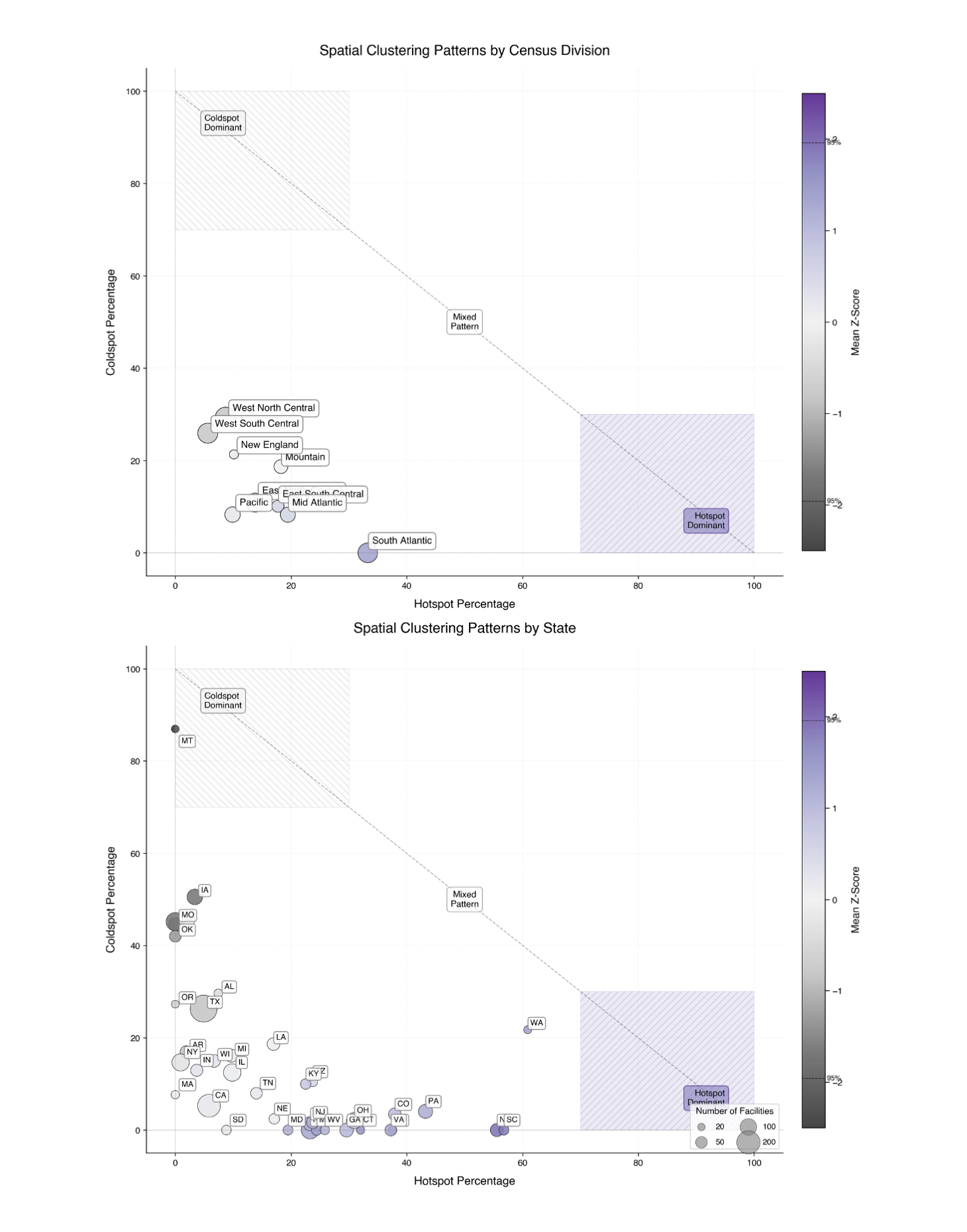

**Figure S9** Hotspot/coldspot summary for LLM readiness score score by census division and state

Scatter plots show the relationship between hotspot percentage (x-axis) and coldspot percentage (y-axis) for LLM readiness score. Circle size represents the number of hospitals, and color intensity indicates mean Z-score from Getis-Ord Gi* analysis. The diagonal line separates regions into coldspot-dominant (upper left), hotspot-dominant (lower right), and mixed pattern areas.
